# TILTomorrow today: dynamic factors predicting changes in intracranial pressure treatment intensity after traumatic brain injury

**DOI:** 10.1101/2024.05.14.24307364

**Authors:** Shubhayu Bhattacharyay, Florian D van Leeuwen, Erta Beqiri, Cecilia Åkerlund, Lindsay Wilson, Ewout W Steyerberg, David W Nelson, Andrew I R Maas, David K Menon, Ari Ercole, the CENTER-TBI investigators and participants

## Abstract

Practices for controlling intracranial pressure (ICP) in traumatic brain injury (TBI) patients admitted to the intensive care unit (ICU) vary considerably between centres. To help understand the rational basis for such variance in care, this study aims to identify the patient-level predictors of changes in ICP management. We extracted all heterogeneous data (2,008 pre-ICU and ICU variables) collected from a prospective cohort (*n*=844, 51 ICUs) of ICP-monitored TBI patients in the Collaborative European NeuroTrauma Effectiveness Research in TBI (CENTER-TBI) study. We developed the TILTomorrow modelling strategy, which leverages recurrent neural networks to map a token-embedded time series representation of all variables (including missing values) to an ordinal, dynamic prediction of the following day’s five-category therapy intensity level (TIL^(Basic)^) score. With 20 repeats of 5-fold cross-validation, we trained TILTomorrow on different variable sets and applied the TimeSHAP (temporal extension of SHapley Additive exPlanations) algorithm to estimate variable contributions towards predictions of next-day changes in TIL^(Basic)^. Based on Somers’ *D_xy_*, the full range of variables explained 68% (95% CI: 65–72%) of the ordinal variation in next-day changes in TIL^(Basic)^ on day one and up to 51% (95% CI: 45–56%) thereafter, when changes in TIL^(Basic)^ became less frequent. Up to 81% (95% CI: 78–85%) of this explanation could be derived from non-treatment variables (i.e., markers of pathophysiology and injury severity), but the prior trajectory of ICU management significantly improved prediction of future de-escalations in ICP-targeted treatment. Whilst there was no significant difference in the predictive discriminability (i.e., area under receiver operating characteristic curve [AUC]) between next-day escalations (0.80 [95% CI: 0.77–0.84]) and de-escalations (0.79 [95% CI: 0.76– 0.82]) in TIL^(Basic)^ after day two, we found specific predictor effects to be more robust with de-escalations. The most important predictors of day-to-day changes in ICP management included preceding treatments, age, space-occupying lesions, ICP, metabolic derangements, and neurological function. Serial protein biomarkers were also important and may serve a useful role in the clinical armamentarium for assessing therapeutic needs. Approximately half of the ordinal variation in day-to-day changes in TIL^(Basic)^ after day two remained unexplained, underscoring the significant contribution of unmeasured factors or clinicians’ personal preferences in ICP treatment. At the same time, specific dynamic markers of pathophysiology associated strongly with changes in treatment intensity and, upon mechanistic investigation, may improve the timing and personalised targeting of future care.

## Introduction

When traumatic brain injury (TBI) patients are admitted to the intensive care unit (ICU), a core focus of their care is to protect and promote potential recovery in brain tissue by either preventing or mitigating raised intracranial pressure (ICP).^1^ To date, the heterogeneous pathophysiological mechanisms that elevate ICP after TBI are not sufficiently characterised for patient-tailored treatment (i.e., precision medicine).^2,3^ Therefore, consensus-based guidelines^4,5^ encourage a precautionary, stepwise approach^6^ to ICP management, in which therapeutic intensity – defined by the perceived risk and complexity of each treatment plan – is incrementally escalated until adequate ICP control is achieved. The overall intensity of a patient’s ICP management can be measured on the latest Therapy Intensity Level (TIL) scale,^7^ which was developed by the interagency TBI Common Data Elements (CDE) scheme^8^ and prospectively validated thereafter.^7,9^

An analysis of high-TIL treatment administration across 52 ICUs participating in the Collaborative European NeuroTrauma Effectiveness Research in TBI (CENTER-TBI) study^10,11^ revealed frequent deviation from the recommended stepwise approach, even with ICP monitoring.^12^ In fact, there was substantial between-centre variation in ICP management (according to TIL) without commensurate variation in six-month functional outcome on the Glasgow Outcome Scale – Extended (GOSE).^13,14^ Baseline injury severity factors, imaging results, and ICP explained only 8.9% of the pseudo-variance in dichotomised high-TIL treatment use.^12^ These results raised the questions about whether contemporary ICP management is performed in a systematic, rational manner in practice and whether some patients are being exposed to unnecessary risks with high-TIL therapies. Answering these questions requires consideration of a patient’s full, time-varying clinical course as well as a more detailed representation of different levels of the TIL scale.

As a first step towards answering the questions above, we aim to identify factors associated with ICP-targeted treatment decisions on an individual patient level. Expanding upon our previous work,^13,15^ we propose a modelling strategy (TILTomorrow) which dynamically predicts next-day TIL^(Basic)^ – the five-category version of TIL – from all pre-ICU and ICU data prospectively recorded for the CENTER-TBI study (Fig. 1). Our primary objective in developing TILTomorrow was to determine how well a patient’s full clinical course can predict upcoming changes in ICP treatment intensity. Our second objective was to estimate the differential contribution of pathophysiological severity, the preceding trajectory of treatment, and unmeasured factors (e.g., personal treatment preferences) towards explanation of next-day changes made to TIL^(Basic)^. Our third objective was to mine the full dataset for dynamic predictors of day-to-day changes in TIL^(Basic)^.

**Fig. 1.**
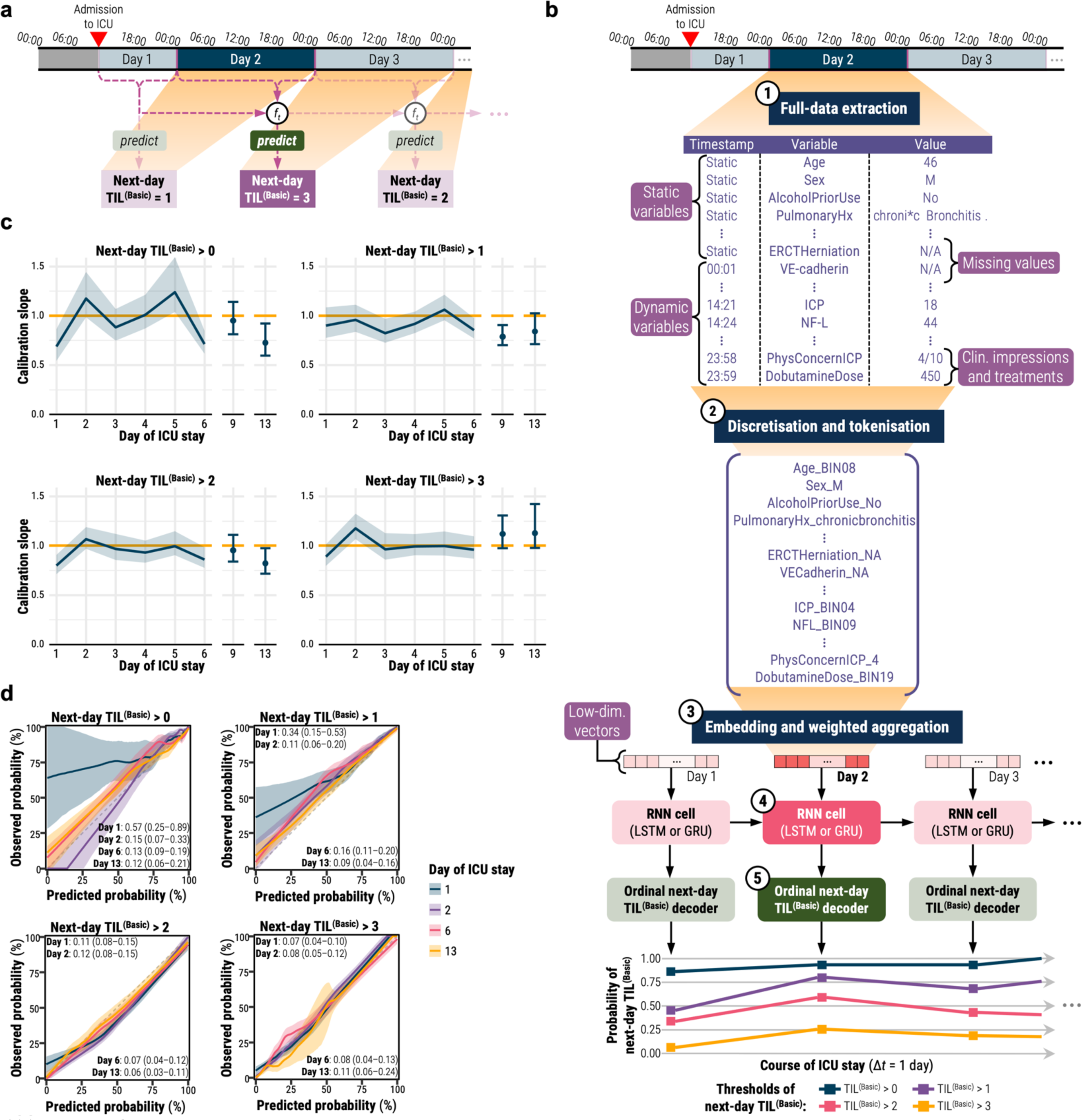
TILTomorrow prediction task and modelling strategy. All shaded regions surrounding curves are 95% confidence intervals derived using bias-corrected bootstrapping (1,000 resamples) to represent the variation across the patient population and across the 20 repeated five-fold cross-validation partitions. (**a**) Illustration of the TILTomorrow dynamic prediction task on a sample patient’s timeline of ICU stay. The objective of the task is to predict the next-day TIL^(Basic)^ score at each calendar day of a patient’s ICU stay. The prediction is dynamic, updated for each calendar day, and must account for temporal variation of variables across all preceding days using a time-series model (*f_t_*). (**b**) Illustration of the TILTomorrow modelling strategy on a sample patient’s timeline of ICU stay. Each patient’s ICU stay is first discretised into non-overlapping time windows, one for each calendar day. From each time window, values for up to 979 dynamic variables were combined with values for up to 1,029 static variables to form the variable set. The variable values were converted to tokens by discretising numerical values into 20-quantile bins from the training set and removing special formatting from text-based entries. Through an embedding layer, a vector was learned for each token encountered in the training set, and tokens were replaced with these vectors. A positive relevance weight, also learned for each token, was used to weight-average the vectors of each calendar day into a single, low-dimensional vector. The sequence of low-dimensional vectors representing a patient’s ICU stay were fed into a gated recurrent neural network (RNN). The RNN outputs were then decoded at each time window into an ordinal prognosis of next-day TIL^(Basic)^ score. The highest-intensity treatments associated with each threshold of TIL^(Basic)^ are decoded in Table 1. (**c**) Probability calibration slope, at each threshold of next-day TIL^(Basic)^, for models trained on the full variable set. The ideal calibration slope of one is marked with a horizontal orange line. (**d**) Ordinal probability calibration curves at four different days after ICU admission. The diagonal dashed line represents the line of perfect calibration. The values in each panel correspond to the maximum absolute error (95% confidence interval) between the curve and the perfect calibration line. Abbreviations: CT=computerised tomography, ER=emergency room, *f_t_*=time-series model, GRU=gated recurrent unit, Hx=history, ICP=intracranial pressure, ICU=intensive care unit, LSTM=long short-term memory, N/A=not available, NF-L=neurofilament light chain, SES=socioeconomic status, TIL=Therapy Intensity Level, TIL^(Basic)^=condensed, five-category TIL scale as defined in Table 1, VE=vascular endothelial.

**Table 1.**
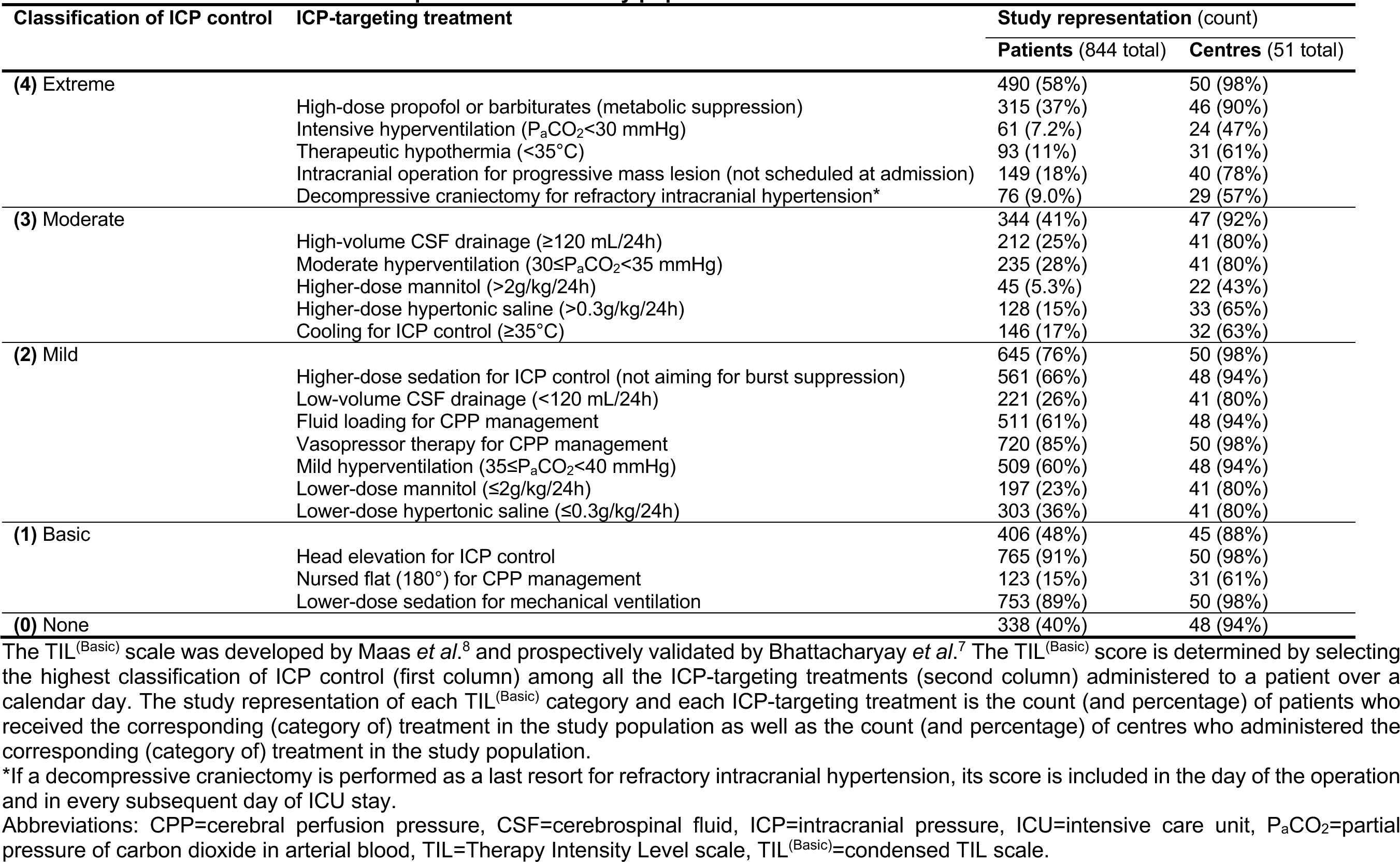
TIL^(Basic)^ scale treatments and representation in study population.

## Methods

### Study design and participants

CENTER-TBI is a longitudinal, observational cohort study (NCT02210221) involving 65 medical centres across 18 European countries and Israel.^10,11^ Patients were recruited between 19 December 2014 and 17 December 2017 if they met the following criteria: (1) presentation within 24 hours of a TBI, (2) clinical indication for a computerised tomography (CT) scan, and (3) no severe pre-existing neurological disorder. In accordance with relevant laws of the European Union and the local country, ethical approval was obtained for each site, and written informed consent by the patient or legal representative was documented electronically. The list of sites, ethical committees, approval numbers, and approval dates can be found online: https://www.center-tbi.eu/project/ethical-approval. The project objectives and design of CENTER-TBI have been described in detail previously.^10,11^

In this work, we apply the following additional inclusion criteria: (1) primary admission to the ICU, (2) at least 16 years old at ICU admission, (3) at least 24 hours of ICU stay, (4) invasive ICP monitoring, (5) no decision to withdraw life-sustaining therapies (WLST) on the first day of ICU stay, and (6) availability of daily TIL assessments from at least two consecutive days.

### Therapy intensity level (TIL)

The endpoint for the TILTomorrow dynamic prediction task (Fig. 1a) is the next-day TIL^(Basic)^ score. The TIL^(Basic)^ scale was developed through an international expert panel to serve as a five-category summary of the full, 38-point TIL score.^8^ TIL^(Basic)^ categorises overall ICP treatment intensity over a given period of time by selecting the highest classification of ICP control amongst all treatments administered in that period of time, as defined in Table 1. By convention, a decompressive craniectomy for refractory intracranial hypertension is scored with TIL^(Basic)^=4 (i.e., extreme ICP control) for every subsequent timepoint. As described later, we account for this effect in our analysis by: (1) referencing TILTomorrow performance against simply carrying forward the last-available TIL^(Basic)^ score and against models trained without treatment (e.g. incidence of decompressive craniectomy) or clinician-impression (e.g., reason for decompressive craniectomy) variables, and (2) focusing only on variables that occur at least a day before a change in TIL^(Basic)^. Since daily use of TIL^(Basic)^ was prospectively validated,^7^ we calculate the TIL^(Basic)^ score over each available calendar day of a patient’s ICU stay. For the CENTER-TBI study, information pertaining to the TIL^(Basic)^ treatments (Table 1) was recorded on days 1–7, 10, 14, 21, and 28 of ICU stay. TIL^(Basic)^ score calculations were excluded on or after the day of any WLST decision. As an overall summary metric, we also calculated TIL^(Basic)^_median_ – the median of the daily TIL^(Basic)^ scores over days 1–7 of ICU stay.

We elected not to use the full TIL score as the model endpoint since it is a point-sum (rather than a truly categorical) score, and the same value changes in TIL can be the result of changing treatments across different intensities. For instance, administering head elevation, low-volume cerebrospinal fluid drainage, and low-dose mannitol is numerically ‘equivalent’ to performing a last-resort decompressive craniectomy.^7^ On the contrary, changes in TIL^(Basic)^ correspond to transitions across specific, interpretable bands of treatment intensity (Table 1).

### Model variables

We extracted all variables collected before and during ICU stays for the CENTER-TBI core study^11^ (v3.0, ICU stratum) using Opal database software.^16^ These variables were sourced from medical records and online test results and include structured (i.e., numerical, binary, or categorical), unstructured (i.e., free text), and missing values. We manually excluded variables which explicitly indicate death or WLST (Supplementary Table S1), and, if a decision to WLST was made during any point of a patient’s ICU stay, we only extracted model variables before the timestamp of WLST decision. We also added features extracted from automatically segmented and expert-corrected high-resolution CT and magnetic resonance (MR) images. These features correspond to the type, location, and volume of space-occupying lesions, and the process of their extraction has been described in detail previously.^17,18^ In total, we included 2,008 variables: 1,029 static (i.e., fixed at ICU admission) variables and 979 dynamic variables (i.e., collected during ICU stay) with varying sampling frequencies. We qualitatively organised the variables into the nine categories listed in Table 2 and further indicated whether variables represented an intervention during ICU admission (e.g., administration and type of glucose management) or a physician-based impression (e.g., reason for not pursuing intracranial surgery following CT scan, Supplementary Table S2). Descriptions for each of the variables can be viewed online at the CENTER-TBI data dictionary: https://www.center-tbi.eu/data/dictionary.

**Table 2.** Variable count per category and subtype.

| Category | Example variable | Count by subtypes |  |  |  |
| --- | --- | --- | --- | --- | --- |
|  |  | All | Static | Dynamic | Interventions and physician impressions |
| Demographics and socioeconomic status | Years of formal education | 22 | 22 | 0 | 0 |
| Medical and behavioural history | Number of prior TBIs or concussions | 186 | 186 | 0 | 0 |
| Injury characteristics and severity | Airbag deployed during accident | 84 | 84 | 0 | 0 |
| Emergency care and ICU admission | Blood transfusion in ER | 234 | 234 | 0 | 14 |
| Brain imaging reports | Cortical sulcal effacement | 939 | 425 | 514 | 19 |
| Laboratory measurements | Serum level of UCH-L1 | 228 | 75 | 153 | 6 |
| ICU medications and management | Vasopressor dose | 141 | 3 | 138 | 127 |
| ICU vitals and assessments | Types of seizures in past day | 125 | 0 | 125 | 0 |
| Surgery and neuromonitoring | Ventriculostomy for CSF drainage | 49 | 0 | 49 | 39 |
| **Total** |  | **2008** | **1029** | **979** | **205** |

Data represent the number of subtype (*column*) variables per category (*row*).
Abbreviations: CSF=cerebrospinal fluid, ER=emergency room, ICU=intensive care unit, SBP=systolic blood pressure, TBI=traumatic brain injury, UCH-L1=ubiquitin carboxy-terminal hydrolase L1.

### TILTomorrow modelling strategy

Whilst strong predictors of functional outcome after TBI are known, this is not the case for TIL. Thus, the TILTomorrow modelling strategy was designed to include *all* static and dynamic variables from CENTER-TBI to produce an evolving prediction of the next calendar day’s TIL^(Basic)^ over each patient’s ICU stay. The large number of variables precludes building such a model by manual feature extraction, motivating our flexible tokenisation-and-embedding approach with no constraints on the number or type of variables per patient. We trained models, through supervised machine learning, with three main components based on our prior studies^13,15,19^: (1) a token-embedding encoder, (2) a gated recurrent neural network (RNN), and (3) an ordinal endpoint output layer. We created 100 partitions of our patient population for repeated *k*-fold cross-validation (20 repeats, 5 folds) with 15% of each training set randomly set aside as an internal validation set.

ICU stays were partitioned into non-overlapping time windows, one per calendar day (Fig. 1a). Static variables were carried forward across all windows (Fig. 1b). All variables were tokenised through one of the following methods: (1) for categorical variables, appending the value to the variable name, (2) for numerical variables, learning the training set distribution and discretising into 20 quantile bins, (3) for text-based entries, removing all special characters, spaces, and capitalisation from the text and appending to the variable name, and (4) for missing values, creating a separate token to designate missingness (Fig. 1b). We selected 20 quantile bins for discretisation based on optimal performance in our previous work.^13,19^ By labelling missing values with separate tokens instead of imputing them, the models could learn potentially significant patterns of missingness and integrate a diverse range of missing data without needing to validate the assumptions of imputation methods on each variable.^20^ During training, the models learned a low-dimensional vector (of either 128, 256, 512, or 1,024 units) and a ‘relevance’ weight for each token in the training set. Therefore, models would take the unique tokens from each time window of a patient, replace them with the corresponding vectors, and average the vectors – each weighted by its corresponding relevance score – into a single vector per time window (Fig. 1b).

Each patient’s sequence of low-dimensional vectors then fed into a gated RNN – either a long short-term memory (LSTM) network or a gated recurrent unit (GRU) – to output another vector per time window. In this manner, the models learned temporal patterns of variable interactions from training set ICU records and updated outputs with each new time window of data. Finally, each RNN output vector was decoded with a multinomial (i.e., softmax) output layer to return a probability at each threshold of next-day TIL^(Basic)^ over time (Fig. 1b). From these outputs, we also calculated the probabilities of TIL^(Basic)^ decreasing, staying the same, or increasing tomorrow in relation to the last available TIL^(Basic)^ score (Supplementary Methods S1). Please note that both threshold-level probability estimates and estimated probabilities of next-day changes in TIL^(Basic)^ are derived from the outputs of the same model, as described in Supplementary Methods S1.

The combinations of hyperparameters – in addition to those already mentioned (embedding vector dimension and RNN type) – and the process of their optimisation in the internal validation sets are reported in Supplementary Methods S2–S3.

### Model and information evaluation

All metrics, curves, and associated confidence intervals (CIs) were calculated on the testing sets using the repeated Bootstrap Bias Corrected Cross-Validation (BBC-CV) method,^21^ as described in Supplementary Methods S2. We calculated metrics and CIs at each day directly preceding a day of TIL assessment in our study population (i.e., days 1–6, 9, 13, 20, and 27).

The reliability of model-generated prediction trajectories was assessed through the calibration of output probabilities at each threshold of next-day TIL^(Basic)^. Using the logistic recalibration framework,^22^ we first measured calibration slope. Calibration slope less(/greater) than one indicates overfitting(/underfitting).^22^ Additionally, we examined smoothed probability calibration curves to detect miscalibrations that might have been overlooked by the logistic recalibration framework.^22^

To evaluate prediction discrimination performance, we calculated the area under the receiver operating characteristic curve (AUC) at each threshold of next-day TIL^(Basic)^. These AUCs are interpreted as the probability of the model correctly discriminating a patient whose next-day TIL^(Basic)^ is above a given threshold from one with next-day TIL^(Basic)^ below. Moreover, we calculated the AUC for prediction of next-day escalation and de-escalation in TIL^(Basic)^. In this case, the AUC represents the probability of the model correctly discriminating a patient who experienced a day-to-day (de-)escalation in TIL^(Basic)^ from one who did not.

We also assessed the information quality achieved by the combination of our modelling strategy and the CENTER-TBI variables in predicting next-day changes in TIL^(Basic)^ by calculating Somers’ *D_xy_*.^23^ In our context, Somers’ *D_xy_* is interpreted as the proportion of ordinal variation in day-to-day changes of TIL^(Basic)^ that is explained by the variation in model output.^24^ The calculation of Somers’ *D_xy_* is detailed in Supplementary Methods S4.

We compared the performance of the TILTomorrow modelling strategy trained on the following factors to test their differential contribution to prediction: (1) the full variable set [2,008 variables], (2) all variables excluding physician-based impressions and treatments (e.g., all variables related to TIL) [1,803 variables], and (3) only static variables repeated in each time window [1,029 variables]. Our rationale for these ablated variable sets was to estimate the extent to which: (1) predictable trajectories of care – independent of other measured factors – influence treatment planning and (2) ICP treatments are responding to recorded events that occur over a patient’s ICU stay. To serve as our reference for model comparison, we also calculated the performance achieved by simply carrying over the last available TIL^(Basic)^ for prediction of next-day TIL^(Basic)^. This reference performance accounts not only for the proportion of the population that did not change in TIL^(Basic)^ on a given day but also for the change in the assessment population caused by patient discharge over time.

### Contributors to transitions in TIL

We applied the TimeSHAP algorithm^25^ on testing set predictions to find specific variables associated with next-day changes in TIL^(Basic)^. TimeSHAP is a temporal extension of the kernel-weighted SHapley Additive exPlanations (KernelSHAP) algorithm,^26^ which estimates the relative contribution (i.e., Shapley value^27^) of each model input to a specific patient’s model output. In our case, this was done by masking sampled combinations of tokens (i.e., coalitions) leading up to a patient’s next-day change in TIL^(Basic)^ and calculating the difference in trained model output for each combination. A kernel-weighted linear regression model was then fit between binary coalition masks and resulting model outputs to estimate the Shapley value for each model input. TimeSHAP extends KernelSHAP by considering each unique combination of tokens and time windows as its own feature. Crucially, TimeSHAP made this computationally tractable for our application, in which models contain many possible tokens, by grouping low-contributing time windows in the distant past together as a single feature (i.e., temporal coalition pruning). TimeSHAP, KernelSHAP, and Shapley values are described in greater, mathematical detail in Supplementary Methods S5.

We estimated token-level Shapley values with the TimeSHAP algorithm at both one day and two days before an upcoming change in TIL^(Basic)^. Our chosen model output for TimeSHAP was the expected next-day TIL^(Basic)^ score, as defined in Supplementary Methods S5. We then calculated the difference between the estimated Shapley values of the two consecutive days for each token to derive its ΔTimeSHAP value. If a token did not exist in the window of either of the two days, then its Shapley value for that day was zero. Therefore, ΔTimeSHAP values were interpreted as the contributions of variable tokens towards the difference in model prediction of next-day TIL^(Basic)^ over the two days directly preceding the change in TIL^(Basic)^, given the patient’s full set of tokens. If a variable had a positive (or negative) ΔTimeSHAP value, it was associated with an increased likelihood of escalation (or de-escalation) in next-day treatment intensity. Moreover, since the calculation of ΔTimeSHAP values required two days of information before the change in TIL^(Basic)^, we only calculated the variable contributions to day-to-day changes in TIL^(Basic)^ that occurred after day two of ICU stay.

## Results

### Study population

Of the 4,509 patients available for analysis in the CENTER-TBI core study, 844 patients from 51 ICUs met the inclusion criteria of this work (Supplementary Fig. S1). The median ICU stay duration of our population was 14 days (*Q*_1_–*Q*_3_: 8.4–23 days) and 86% (*n*=722) stayed through at least seven calendar days. Since the regularity of TIL^(Basic)^ assessments decreased substantially after 14 days, and since less than half of the population remained in the ICU for 21 days (Supplementary Fig. S2), we focused our analysis on the first 14 days of ICU stay. Summary characteristics of the overall population as well as those stratified by whether patients had a day-to-day change in TIL^(Basic)^ over their first week in the ICU are detailed in Table 3. On average, patients who did not experience a change in TIL^(Basic)^ over their first week were significantly younger, had higher baseline ICP values, and resulted in poorer functional recovery at six months post-injury (Table 3). However, their mean ICU stay duration was not significantly different.

**Table 3.** Summary characteristics of the study population stratified by day-to-day changes in _TIL(Basic)_

| Summary characteristic | Overall<br>(*n*=844,<br>51 centres) | Day-to-day change in TIL<sup>(Basic)</sup> during first week in ICU |  |  |
| --- | --- | --- | --- | --- |
|  |  | Yes (*n*=677,<br>50 centres) | No (*n*=167,<br>40 centres) | *p*-value<sup>‡</sup> |
| Age [years] | 47 (29–61) | 48 (30–62) | 41 (27–58) | 0.047 |
| Sex: Female | 212 (25%) | 165 (24%) | 47 (28%) | 0.36 |
| Baseline Glasgow Coma Scale (*n* *=795) |  |  |  | 0.67 |
| 3–8 | 540 (68%) | 426 (67%) | 114 (71%) |  |
| 9–12 | 138 (17%) | 112 (18%) | 26 (16%) |  |
| 13–15 | 117 (15%) | 96 (15%) | 21 (13%) |  |
| Baseline CT lesions (*n* *=730) |  |  |  |  |
| Epidural haematoma | 165 (23%) | 136 (23%) | 29 (19%) | 0.36 |
| Intracerebral haemorrhage | 594 (81%) | 480 (83%) | 114 (77%) | 0.11 |
| Subdural haematoma | 465 (64%) | 368 (63%) | 97 (65%) | 0.76 |
| Traumatic subarachnoid haemorrhage | 633 (87%) | 502 (86%) | 131 (88%) | 0.73 |
| First-day mean ICP [mmHg] (*n* *=811) | 11 (7.0–15) | 10. (6.8–14) | 12 (8.2–17) | <0.001 |
| TIL<sup>(Basic)</sup><sub>median</sub> | 2 (2–4) | 2 (2–3) | 4 (2–4) | <0.001 |
| Refractory intracranial hypertension (*n* *=836) | 143 (17%) | 85 (13%) | 58 (35%) | <0.001 |
| ICU stay duration [days] | 14 (8.4–23) | 14 (8.1–23) | 14 (8.8–23) | 0.90 |
| Six-month GOSE (*n* *=738) |  |  |  | 0.018 |
| (1) Death | 181 (25%) | 139 (23%) | 42 (29%) |  |
| (2 or 3) Vegetative/lower SD | 181 (25%) | 154 (26%) | 27 (18%) |  |
| (4) Upper SD | 70 (9.5%) | 48 (8.1%) | 22 (15%) |  |
| (5) Lower MD | 122 (17%) | 96 (16%) | 26 (18%) |  |
| (6) Upper MD | 73 (10%) | 65 (11%) | 8 (5.5%) |  |
| (7) Lower GR | 55 (7.5%) | 42 (7.1%) | 13 (8.9%) |  |
| (8) Upper GR | 56 (7.6%) | 48 (8.1%) | 8 (5.5%) |  |
| Baseline prognosis<sup>†</sup> [%] (*n* *=749) |  |  |  |  |
| Pr(GOSE>1) | 85 (64–94) | 85 (66–95) | 83 (56–93) | 0.010 |
| Pr(GOSE>3) | 54 (31–75) | 54 (33–76) | 52 (24–71) | 0.019 |
| Pr(GOSE>4) | 40. (22–59) | 41 (24–60.) | 38 (16–54) | 0.010 |
| Pr(GOSE>5) | 22 (11–36) | 22 (12–38) | 19 (8.9–30.) | 0.0022 |
| Pr(GOSE>6) | 13 (6.7–21) | 13 (7.1–22) | 11 (5.2–17) | 0.0034 |
| Pr(GOSE>7) | 5.2 (2.5–9.5) | 5.4 (2.7–9.9) | 4.2 (2.2–8.6) | 0.0071 |

Data are median (Q<sub>1</sub>–Q<sub>3</sub>) for numerical characteristics and *n* (% of column group) for categorical characteristics unless otherwise indicated. Units or numerical definitions of characteristics are provided in square brackets.
\*Limited sample size of non-missing values for characteristic.
<sup>†</sup>Ordinal functional outcome prognostic scores were calculated through tokenised embedding of all clinical information in the first 24 hours of ICU stay, as described previously.<sup>15</sup>
<sup>‡</sup>*p*-values, comparing patients who experienced a day-to-day change in TIL<sup>(Basic)</sup> in the first week of ICU stay to those who did not, are derived from Welch's *t*-test for numeric variables and $\chi^2$ contingency table test for categorical variables.
Abbreviations: CT=Computerised tomography, GOSE=Glasgow Outcome Scale–Extended, GR=good recovery, ICP=intracranial pressure, ICU=intensive care unit, MD=moderate disability, $\text{Pr}(\text{GOSE}>\bullet)$ =“probability of GOSE greater than $\bullet$ at six months post-injury” as previously calculated from the first 24 hours of admission,<sup>27</sup> SD=severe disability, TIL=Therapy Intensity Level scale, $\text{TIL}^{(\text{Basic})}$ =condensed TIL scale as measured in Table 1 for each calendar day, $\text{TIL}^{(\text{Basic})}_{\text{median}}$ =median $\text{TIL}^{(\text{Basic})}$ over first week of ICU stay.

The representation of each ICP-targeting treatment and TIL^(Basic)^ score in our study is listed in Table 1. The least-represented treatment (higher-dose mannitol) was administered to 45 patients (5.3%) across 22 ICUs, whereas the least-represented TIL^(Basic)^ score (TIL^(Basic)^=1) applied to 344 patients (41%) across 47 ICUs. A decompressive craniectomy for refractory intracranial hypertension was performed in 76 patients (9.0%) across 29 ICUs, and the median timepoint for such an operation was day three (*Q*_1_–*Q*_3_: two–five) of ICU stay.

The distribution of TIL^(Basic)^ values at each day of TIL assessment and the transitions of TIL^(Basic)^ scores between days of assessment are visualised in Fig. 2a. No more than 2.4% of the population’s TIL^(Basic)^ scores were missing at any given assessment day, and the proportion of patients receiving basic-to-no ICP-targeting treatment (i.e., TIL^(Basic)^≤1) increased over time (Supplementary Fig. S2). The distribution of day-to-day changes in TIL^(Basic)^ (Fig. 2b) demonstrates that there was considerably more change in TIL^(Basic)^ from day one to day two than there was in any other pair of consecutive days. On the rest of the days in the first week, 69–75% of the population did not experience a change in TIL^(Basic)^ from one day to the next (Fig. 2b). The distribution of next-day TIL^(Basic)^ given the current day’s TIL^(Basic)^ (Supplementary Fig. S3) show that at least 79% of day-to-day therapeutic transitions happen within one TIL^(Basic)^ category, except for escalations from TIL^(Basic)^=0 and de-escalations from TIL^(Basic)^=4 from day one to two. When a change in TIL^(Basic)^ did occur, the distributions of TIL^(Basic)^ before and after the change (Supplementary Fig. S4) reflect a gradual trend towards de-escalation at later days of ICU stay as expected.

**Fig. 2.**
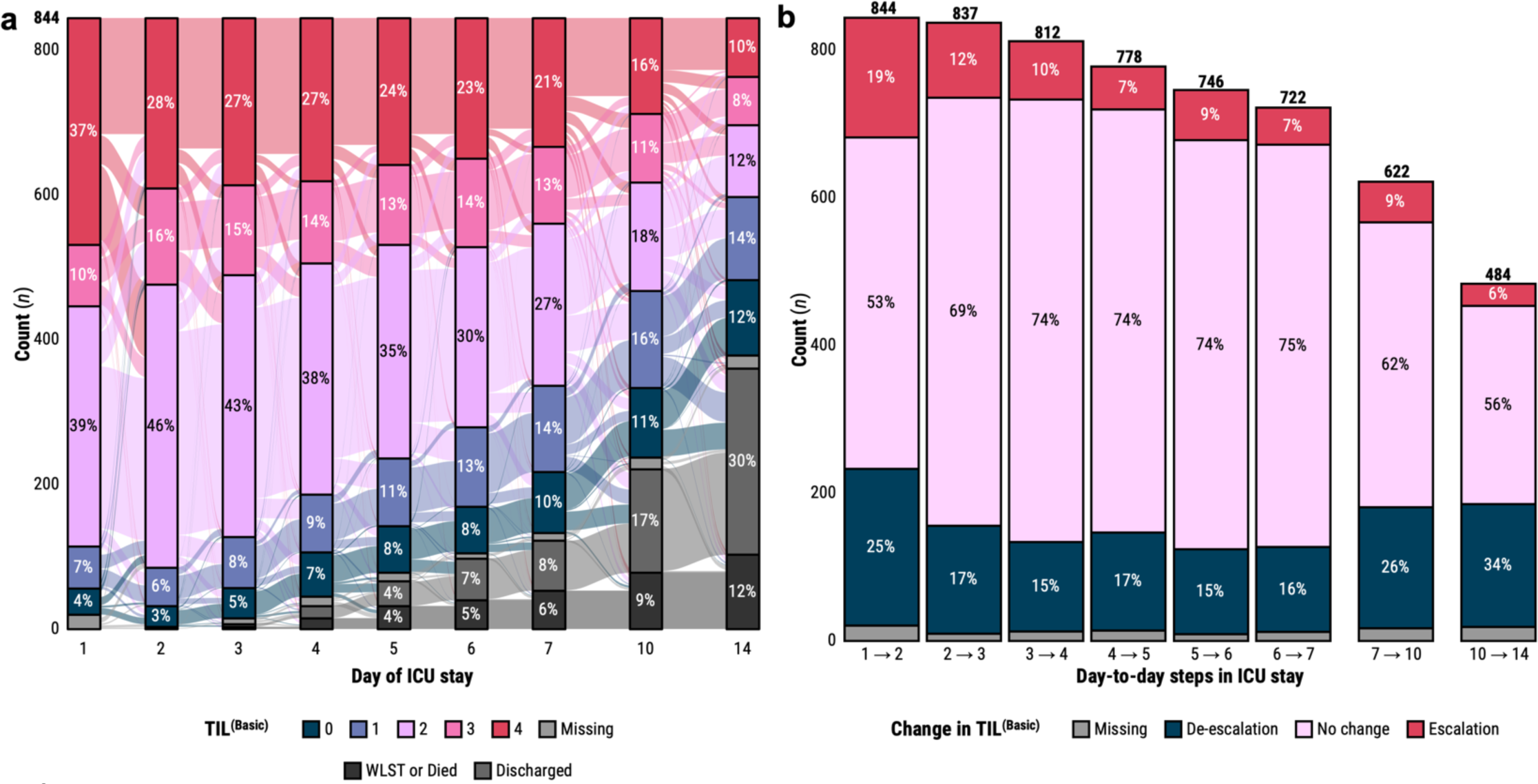
Distributions of TIL ^(Basic)^ and its day-to-day changes in the study population. (**a**) Alluvial diagram of the evolution of the TIL^(Basic)^ distribution in the study population over the assessed days of ICU stay. Percentages which round to 2% or lower are not shown. (**b**) Distributions of day-to-day changes in TIL^(Basic)^. The numbers above each bar represent the number of study patients remaining in the ICU after the corresponding day-to-day step. Percentages which round to 2% or lower are not shown. Abbreviations: ICU=intensive care unit, TIL=Therapy Intensity Level, TIL^(Basic)^=condensed, five-category TIL scale as defined in Table 1, WLST=withdrawal of life-sustaining therapies.

### Reliability and performance of TILTomorrow

With both calibration slopes (Fig. 1c) and smoothed calibration curves (Fig. 1d) across the thresholds of next-day TIL^(Basic)^, we observed that the TILTomorrow modelling strategy achieved sufficient testing set calibration for analysis from day two of ICU stay onwards. The 95% CI of the calibration slope pertaining to prediction of next-day TIL^(Basic)^ > 0 was wider than that of other thresholds but still centred around a well-calibrated slope of one.

In the first week of ICU stay, TILTomorrow correctly discriminated patients at each threshold of next-day TIL^(Basic)^ between 79% (95% CI: 77–82%) and 95% (95% CI: 93– 96%) of the time (Fig. 3a). However, this apparently strong predictive power was in fact largely because TIL^(Basic)^ tended not to change greatly (i.e., the “inertia” of TIL) across day-to-day steps (Fig. 2b), especially at higher thresholds of next-day TIL^(Basic)^ (violet lines in Fig. 3a). After removing all treatments and physician-based impressions from the model variable set (including all variables related to TIL), the first-week AUCs dropped to between 0.65 (95% CI: 0.62–0.68) and 0.86 (95% CI: 0.82–0.89) with significantly lower performance at higher thresholds of next-day TIL^(Basic)^ (Fig. 3a). Models trained with only static variables achieved only marginally better discrimination than an uninformative predictor (best AUC: 0.60 [95% CI: 0.56–0.63], Fig. 3a).

**Fig. 3.**
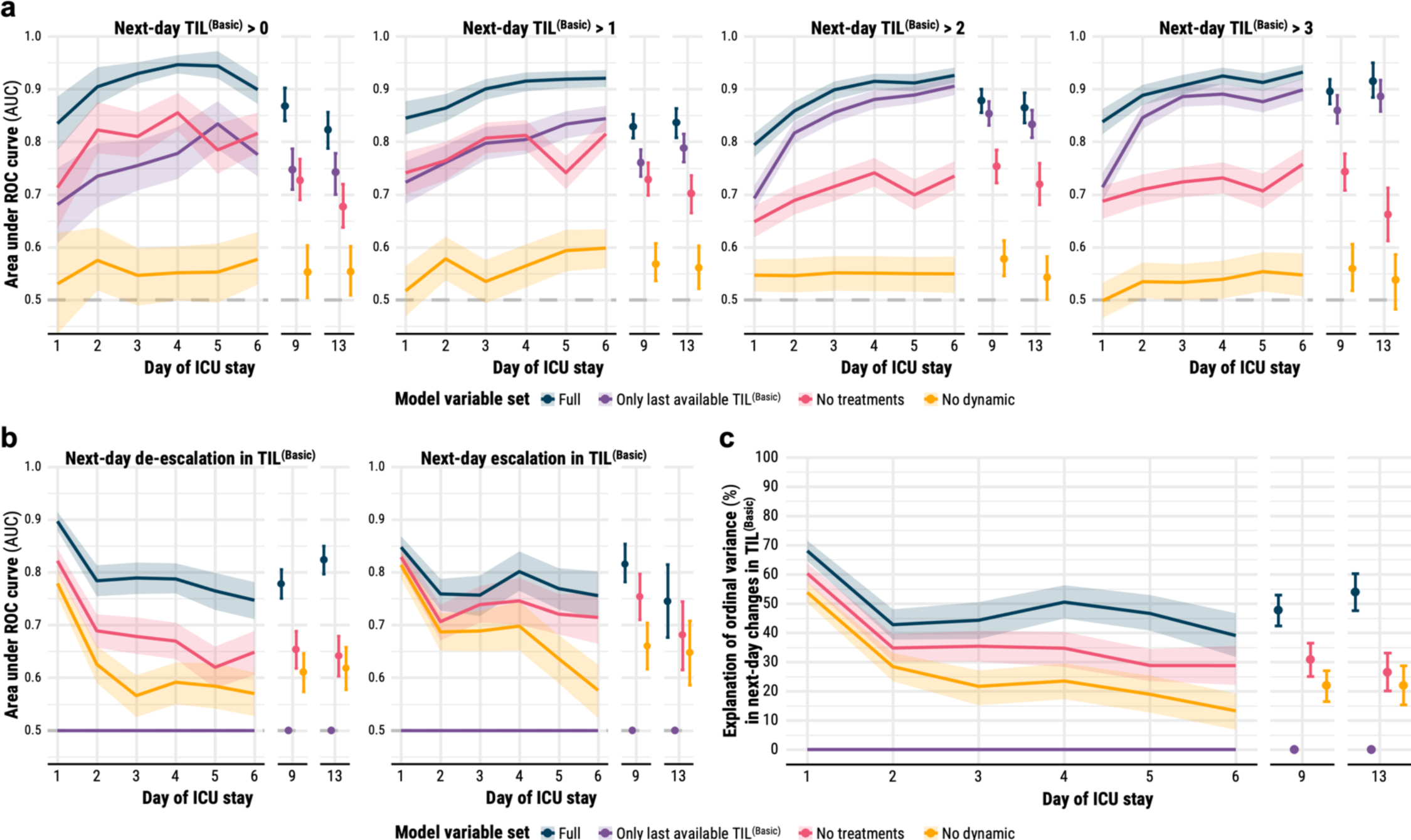
Differential performance in discriminating and explaining next-day TIL^(Basic)^. All shaded regions surrounding curves and error bars are 95% confidence intervals derived using bias-corrected bootstrapping (1,000 resamples) to represent the variation across 20 repeated five-fold cross-validation partitions. (**a**) Discrimination performance in prediction of next-day TIL^(Basic)^ – measured by AUC at each threshold of TIL^(Basic)^ – by models trained on different variable sets. The violet line represents the performance achieved by simply carrying the last available TIL^(Basic)^ forward to account for the effect of day-to-day stasis in TIL^(Basic)^ on prediction. The horizontal dashed line (AUC=0.5) represents the performance of uninformative prediction. (**b**) Discrimination performance in prediction of next-day de-escalation or escalation in TIL^(Basic)^ – measured by AUC – by models trained on different variable sets. The horizontal dashed line (AUC=0.5) represents the performance of uninformative prediction. (**c**) Explanation of ordinal variation in next-day changes in TIL^(Basic)^ – measured by Somers’ *D_xy_* – by models trained on different variable sets. Abbreviations: AUC=area under the receiver operating characteristic (ROC) curve, ICU=intensive care unit, TIL=Therapy Intensity Level, TIL^(Basic)^=condensed, five-category TIL scale as defined in Table 1.

To completely account for the inertia of TIL^(Basic)^ across day-to-day steps, we calculated discrimination performance in the prediction of changes in next-day TIL^(Basic)^ (Fig. 3b). Prediction performance was highest on day one across all variable sets, with the full-variable model correctly discriminating next-day de-escalations 90% (95% CI: 88–91%) of the time and next-day escalations 85% (95% CI: 83–87%) of the time. Within each variable set, change-in-TIL^(Basic)^ prediction performance did not change significantly from day two onwards, except for the prediction of next-day escalation from static variables. Treatment and physician-based impression variables significantly improved performance in prediction of next-day de-escalations in TIL^(Basic)^ but not in prediction of next-day escalations in TIL^(Basic)^ (Fig. 3b). Moreover, static variables achieved greater discrimination in the prediction of TIL^(Basic)^ escalations than in the prediction of TIL^(Basic)^ de-escalations from days two to four of ICU stay.

### Differential explanation of next-day changes in TIL

The full set of 2,008 variables explained 68% (95% CI: 65–72%) of the ordinal variation in next-day changes in TIL^(Basic)^ on day one and up to 51% (95% CI: 45–56%) through the rest of the first week (Fig. 3c). For the same endpoint, the 1,803 variables which exclude treatments and physician-based impressions explained 60% (95% CI: 57–64%) of the ordinal variation on day one and up to 35% (95% CI: 30–41%) thereafter (Fig. 3c). From Fig. 3b, we found that the explanation added from the prior trajectory of ICU management related more to informative patterns of treatment de-escalation than to those of escalation. At the same time, most of the explanation achieved by the full variable model could also be achieved without explicit information about the patient’s treatments. The 1,029 static variables explained 54% (95% CI: 50–57%) of the ordinal variation in next-day changes in TIL^(Basic)^ on day one and decreased in explanation significantly from days two (28% [95% CI: 23–33%]) to six (13% [95% CI: 7–19%]) (Fig. 3c). In other words, the explanatory impact of dynamic variables increased over time in the ICU. Most of the explanatory information in static variables contributed towards prediction of treatment escalations earlier in patients’ ICU stays (Fig. 3b).

### Variables associated with next-day changes in TIL

During the days of consecutive TIL assessment that were eligible for ΔTimeSHAP calculation (days 2–7), 575 patients (68% of population) experienced a total of 1,004 day-to-day changes in TIL^(Basic)^. The associative contributions of highest-impact variables towards prediction of these changes – both for models trained on all variables and for those trained without treatment variables – are visualised in Fig. 4. The number of points for each variable in Fig. 4 equals the number of times each variable was represented across the 1,004 changes in TIL^(Basic)^. Moreover, we annotated several specific values of categorical variables in Fig. 4 because of their visually consistent association with next-day TIL^(Basic)^ de-escalation (i.e., negative ΔTimeSHAP) or TIL^(Basic)^ escalation (i.e., positive ΔTimeSHAP). Across the leading predictors of next-day changes in TIL^(Basic)^ (Fig. 4), we found the following categories of variables:

- the preceding trajectory of ICU management (e.g., extubation, prior trajectory of TIL, ending nasogastric feeding),
- age at admission,
- bleeding risk factors (e.g., history of taking anticoagulants, baseline platelet count),
- brain imaging results (e.g., traumatic subarachnoid haemorrhage, subdural haematoma, intraparenchymal haemorrhage),
- haemodynamics and intracranial hypertension (e.g., ICP, blood pressure, respiratory efficiency),
- markers of systemic inflammation (e.g., ventilator-associated pneumonia [which may also reflect long ventilation time], eosinophils),
- metabolic derangements (e.g., sodium, calcium, alanine aminotransferase),
- neurological function (e.g., Glasgow Coma Scale [GCS] eye and motor scores),
- protein biomarkers (e.g., neurofilament-light chain, total tau protein).

**Fig 4.**
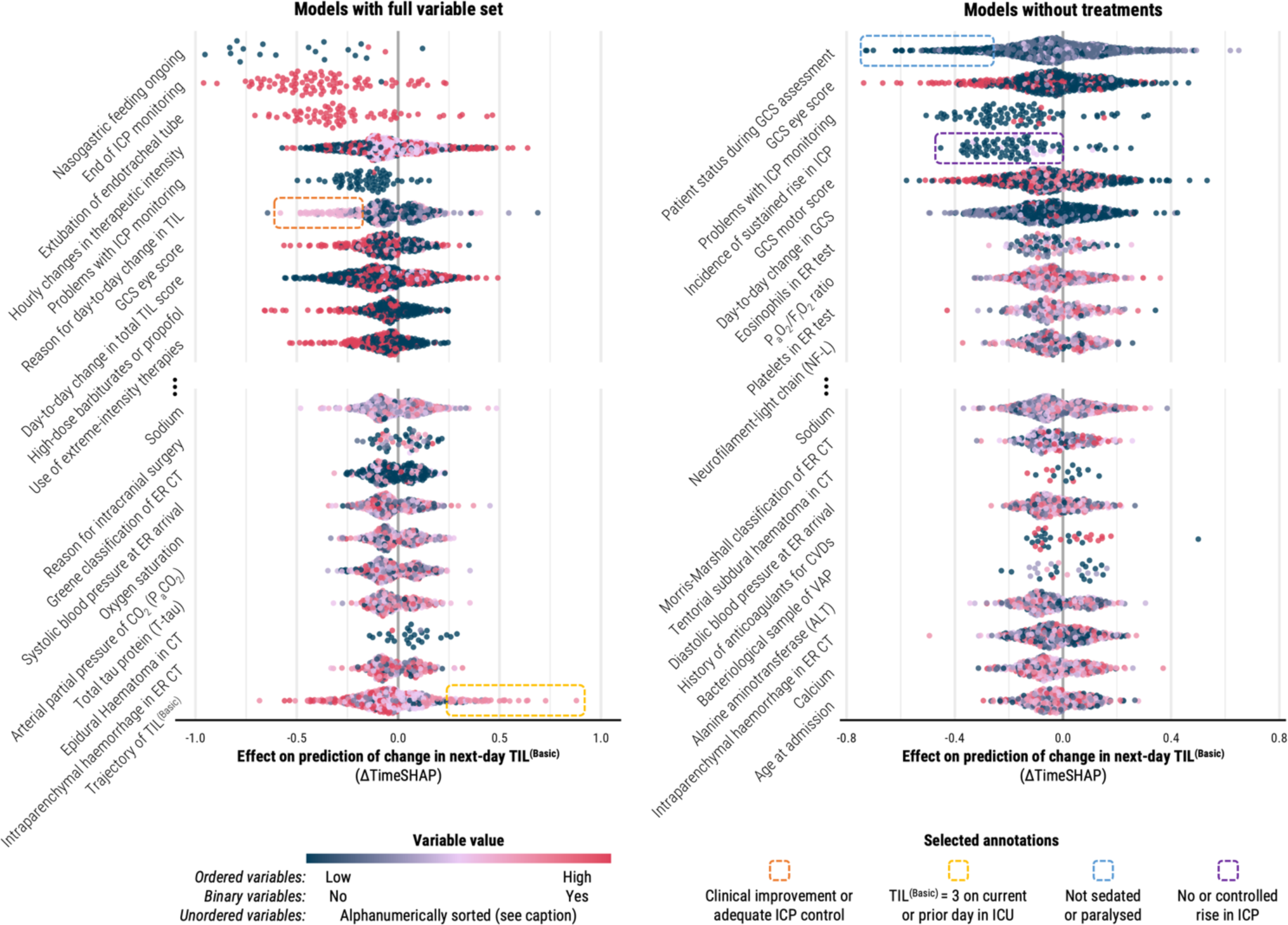
Population-level variable contributions to prediction of changes in next-day TIL^(Basic)^ at days directly preceding a change in TIL^(Basic)^. The ΔTimeSHAP values on the left panel are from the models trained on the full variable set whilst the ΔTimeSHAP values on the right panel are from the models trained without clinician impressions or treatments. ΔTimeSHAP values are interpreted as the relative contributions of variables towards the difference in model prediction of next-day TIL^(Basic)^ over the two days directly preceding the change in TIL^(Basic)^ (Supplementary Methods S5). Therefore, the study population represented in this figure is limited to patients who experienced a change in TIL^(Basic)^ after day two of ICU stay (*n* = 575). A positive ΔTimeSHAP value signifies association with an increased likelihood of escalation in next-day TIL^(Basic)^, whereas a negative ΔTimeSHAP value signifies association with an increased likelihood of de-escalation. The variables were selected by first identifying the ten variables with non-missing value tokens with the most negative median ΔTimeSHAP values across the population (above the ellipses) and then, amongst the remaining variables, selecting the ten with non-missing value tokens with the most positive median ΔTimeSHAP values (below the ellipses). Each point represents the mean ΔTimeSHAP value, taken across all 20 repeated cross-validation partitions, for a token preceding an individual patient’s change in TIL^(Basic)^. The number of points for each variable, therefore, indicates the relative occurrence of that variable before changes in TIL^(Basic)^ in the study population. The colour of the point represents the relative ordered value of a token within a variable, and for unordered variables (e.g., patient status during GCS assessment), tokens were sorted alphanumerically (the sort index per possible unordered variable token is provided in the CENTER-TBI data dictionary: https://www.center-tbi.eu/data/dictionary). Abbreviations: CVDs=cardiovascular diseases, ER=emergency room, F*_I_*O_2_=fraction of inspired oxygen, GCS=Glasgow Coma Scale, ICP=intracranial pressure, P_a_O_2_=partial pressure of oxygen, TIL=Therapy Intensity Level, TIL^(Basic)^=condensed, five-category TIL scale as defined in Table 1, VAP=ventilator-associated pneumonia.

The most robust predictors of next-day de-escalation in TIL^(Basic)^ were other clinical indicators of treatment de-escalation (e.g., ending nasogastric feeding), improvement in patients’ eye-opening responses, previous administration of barbiturates or propofol, and sufficient control of ICP. Overall, the effects of predictors for TIL^(Basic)^ escalation were not as robust as those for de-escalation (Fig. 4); however, stratifying the ΔTimeSHAP values by the pre-transition TIL^(Basic)^ score revealed more consistent associations per level of treatment intensity (Supplementary Fig. S5). For example, high ICP values were robustly predictive of escalations from TIL^(Basic)^=2, and the prior administration of certain therapies could be predictive of a future escalation or de-escalation based on the current TIL^(Basic)^ score (Supplementary Fig. S5). Apart from treatment variables, the factors that contributed the most towards prediction of de-escalation from extreme ICP management (i.e., TIL^(Basic)^=4) were neurological improvements in motor and eye response with sufficiently controlled ICP and high blood oxygen saturation (Supplementary Fig. S5). The ΔTimeSHAP values of missing variables (Supplementary Fig. S6) demonstrated that missingness of a variable (e.g., missing report of daily complications) could have a significant de-escalating associative effect on model output.

### Conceptual model of changes in treatment intensity

We combined the results from the differential explanation of next-day changes in TIL^(Basic)^ (Fig. 3b–c) and the variable contributions towards prediction of these events (Fig. 4) to produce a conceptual model of day-to-day changes in treatment intensity (Fig. 5). Given the considerable difference in explanation performance between day one and subsequent days of ICU stay, we separated these explanation percentages in our model.

**Fig 5.**
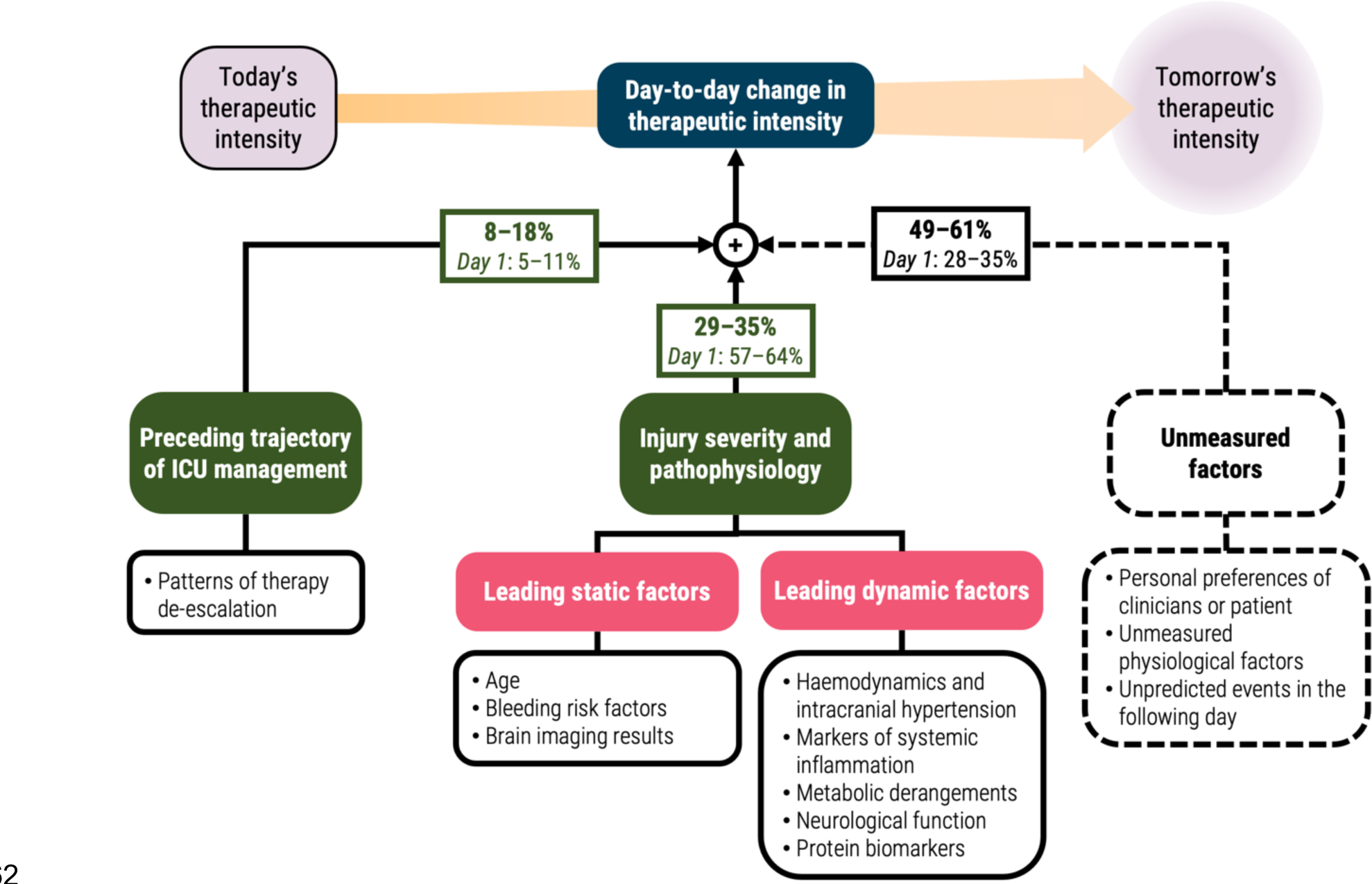
Conceptual diagram of factors explaining day-to-day changes in therapeutic intensity. The percentage values represent the differential explanation of ordinal variation in next-day changes in TIL^(Basic)^ as measured by Somers’ *D_xy_*. The bolded percentage values represent the 95% confidence interval of Somers’ *D_xy_* from days 2–6 of ICU stay, whilst the percentage values below them represent the 95% confidence interval of Somers’ *D_xy_* from day 1 of ICU stay (Fig. 3c). The 95% confidence intervals were derived using bias-corrected bootstrapping (1,000 resamples) to represent the variation across 20 repeated five-fold cross-validation partitions. The leading static and dynamic pathophysiological factors were determined by qualitative categorisation of the variables with the highest contribution to next-day changes in TIL^(Basic)^ based on ΔTimeSHAP values (Fig. 4). Abbreviations: TIL=Therapy Intensity Level, TIL^(Basic)^=condensed, five-category TIL scale as defined in Table 1.

## Discussion

We present the first approach to dynamic prediction of future therapy intensity levels (TIL) in ICP-monitored TBI patients. The TILTomorrow modelling strategy allowed us to exploit the full clinical context (2,008 variables) captured in a large neurotrauma dataset over time to uncover factors associated with next-day changes in TIL^(Basic)^.^19^ By including missing value tokens, models discovered meaningful patterns of missingness (Supplementary Fig. S6).^20^ Moreover, our approach mapped clinical events to evolving predictions at each ordinal level of next-day TIL^(Basic)^, which is an improvement in statistical power and clinical information over using a dichotomised measure of therapeutic intensity (e.g., high-TIL therapies).^15^

The main results of this study are summarised in the conceptual diagram of changes in TIL^(Basic)^ (Fig. 5). Amongst all day-to-day steps, the transition from day one to day two had the greatest number of changes in TIL^(Basic)^ (Fig. 2b), which were also the most predictable (68% [95% CI: 65–72%] explanation, Fig. 3c). From day two onwards, the ordinal explanation of changes in next-day TIL^(Basic)^ dropped to between 39% (95% CI: 32–47%) and 51% (95% CI: 45–56%). This difference suggests that first-to-second-day changes in treatment intensity might have been the most systematic, possibly associated with primary injury severity and initial patient responses to treatment (Fig. 3c). Later in ICU stay, the predictive influence of a patient’s treatment trajectory increased (mostly through informative patterns of de-escalation, Fig. 3b), and that of static factors decreased (Fig. 3c). Whilst static factors are poor predictors of TIL^(Basic)^ on any given day (Fig. 3a and as shown previously^12^), they achieve considerable discrimination performance in prediction of escalations up to day four (AUC: 0.70 [95% CI: 0.65–0.74], Fig. 3b). This may indicate the potential of certain primary injury factors for justifying earlier intervention as to avoid tolerating suboptimal ICP management for a few days. Apart from age, the highest-contributing static factors were space-occupying lesions (also reflected in a recent study^28^) and bleeding risk factors (Fig. 4), both of which can complicate ICP control. As targets of TIL therapies, ICP and haemodynamic factors are expectedly high-contributing, with different effects based on the pre-transition TIL^(Basic)^ score (Supplementary Fig. S5). Metabolic complications (i.e., abnormalities in renal or liver function and electrolytic imbalances) have previously been shown to be significantly more common in patients receiving high-TIL therapies^12^ and an important marker for physiological endotyping.^29^ Moreover, in a prior study, serial protein biomarkers (in addition to GCS) were key descriptors for clustering TBI patient trajectories in the ICU.^30^ Therefore, the results from these dynamic variables support the links between TIL and pathophysiology – including systemic factors (e.g., metabolism and inflammation) – after TBI.^7^ This is potentially of clinical importance since protein biomarkers are not measured serially as part of typical routine care outside of research studies (e.g., CENTER-TBI) and a few centres. It is still uncertain whether serial biomarker measurement would improve care outcomes. However, analysing the temporal dynamics of these biomarkers may not only enable a more precise characterisation of patients’ treatment needs but also elucidate biological mechanisms underpinning variable treatment response. Finally, whilst we found no significant difference in full-model prediction performance between next-day escalations and de-escalations of TIL^(Basic)^ (Fig. 3b), high-impact predictors had a more robust signal with de-escalations than they did with escalations (i.e., more consistently negative ΔTimeSHAP values in Fig. 4). This suggests that escalation prediction may be the effect of a complex interaction of factors which is difficult to perceive with ΔTimeSHAP values.

The underlying assumption of this work is that a more protocolised management of ICP would also be more predictable based on the dynamic condition of a TBI patient. Even with wide inter-centre variation in ICP-targeting treatment,^14^ we would expect the measurable factors which rationally drive day-to-day changes in TIL to predict such changes on an individual level. After day two, approximately half of the ordinal variation in day-to-day changes in TIL^(Basic)^ is unexplained by the full CENTER-TBI variable set, and we propose four reasons for this remaining uncertainty (Fig. 5). First, certain clinical events or complications that could suddenly trigger a (de-)escalation in TIL (e.g., sustained rise in ICP) might not have been predictable from the day before. Second, there are probably important physiological factors, either unmeasured or not included in our variable set, which would have improved TIL prediction. Most notably, high-resolution waveforms of ICP^31^ and arterial blood pressure (ABP) and their derived metrics (e.g., pressure-time dose^32^ and vascular reactivity^33^) are more likely to elucidate ICP management decisions than the bihourly clinician-recorded ICP or CPP values available in our variable set.^34^ Prior analyses of additional physiological modalities – e.g., cerebral microdialysis,^35^ automated pupillometry,^36,37^ and motion sensing^38^ – have also demonstrated independent associations with TIL or other short-term endpoints after TBI. Third, assuming different centres have different protocols for ICP management, there may not have been enough patient representation across the spectrum of TBI severity from each centre for TILTomorrow to learn centre-specific guidelines. Fourth, a part of ICP management may be driven by the personal preferences of clinicians in deviation from general guidelines. At the same time, we recognise that predictability does not guarantee a systematised delivery of care. We therefore investigated differential explanation of (Fig. 3b–c) and specific variable contributions towards (Fig. 4) changes in TIL to bridge prediction performance to a plausible concept of ICP management (Fig. 5).

Our results support the use of TIL as an intermediate outcome after TBI.^7^ Specific categories of pathophysiological variables – both static and dynamic – associate well with changes in TIL (Fig. 4 and 5). Since TIL rates the relative risk and complexity of administered treatments, it is logical to minimise TIL when all other factors are held equal. On the other hand, TIL is also a complicated marker of pathophysiology. Since around half of the ordinal variation in changes in TIL is not explained by measured variables (Fig. 5), we hypothesise that TIL’s sensitivity to pathophysiology is partially confounded by the personal preferences of clinical teams. Nevertheless, TIL was previously shown to be a stronger indicator of refractory intracranial hypertension than ICP itself and, thus, a more suitable intermediate endpoint for TBI management.^7^ Since the full information pertaining to TIL was only date-stamped in CENTER-TBI, the highest resolution at which we could assess TIL^(Basic)^ was once per calendar day (Table 1). However, clinicians were also asked to record qualitatively whether treatment intensity was decreasing or increasing every four hours, and these indications (from the day before a change in TIL^(Basic)^) were amongst the strongest predictors of next-day changes in TIL^(Basic)^ (Fig. 4). This result supports a higher resolution TIL for monitoring pathophysiological severity; however, daily TIL scores have been shown to be reliable estimates of hourly TIL scores,^9^ and CENTER-TBI has demonstrated the practical feasibility of daily TIL assessment for a large-scale study (≤2.4% missingness, Fig. 2a).

TILTomorrow can potentially be useful in other heterogeneous-data-intensive clinical domains as a framework for decoding factors tied to treatment decision-making or other dynamic endpoints. This can inform the design of future causal inference models of individualised treatment effects from observational data.^39^ TILTomorrow was not conceived for clinical deployment and should not be used for real-time decision support due to concerns of self-fulfilling prophecies, generalisability, and variable robustness.^40^ Our focus was on explanatory modelling, to derive insightful patterns from the CENTER-TBI data and quantify the predictability of ICP management. Furthermore, ΔTimeSHAP values on observational data are merely associative and cannot be interpreted for causal inference. We used TimeSHAP in this work to highlight potential areas of investigation from a wider, data-driven approach. Pathophysiological predictors of the need for higher TIL (Fig. 4 and 5) could be useful for improving the timing and precision of future clinical decision-making (e.g., performing decompressive craniectomy in a timely but targeted way) but would require more evidence and feasibility studies than just their predictive power in our data.

We recognise several additional limitations in this study. TILTomorrow discretised both numerical variables into binned tokens and time into daily windows, which caused some loss of information. Limited by the resolution of available TIL assessments, we chose a daily time window to avoid inconsistent lead times in our prediction task (Fig. 1a). The highest resolution of regularly recorded variables (e.g., ICP) in the CENTER-TBI core study is once every two hours,^13^ and, since TILTomorrow takes the unique set of tokens per daily window prior to embedding, these numerical variables would be reduced to the unique set of quantiles represented in a day (Fig. 1b). An encoding strategy which can integrate high-resolution ICP, CPP, and other clinical information into broader time windows may improve prediction performance. Additionally, the daily TIL^(Basic)^ score accounts for 33% of the information in the full, 38-point TIL score.^7^ As explained in the Methods, we used TIL^(Basic)^ as the model endpoint over the full TIL score since it would enable us to uncover factors associated with changes across specific, interpretable bands of treatment intensity (Table 1). Nevertheless, a regression-based prediction of next-day full TIL may capture more nuanced patterns of factors associated with changes in ICP management. Finally, our results may encode recruitment, collection, and clinical biases native to our European patient set. Selective recording of clinical data – with selective missingness – may have biased our analyses, and findings may not generalise to other populations.^41^ Given the broad inter-centre variation in ICP-targeted care,^14^ the results of TILTomorrow are likely to vary considerably depending on the protocols of specific centres. We encourage investigators to apply the TILTomorrow approach to other longitudinal, granular ICU datasets of TBI patients – particularly in low- and middle-income countries where the burden of TBI is disproportionately higher^42^ – and compare their results.

## Data and code availability

Individual participant data, including data dictionary, the study protocol, and analysis scripts are available online, conditional to approved study proposal, with no end date. Interested investigators must submit a study proposal to the management committee at https://www.center-tbi.eu/data. Signed confirmation of a data access agreement is required, and all access must comply with regulatory restrictions imposed on the original study.

All code used in this project can be found at the following online repository: https://github.com/sbhattacharyay/TILTomorrow (DOI: 10.5281/zenodo.11060742).

## Acknowledgments

This research was supported by the National Institute for Health Research (NIHR) Brain Injury MedTech Co-operative. CENTER-TBI was supported by the European Union 7^th^ Framework programme (EC grant 602150). Additional funding was obtained from the Hannelore Kohl Stiftung (Germany), from OneMind (USA), from Integra LifeSciences Corporation (USA), and from NeuroTrauma Sciences (USA). CENTER-TBI also acknowledges interactions and support from the International Initiative for TBI Research (InTBIR) investigators. S.B. is funded by a Gates Cambridge Scholarship. E.B. is funded by the Medical Research Council (MR N013433-1) and by a Gates Cambridge Scholarship. The funders had no role in study design, data collection and analysis, decision to publish, or preparation of the manuscript.

We are grateful to the patients and families of our study for making our efforts to improve TBI care possible. S.B. would like to thank Kathleen Mitchell-Fox (Princeton University) for offering comments on the manuscript.

## The CENTER-TBI investigators and participants

The co-lead investigators of CENTER-TBI are designated with an asterisk (*), and their contact email addresses are listed below.

Cecilia Åkerlund^1^, Krisztina Amrein^2^, Nada Andelic^3^, Lasse Andreassen^4^, Audny Anke^5^, Anna Antoni^6^, Gérard Audibert^7^, Philippe Azouvi^8^, Maria Luisa Azzolini^9^, Ronald Bartels^10^, Pál Barzó^11^, Romuald Beauvais^12^, Ronny Beer^13^, Bo-Michael Bellander^14^, Antonio Belli^15^, Habib Benali^16^, Maurizio Berardino^17^, Luigi Beretta^9^, Morten Blaabjerg^18^, Peter Bragge^19^, Alexandra Brazinova^20^, Vibeke Brinck^21^, Joanne Brooker^22^, Camilla Brorsson^23^, Andras Buki^24^, Monika Bullinger^25^, Manuel Cabeleira^26^, Alessio Caccioppola^27^, Emiliana Calappi^27^, Maria Rosa Calvi^9^, Peter Cameron^28^, Guillermo Carbayo Lozano^29^, Marco Carbonara^27^, Simona Cavallo^17^, Giorgio Chevallard^30^, Arturo Chieregato^30^, Giuseppe Citerio^31,32^, Hans Clusmann^33^, Mark Coburn^34^, Jonathan Coles^35^, Jamie D. Cooper^36^, Marta Correia^37^, Amra Čović^38^, Nicola Curry^39^, Endre Czeiter^24^, Marek Czosnyka^26^, Claire Dahyot-Fizelier^40^, Paul Dark^41^, Helen Dawes^42^, Véronique De Keyser^43^, Vincent Degos^16^, Francesco Della Corte^44^, Hugo den Boogert^10^, Bart Depreitere^45^, Đula Đilvesi^46^, Abhishek Dixit^47^, Emma Donoghue^22^, Jens Dreier^48^, Guy-Loup Dulière^49^, Ari Ercole^47^, Patrick Esser^42^, Erzsébet Ezer^50^, Martin Fabricius^51^, Valery L. Feigin^52^, Kelly Foks^53^, Shirin Frisvold^54^, Alex Furmanov^55^, Pablo Gagliardo^56^, Damien Galanaud^16^, Dashiell Gantner^28^, Guoyi Gao^57^, Pradeep George^58^, Alexandre Ghuysen^59^, Lelde Giga^60^, Ben Glocker^61^, Jagoš Golubovic^46^, Pedro A. Gomez^62^, Johannes Gratz^63^, Benjamin Gravesteijn^64^, Francesca Grossi^44^, Russell L. Gruen^65^, Deepak Gupta^66^, Juanita A. Haagsma^64^, Iain Haitsma^67^, Raimund Helbok^13^, Eirik Helseth^68^, Lindsay Horton^69^, Jilske Huijben^64^, Peter J. Hutchinson^70^, Bram Jacobs^71^, Stefan Jankowski^72^, Mike Jarrett^21^, Ji-yao Jiang^58^, Faye Johnson^73^, Kelly Jones^52^, Mladen Karan^46^, Angelos G. Kolias^70^, Erwin Kompanje^74^, Daniel Kondziella^51^, Evgenios Kornaropoulos^47^, Lars-Owe Koskinen^75^, Noémi Kovács^76^, Ana Kowark^77^, Alfonso Lagares^62^, Linda Lanyon^58^, Steven Laureys^78^, Fiona Lecky^79,80^, Didier Ledoux^78^, Rolf Lefering^81^, Valerie Legrand^82^, Aurelie Lejeune^83^, Leon Levi^84^, Roger Lightfoot^85^, Hester Lingsma^64^, Andrew I.R. Maas^43,86,^*, Ana M. Castaño-León^62^, Marc Maegele^87^, Marek Majdan^20^, Alex Manara^88^, Geoffrey Manley^89^, Costanza Martino^90^, Hugues Maréchal^49^, Julia Mattern^91^, Catherine McMahon^92^, Béla Melegh^93^, David Menon^47,^*, Tomas Menovsky^43,86^, Ana Mikolic^64^, Benoit Misset^78^, Visakh Muraleedharan^58^, Lynnette Murray^28^, Ancuta Negru^94^, David Nelson^1^, Virginia Newcombe^47^, Daan Nieboer^64^, József Nyirádi^2^, Otesile Olubukola^79^, Matej Oresic^95^, Fabrizio Ortolano^27^, Aarno Palotie^96,97,98^, Paul M. Parizel^99^, Jean-François Payen^100^, Natascha Perera^12^, Vincent Perlbarg^16^, Paolo Persona^101^, Wilco Peul^102^, Anna Piippo-Karjalainen^103^, Matti Pirinen^96^, Dana Pisica^64^, Horia Ples^94^, Suzanne Polinder^64^, Inigo Pomposo^29^, Jussi P. Posti^104^, Louis Puybasset^105^, Andreea Radoi^106^, Arminas Ragauskas^107^, Rahul Raj^103^, Malinka Rambadagalla^108^, Isabel Retel Helmrich^64^, Jonathan Rhodes^109^, Sylvia Richardson^110^, Sophie Richter^47^, Samuli Ripatti^96^, Saulius Rocka^107^, Cecilie Roe^111^, Olav Roise^112,113^, Jonathan Rosand^114^, Jeffrey V. Rosenfeld^115^, Christina Rosenlund^116^, Guy Rosenthal^55^, Rolf Rossaint^77^, Sandra Rossi^101^, Daniel Rueckert^61^ Martin Rusnák^117^, Juan Sahuquillo^106^, Oliver Sakowitz^91,118^, Renan Sanchez-Porras^118^, Janos Sandor^119^, Nadine Schäfer^81^, Silke Schmidt^120^, Herbert Schoechl^121^, Guus Schoonman^122^, Rico Frederik Schou^123^, Elisabeth Schwendenwein^6^, Charlie Sewalt^64^, Ranjit D. Singh^102^, Toril Skandsen^124,125^, Peter Smielewski^26^, Abayomi Sorinola^126^, Emmanuel Stamatakis^47^, Simon Stanworth^39^, Robert Stevens^127^, William Stewart^128^, Ewout W. Steyerberg^64,129^, Nino Stocchetti^130^, Nina Sundström^131^, Riikka Takala^132^, Viktória Tamás^126^, Tomas Tamosuitis^133^, Mark Steven Taylor^20^, Aurore Thibaut^78^, Braden Te Ao^52^, Olli Tenovuo^104^, Alice Theadom^52^, Matt Thomas^88^, Dick Tibboel^134^, Marjolein Timmers^74^, Christos Tolias^135^, Tony Trapani^28^, Cristina Maria Tudora^94^, Andreas Unterberg^91^, Peter Vajkoczy^136^, Shirley Vallance^28^, Egils Valeinis^60^, Zoltán Vámos^50^, Mathieu van der Jagt^137^, Gregory Van der Steen^43^, Joukje van der Naalt^71^, Jeroen T.J.M. van Dijck^102^, Inge A. M. van Erp^102^, Thomas A. van Essen^102^, Wim Van Hecke^138^, Caroline van Heugten^139^, Ernest van Veen^64^, Thijs Vande Vyvere^140^, Roel P. J. van Wijk^102^, Alessia Vargiolu^32^, Emmanuel Vega^83^, Kimberley Velt^64^, Jan Verheyden^138^, Paul M. Vespa^141^, Anne Vik^124,142^, Rimantas Vilcinis^133^, Victor Volovici^67^, Nicole von Steinbüchel^38^, Daphne Voormolen^64^, Petar Vulekovic^46^, Kevin K.W. Wang^143^, Daniel Whitehouse^47^, Eveline Wiegers^64^, Guy Williams^47^, Lindsay Wilson^69^, Stefan Winzeck^47^, Stefan Wolf^144^, Zhihui Yang^114^, Peter Ylén^145^, Alexander Younsi^91^, Frederick A. Zeiler^47,146^, Veronika Zelinkova^20^, Agate Ziverte^60^, Tommaso Zoerle^27^

^1^Department of Physiology and Pharmacology, Section of Perioperative Medicine and Intensive Care, Karolinska Institutet, Stockholm, Sweden

^2^János Szentágothai Research Centre, University of Pécs, Pécs, Hungary

^3^Division of Clinical Neuroscience, Department of Physical Medicine and Rehabilitation, Oslo University Hospital and University of Oslo, Oslo, Norway

^4^Department of Neurosurgery, University Hospital Northern Norway, Tromso, Norway

^5^Department of Physical Medicine and Rehabilitation, University Hospital Northern Norway, Tromso, Norway

^6^Trauma Surgery, Medical University Vienna, Vienna, Austria

^7^Department of Anesthesiology & Intensive Care, University Hospital Nancy, Nancy, France

^8^Raymond Poincare hospital, Assistance Publique – Hopitaux de Paris, Paris, France

^9^Department of Anesthesiology & Intensive Care, S Raffaele University Hospital, Milan, Italy

^10^Department of Neurosurgery, Radboud University Medical Center, Nijmegen, The Netherlands

^11^Department of Neurosurgery, University of Szeged, Szeged, Hungary

^12^International Projects Management, ARTTIC, Munchen, Germany

^13^Department of Neurology, Neurological Intensive Care Unit, Medical University of Innsbruck, Innsbruck, Austria

^14^Department of Neurosurgery & Anesthesia & intensive care medicine, Karolinska University Hospital, Stockholm, Sweden

^15^NIHR Surgical Reconstruction and Microbiology Research Centre, Birmingham, UK

^16^Anesthesie-Réanimation, Assistance Publique – Hopitaux de Paris, Paris, France

^17^Department of Anesthesia & ICU, AOU Città della Salute e della Scienza di Torino -

Orthopedic and Trauma Center, Torino, Italy

^18^Department of Neurology, Odense University Hospital, Odense, Denmark

^19^BehaviourWorks Australia, Monash Sustainability Institute, Monash University, Victoria, Australia

^20^Department of Public Health, Faculty of Health Sciences and Social Work, Trnava

University, Trnava, Slovakia

^21^Quesgen Systems Inc., Burlingame, California, USA

^22^Australian & New Zealand Intensive Care Research Centre, Department of Epidemiology and Preventive Medicine, School of Public Health and Preventive Medicine, Monash University, Melbourne, Australia

^23^Department of Surgery and Perioperative Science, Umeå University, Umeå, Sweden

^24^Department of Neurosurgery, Medical School, University of Pécs, Hungary and Neurotrauma Research Group, János Szentágothai Research Centre, University of Pécs, Hungary

^25^Department of Medical Psychology, Universitätsklinikum Hamburg-Eppendorf, Hamburg, Germany

^26^Brain Physics Lab, Division of Neurosurgery, Dept of Clinical Neurosciences, University of Cambridge, Addenbrooke’s Hospital, Cambridge, UK

^27^Neuro ICU, Fondazione IRCCS Cà Granda Ospedale Maggiore Policlinico, Milan, Italy

^28^ANZIC Research Centre, Monash University, Department of Epidemiology and Preventive Medicine, Melbourne, Victoria, Australia

^29^Department of Neurosurgery, Hospital of Cruces, Bilbao, Spain

^30^NeuroIntensive Care, Niguarda Hospital, Milan, Italy

^31^School of Medicine and Surgery, Università Milano Bicocca, Milano, Italy

^32^NeuroIntensive Care Unit, Department Neuroscience, IRCCS Fondazione San Gerardo dei Tintori, Monza, Italy

^33^Department of Neurosurgery, Medical Faculty RWTH Aachen University, Aachen,

Germany

^34^Department of Anesthesiology and Intensive Care Medicine, University Hospital Bonn, Bonn, Germany

^35^Department of Anesthesia & Neurointensive Care, Cambridge University Hospital NHS Foundation Trust, Cambridge, UK

^36^School of Public Health & PM, Monash University and The Alfred Hospital, Melbourne, Victoria, Australia

^37^Radiology/MRI department, MRC Cognition and Brain Sciences Unit, Cambridge, UK

^38^Institute of Medical Psychology and Medical Sociology, Universitätsmedizin Göttingen, Göttingen, Germany

^39^Oxford University Hospitals NHS Trust, Oxford, UK

^40^Intensive Care Unit, CHU Poitiers, Potiers, France

^41^University of Manchester NIHR Biomedical Research Centre, Critical Care Directorate, Salford Royal Hospital NHS Foundation Trust, Salford, UK

^42^Movement Science Group, Faculty of Health and Life Sciences, Oxford Brookes University, Oxford, UK

^43^Department of Neurosurgery, Antwerp University Hospital, Edegem, Belgium

^44^Department of Anesthesia & Intensive Care, Maggiore Della Carità Hospital, Novara, Italy

^45^Department of Neurosurgery, University Hospitals Leuven, Leuven, Belgium

^46^Department of Neurosurgery, Clinical centre of Vojvodina, Faculty of Medicine, University of Novi Sad, Novi Sad, Serbia

^47^Division of Anaesthesia, University of Cambridge, Addenbrooke’s Hospital, Cambridge, UK

^48^Center for Stroke Research Berlin, Charité – Universitätsmedizin Berlin, corporate member of Freie Universität Berlin, Humboldt-Universität zu Berlin, and Berlin Institute of Health, Berlin, Germany

^49^Intensive Care Unit, CHR Citadelle, Liège, Belgium

^50^Department of Anaesthesiology and Intensive Therapy, University of Pécs, Pécs, Hungary

^51^Departments of Neurology, Clinical Neurophysiology and Neuroanesthesiology, Region Hovedstaden Rigshospitalet, Copenhagen, Denmark

^52^National Institute for Stroke and Applied Neurosciences, Faculty of Health and Environmental Studies, Auckland University of Technology, Auckland, New Zealand

^53^Department of Neurology, Erasmus MC, Rotterdam, the Netherlands

^54^Department of Anesthesiology and Intensive care, University Hospital Northern Norway, Tromso, Norway

^55^Department of Neurosurgery, Hadassah-hebrew University Medical center, Jerusalem,

Israel

^56^Fundación Instituto Valenciano de Neurorrehabilitación (FIVAN), Valencia, Spain

^57^Department of Neurosurgery, Shanghai Renji hospital, Shanghai Jiaotong University/school of medicine, Shanghai, China

^58^Karolinska Institutet, INCF International Neuroinformatics Coordinating Facility, Stockholm, Sweden

^59^Emergency Department, CHU, Liège, Belgium

^60^Neurosurgery clinic, Pauls Stradins Clinical University Hospital, Riga, Latvia

^61^Department of Computing, Imperial College London, London, UK

^62^Department of Neurosurgery, Hospital Universitario 12 de Octubre, Madrid, Spain

^63^Department of Anesthesia, Critical Care and Pain Medicine, Medical University of Vienna, Austria

^64^Department of Public Health, Erasmus Medical Center-University Medical Center, Rotterdam, The Netherlands

^65^College of Health and Medicine, Australian National University, Canberra, Australia

^66^Department of Neurosurgery, Neurosciences Centre & JPN Apex trauma centre, All India Institute of Medical Sciences, New Delhi-110029, India

^67^Department of Neurosurgery, Erasmus MC, Rotterdam, the Netherlands

^68^Department of Neurosurgery, Oslo University Hospital, Oslo, Norway

^69^Division of Psychology, University of Stirling, Stirling, UK

^70^Division of Neurosurgery, Department of Clinical Neurosciences, Addenbrooke’s Hospital & University of Cambridge, Cambridge, UK

^71^Department of Neurology, University of Groningen, University Medical Center

Groningen, Groningen, Netherlands

^72^Neurointensive Care, Sheffield Teaching Hospitals NHS Foundation Trust, Sheffield, UK

^73^Salford Royal Hospital NHS Foundation Trust Acute Research Delivery Team, Salford, UK

^74^Department of Intensive Care and Department of Ethics and Philosophy of Medicine, Erasmus Medical Center, Rotterdam, The Netherlands

^75^Department of Clinical Neuroscience, Neurosurgery, Umeå University, Umeå, Sweden

^76^Hungarian Brain Research Program - Grant No. KTIA_13_NAP-A-II/8, University of Pécs, Pécs, Hungary

^77^Department of Anaesthesiology, University Hospital of Aachen, Aachen, Germany

^78^Cyclotron Research Center, University of Liège, Liège, Belgium

^79^Centre for Urgent and Emergency Care Research (CURE), Health Services Research Section, School of Health and Related Research (ScHARR), University of Sheffield, Sheffield, UK

^80^Emergency Department, Salford Royal Hospital, Salford UK

^81^Institute of Research in Operative Medicine (IFOM), Witten/Herdecke University, Cologne, Germany

^82^VP Global Project Management CNS, ICON, Paris, France

^83^Department of Anesthesiology-Intensive Care, Lille University Hospital, Lille, France

^84^Department of Neurosurgery, Rambam Medical Center, Haifa, Israel

^85^Department of Anesthesiology & Intensive Care, University Hospitals Southhampton NHS Trust, Southhampton, UK

^86^Department of Translational Neuroscience, Faculty of Medicine and Health Science,

University of Antwerp, Antwerp, Belgium

^87^Cologne-Merheim Medical Center (CMMC), Department of Traumatology, Orthopedic Surgery and Sportmedicine, Witten/Herdecke University, Cologne, Germany

^88^Intensive Care Unit, Southmead Hospital, Bristol, Bristol, UK

^89^Department of Neurological Surgery, University of California, San Francisco, California, USA

^90^Department of Anesthesia & Intensive Care,M. Bufalini Hospital, Cesena, Italy

^91^Department of Neurosurgery, University Hospital Heidelberg, Heidelberg, Germany

^92^Department of Neurosurgery, The Walton centre NHS Foundation Trust, Liverpool, UK

^93^Department of Medical Genetics, University of Pécs, Pécs, Hungary

^94^Department of Neurosurgery, Emergency County Hospital Timisoara, Timisoara, Romania

^95^School of Medical Sciences, Örebro University, Örebro, Sweden

^96^Institute for Molecular Medicine Finland, University of Helsinki, Helsinki, Finland

^97^Analytic and Translational Genetics Unit, Department of Medicine; Psychiatric & Neurodevelopmental Genetics Unit, Department of Psychiatry; Department of Neurology, Massachusetts General Hospital, Boston, MA, USA

^98^Program in Medical and Population Genetics; The Stanley Center for Psychiatric Research, The Broad Institute of MIT and Harvard, Cambridge, MA, USA

^99^Department of Radiology, University of Antwerp, Edegem, Belgium

^100^Department of Anesthesiology & Intensive Care, University Hospital of Grenoble, Grenoble, France

^101^Department of Anesthesia & Intensive Care, Azienda Ospedaliera Università di Padova, Padova, Italy

^102^Dept. of Neurosurgery, Leiden University Medical Center, Leiden, The Netherlands and Dept. of Neurosurgery, Medical Center Haaglanden, The Hague, The Netherlands

^103^Department of Neurosurgery, Helsinki University Central Hospital

^104^Division of Clinical Neurosciences, Department of Neurosurgery and Turku Brain Injury Centre, Turku University Hospital and University of Turku, Turku, Finland

^105^Department of Anesthesiology and Critical Care, Pitié -Salpêtrière Teaching Hospital, Assistance Publique, Hôpitaux de Paris and University Pierre et Marie Curie, Paris, France

^106^Neurotraumatology and Neurosurgery Research Unit (UNINN), Vall d’Hebron Research Institute, Barcelona, Spain

^107^Department of Neurosurgery, Kaunas University of technology and Vilnius University, Vilnius, Lithuania

^108^Department of Neurosurgery, Rezekne Hospital, Latvia

^109^Department of Anaesthesia, Critical Care & Pain Medicine NHS Lothian & University of Edinburg, Edinburgh, UK

^110^Director, MRC Biostatistics Unit, Cambridge Institute of Public Health, Cambridge, UK

^111^Department of Physical Medicine and Rehabilitation, Oslo University Hospital/University of Oslo, Oslo, Norway

^112^Division of Orthopedics, Oslo University Hospital, Oslo, Norway

^113^Institue of Clinical Medicine, Faculty of Medicine, University of Oslo, Oslo, Norway

^114^Broad Institute, Cambridge MA Harvard Medical School, Boston MA, Massachusetts General Hospital, Boston MA, USA

^115^National Trauma Research Institute, The Alfred Hospital, Monash University, Melbourne, Victoria, Australia

^116^Department of Neurosurgery, Odense University Hospital, Odense, Denmark

^117^International Neurotrauma Research Organisation, Vienna, Austria

^118^Klinik für Neurochirurgie, Klinikum Ludwigsburg, Ludwigsburg, Germany

^119^Division of Biostatistics and Epidemiology, Department of Preventive Medicine, University of Debrecen, Debrecen, Hungary

^120^Department Health and Prevention, University Greifswald, Greifswald, Germany

^121^Department of Anaesthesiology and Intensive Care, AUVA Trauma Hospital, Salzburg, Austria

^122^Department of Neurology, Elisabeth-TweeSteden Ziekenhuis, Tilburg, the Netherlands

^123^Department of Neuroanesthesia and Neurointensive Care, Odense University Hospital, Odense, Denmark

^124^Department of Neuromedicine and Movement Science, Norwegian University of Science and Technology, NTNU, Trondheim, Norway

^125^Department of Physical Medicine and Rehabilitation, St.Olavs Hospital, Trondheim University Hospital, Trondheim, Norway

^126^Department of Neurosurgery, University of Pécs, Pécs, Hungary

^127^Division of Neuroscience Critical Care, John Hopkins University School of Medicine, Baltimore, USA

^128^Department of Neuropathology, Queen Elizabeth University Hospital and University of

Glasgow, Glasgow, UK

^129^Dept. of Department of Biomedical Data Sciences, Leiden University Medical Center, Leiden, The Netherlands

^130^Department of Pathophysiology and Transplantation, Milan University, and Neuroscience ICU, Fondazione IRCCS Cà Granda Ospedale Maggiore Policlinico, Milano, Italy

^131^Department of Radiation Sciences, Biomedical Engineering, Umeå University, Umeå, Sweden

^132^Perioperative Services, Intensive Care Medicine and Pain Management, Turku University Hospital and University of Turku, Turku, Finland

^133^Department of Neurosurgery, Kaunas University of Health Sciences, Kaunas, Lithuania

^134^Intensive Care and Department of Pediatric Surgery, Erasmus Medical Center, Sophia Children’s Hospital, Rotterdam, The Netherlands

^135^Department of Neurosurgery, Kings college London, London, UK

^136^Neurologie, Neurochirurgie und Psychiatrie, Charité – Universitätsmedizin Berlin, Berlin, Germany

^137^Department of Intensive Care Adults, Erasmus MC– University Medical Center Rotterdam, Rotterdam, the Netherlands

^138^icoMetrix NV, Leuven, Belgium

^139^Movement Science Group, Faculty of Health and Life Sciences, Oxford Brookes University, Oxford, UK

^140^Radiology Department, Antwerp University Hospital, Edegem, Belgium

^141^Director of Neurocritical Care, University of California, Los Angeles, USA

^142^Department of Neurosurgery, St.Olavs Hospital, Trondheim University Hospital, Trondheim, Norway

^143^Department of Emergency Medicine, University of Florida, Gainesville, Florida, USA

^144^Department of Neurosurgery, Charité – Universitätsmedizin Berlin, corporate member of Freie Universität Berlin, Humboldt-Universität zu Berlin, and Berlin Institute of Health, Berlin, Germany

^145^VTT Technical Research Centre, Tampere, Finland

^146^Section of Neurosurgery, Department of Surgery, Rady Faculty of Health Sciences, University of Manitoba, Winnipeg, MB, Canada

*Co-lead investigators: (AIRM) and (DM)

