## Supplementary for "TILTomorrow today: dynamic factors predicting changes in intracranial pressure treatment intensity after traumatic brain injury"

##### TABLE OF CONTENTS

**Supplementary Figures** ... **2**

*Supplementary Fig. S1. Flow diagram for patient enrolment.* ... *2*

*Supplementary Fig. S2. Distributions of $TIL^{(Basic)}$ in the study population over days of ICU stay.* ... *2*

*Supplementary Fig. S3. Distributions of $TIL^{(Basic)}$ in the study population stratified by previous $TIL^{(Basic)}$ score.* ... *3*

*Supplementary Fig. S4. Distributions of $TIL^{(Basic)}$ directly preceding/following a change in $TIL^{(Basic)}$.* ... *4*

*Supplementary Fig. S5. Population-level ΔTimeSHAP values stratified by pre-transition $TIL^{(Basic)}$ score.* ... *5*

*Supplementary Fig. S6. Population-level ΔTimeSHAP values for missing value tokens.* ... *8*

**Supplementary Tables** ... **9**

*Supplementary Table S1. Manually excluded variables indicating death or withdrawal of life-sustaining treatment.* ... *9*

*Supplementary Table S2. Physician-based impression variables.* ... *13*

**Supplementary Methods** ... **17**

*Supplementary Methods S1. Description of model endpoints and outputs for TILTomorrow.* ... *17*
Post-processing calibration ... 17

*Supplementary Methods S2. Repeated Bootstrap Bias Corrected with Dropping Cross-Validation (BBCD-CV).* ... *19*
Dropout of low-performing hyperparametric configurations ... 19
Confidence intervals for testing set performance ... 19

*Supplementary Methods S3. Hyperparameter optimisation report.* ... *20*
Summary ... 20
Overview of tested hyperparameters ... 20
Tested hyperparameters per model version ... 20

*Supplementary Methods S4. Calculation of Somers' $D_{xy}$.* ... *25*

*Supplementary Methods S5. Explanation of model outputs with Shapley value estimations.* ... *27*
Shapley values ... 27
KernelSHAP ... 27
TimeSHAP and ΔTimeSHAP ... 28

**Supplementary References** ... **30**

#### SUPPLEMENTARY FIGURES

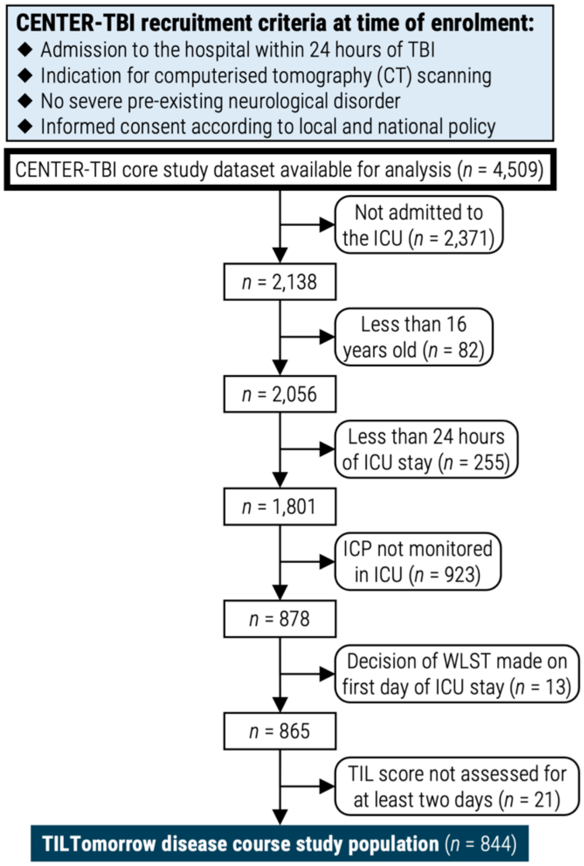


**Supplementary Fig. S1. Flow diagram for patient enrolment.** Abbreviations: CENTER-TBI=Collaborative European NeuroTrauma Effectiveness Research in TBI, ICP=intracranial pressure, ICU=intensive care unit, TBI=traumatic brain injury, TIL=Therapy Intensity Level scale, WLST=withdrawal of life-sustaining therapies.

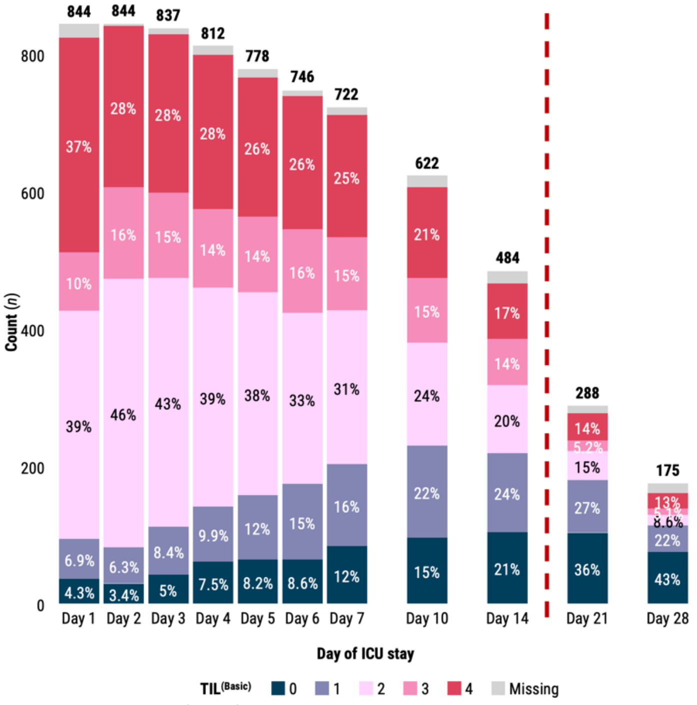


**Supplementary Fig. S2. Distributions of TIL$^{(Basic)}$ in the study population over days of ICU stay.** Percentages are calculated out of the number of study patients remaining in the ICU at the corresponding day (written above each bar), and percentages which round to 2% or lower are not shown. The days of ICU stay before the vertical, dashed red line were used for assessment of the TILTomorrow modelling strategy. Abbreviations: ICU=intensive care unit, TIL=Therapy Intensity Level, TIL$^{(Basic)}$=condensed, five-category TIL scale as defined in Table 1.

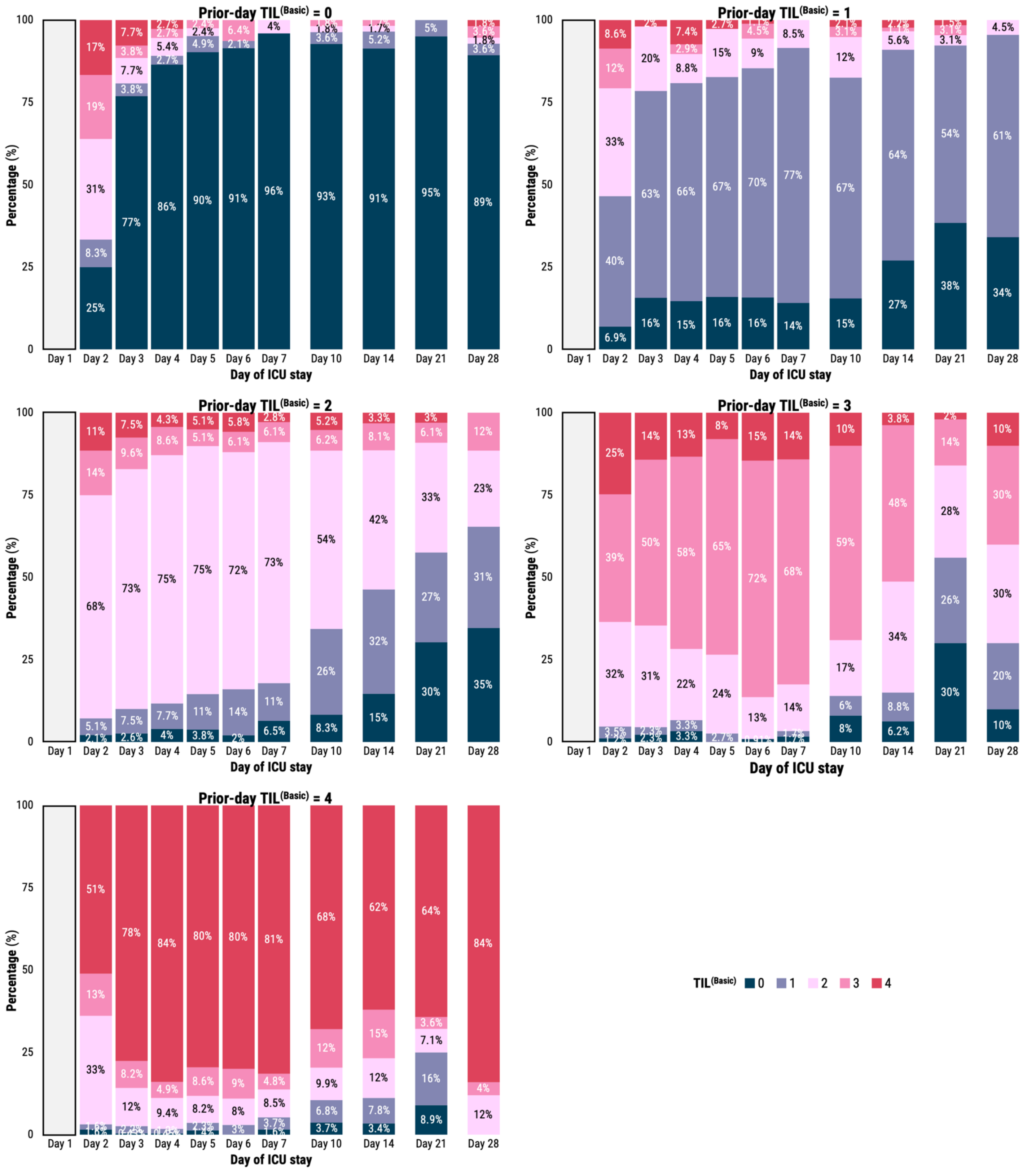


**Supplementary Fig. S3. Distributions of TIL$^{(Basic)}$ in the study population stratified by previous TIL$^{(Basic)}$ score.** Percentages are calculated out of the number of study patients remaining in the ICU at the corresponding day whose prior-day TIL$^{(Basic)}$ score equalled the score above the panel. Abbreviations: ICU=intensive care unit, TIL=Therapy Intensity Level, TIL$^{(Basic)}$=condensed, five-category TIL scale as defined in Table 1.

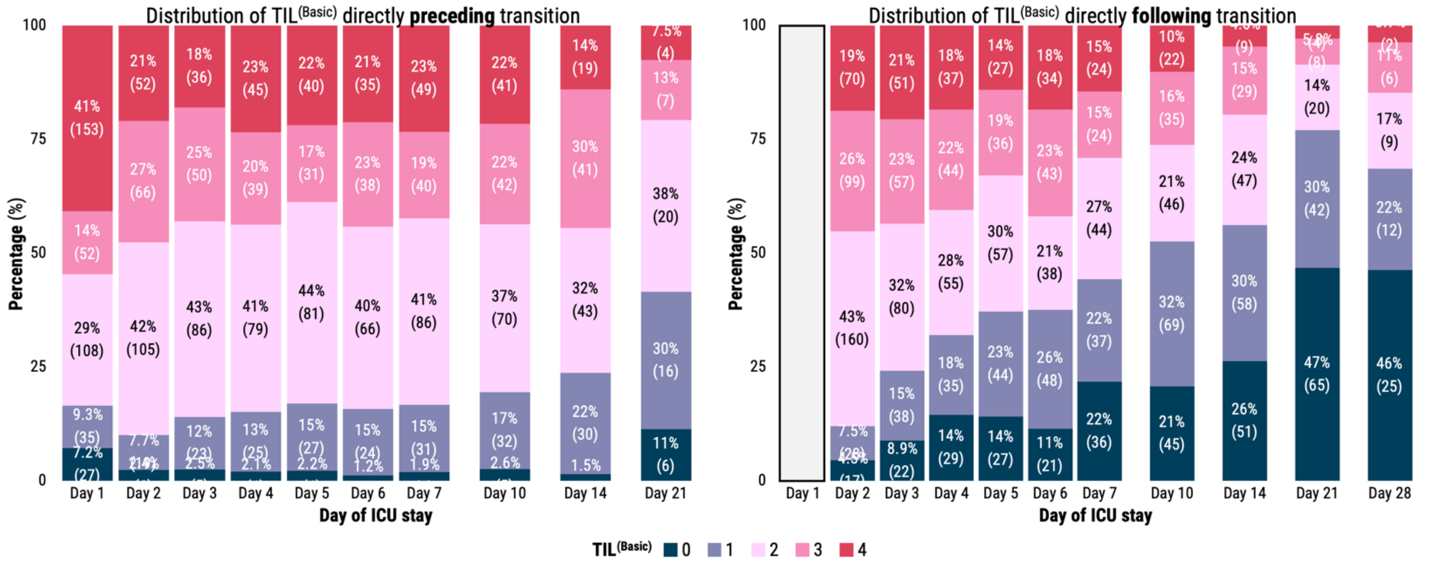


**Supplementary Fig. S4. Distributions of TIL$^{(Basic)}$ directly preceding/following a change in TIL$^{(Basic)}$.** Percentages are calculated out of the number of study patients who experienced a day-to-day change in TIL$^{(Basic)}$ either directly after (left-hand side) or directly before (right-hand side) the corresponding day of ICU stay. Abbreviations: ICU=intensive care unit, TIL=Therapy Intensity Level, TIL$^{(Basic)}$=condensed, five-category TIL scale as defined in Table 1.

**Supplementary Fig. S5. Population-level ΔTimeSHAP values stratified by pre-transition TIL$^{(Basic)}$ score.** Legend provided at end of figure (p. 7).

**a**

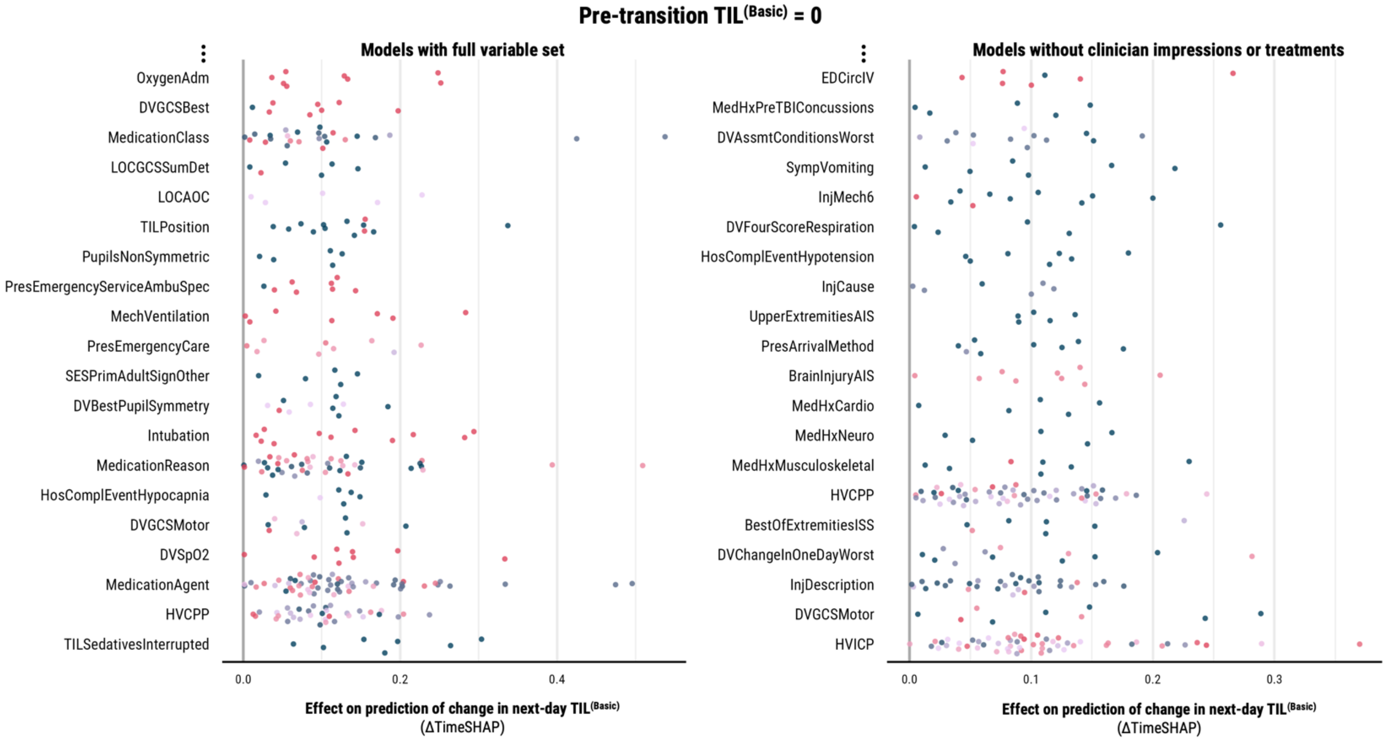


**b**

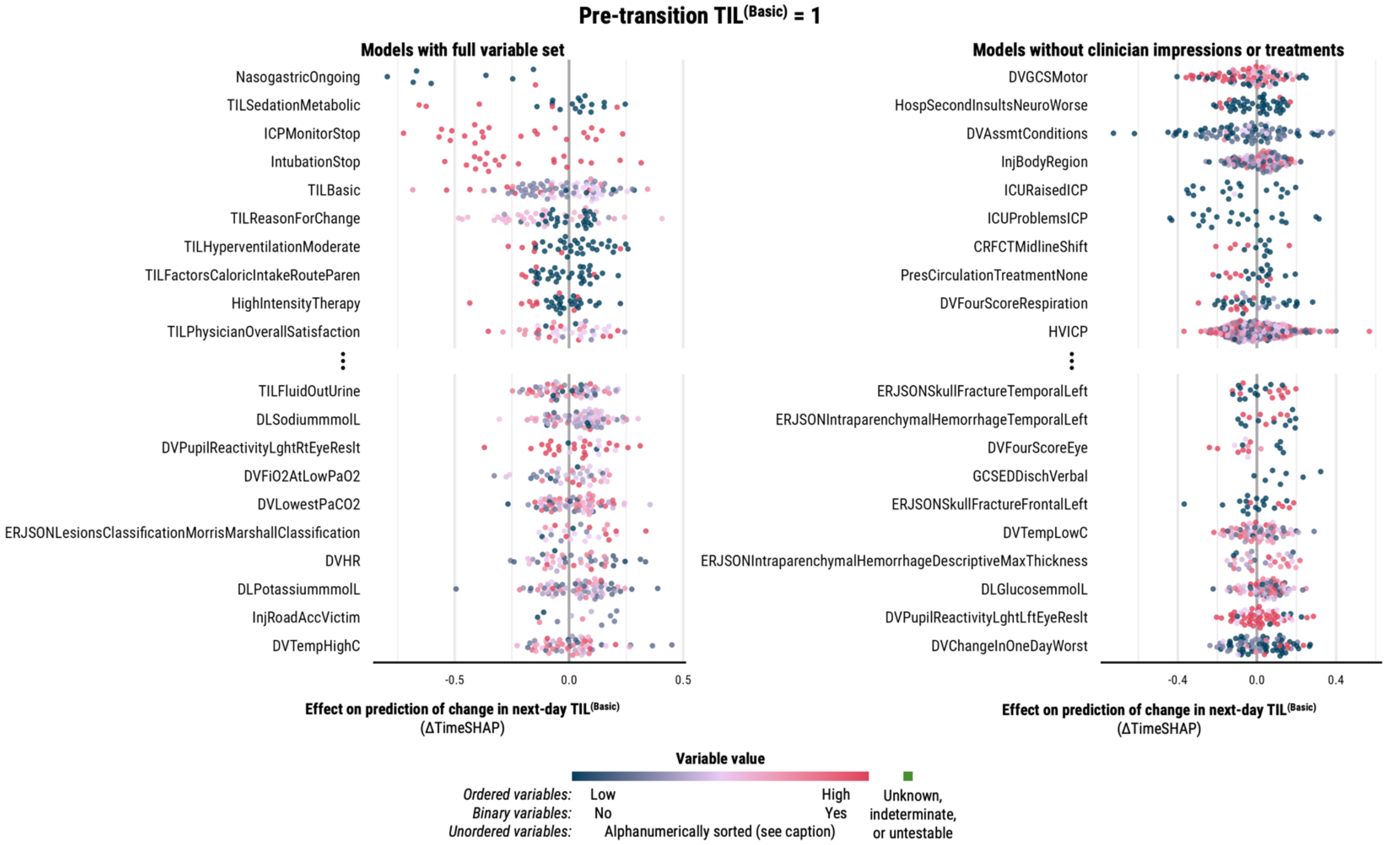

**Supplementary Fig. S5** (*continued*).

**c**

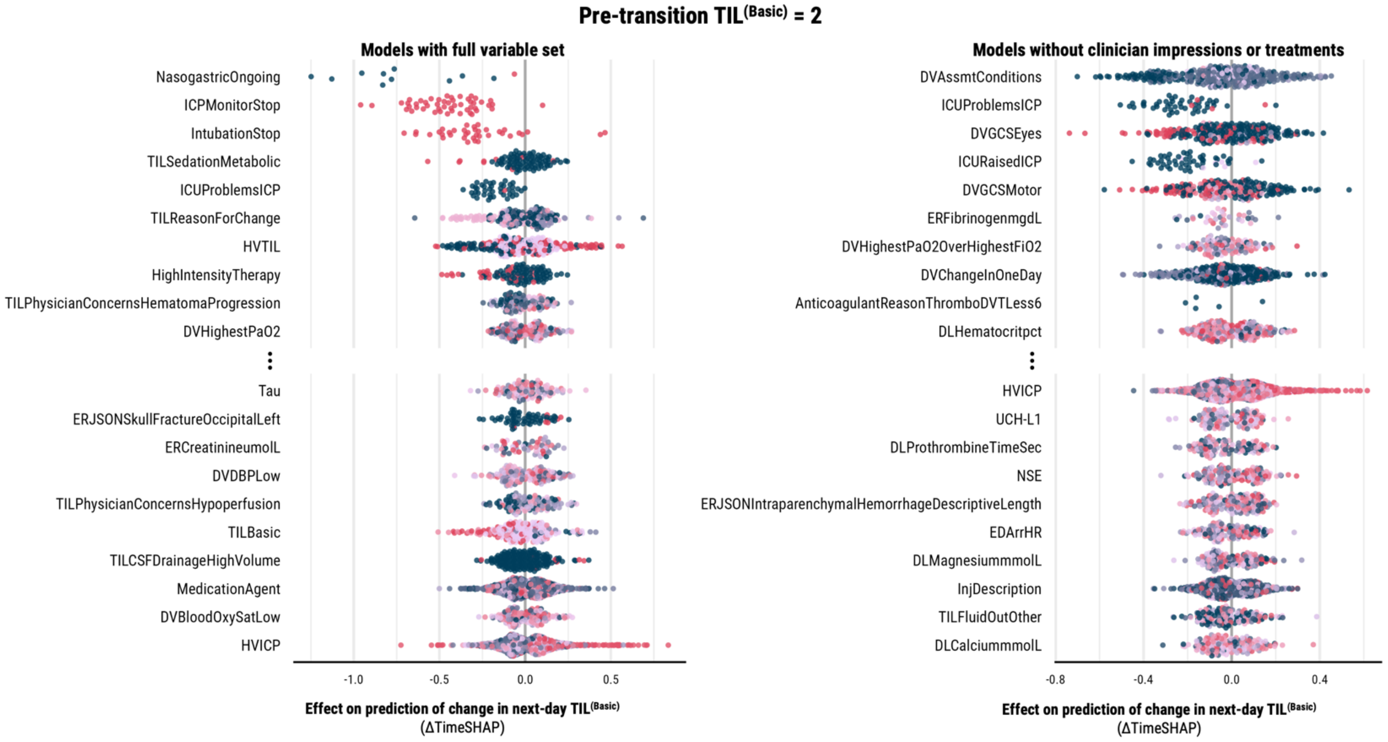


**d**

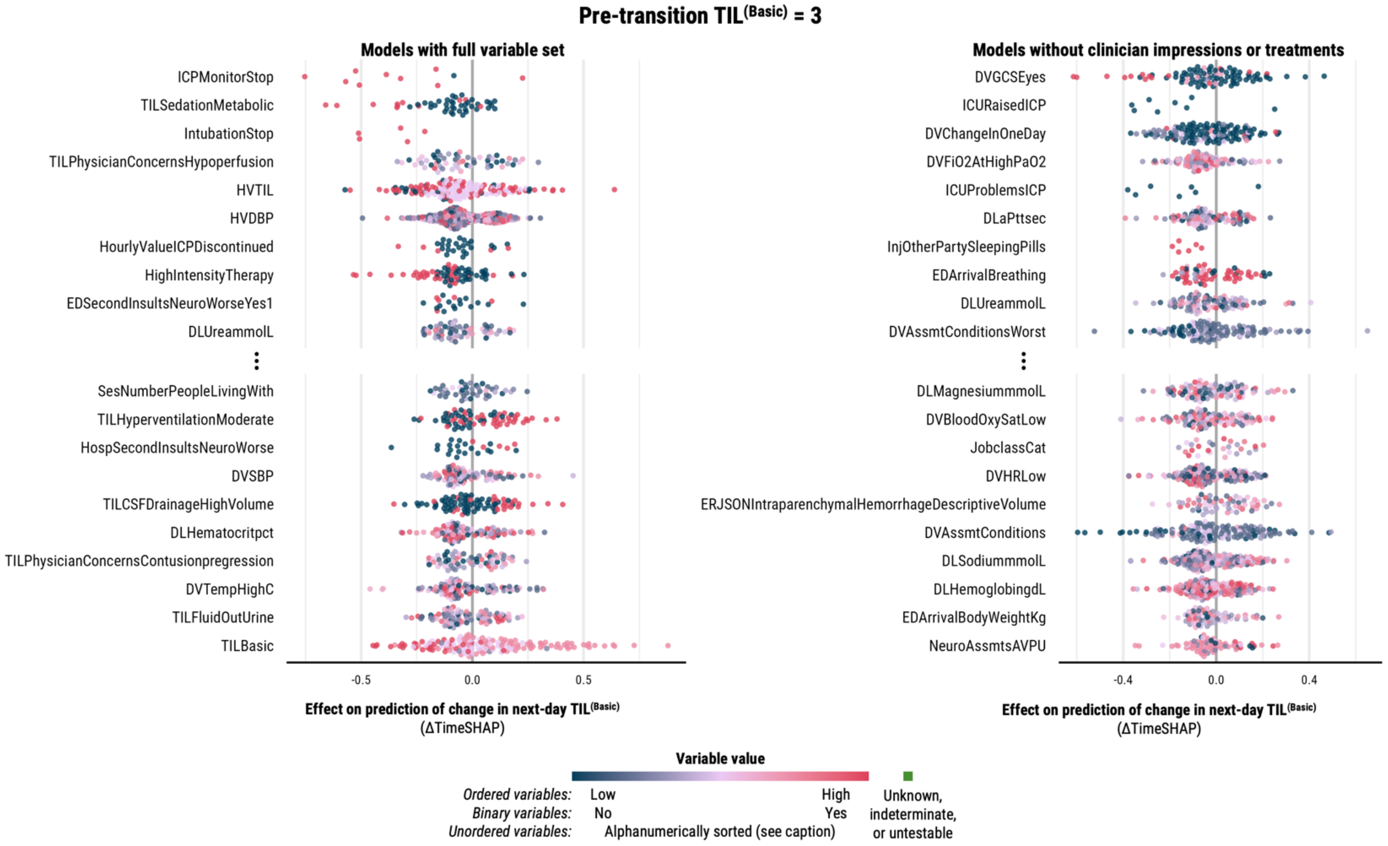

**Supplementary Fig. S5** (*continued*).

**e**

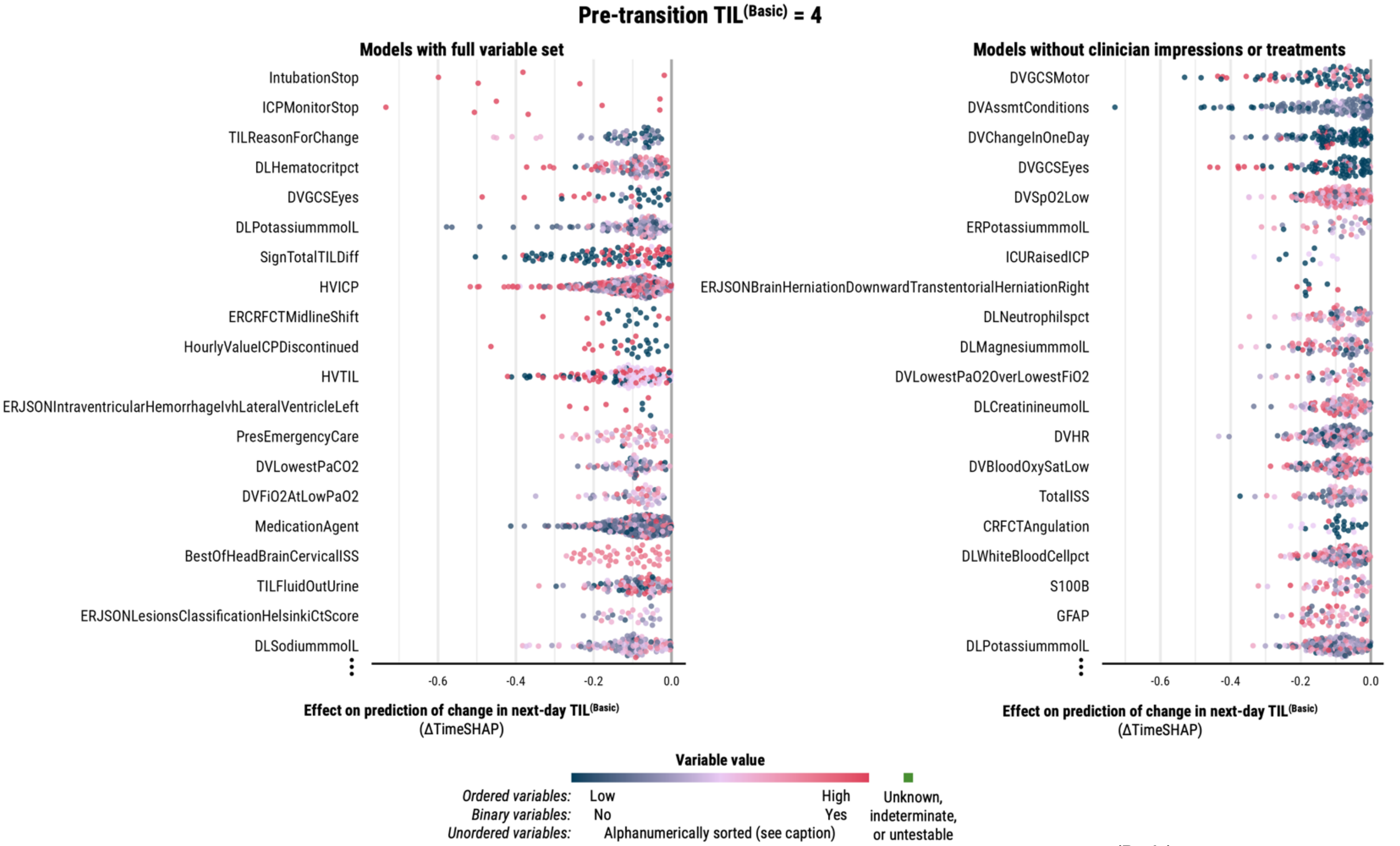


**Supplementary Fig. S5. Population-level ΔTimeSHAP values stratified by pre-transition TIL$^{(Basic)}$ score.** Within each panel (**a–e**), the ΔTimeSHAP values on the left-hand side are from the models trained on the full variable set whilst the ΔTimeSHAP values on the right-hand side are from the models trained without clinician impressions or treatments. ΔTimeSHAP values are interpreted as the relative contributions of variables towards the difference in model prediction of next-day TIL$^{(Basic)}$ over the two days directly preceding the change in TIL$^{(Basic)}$ (Supplementary Methods S5). Therefore, the study population represented in this figure is limited to patients who experienced a change in TIL$^{(Basic)}$ after day two of ICU stay (*n*=575). The variables were selected by first identifying the ten variables with non-missing value tokens with the most negative median ΔTimeSHAP values across the population (above the ellipses) and then, amongst the remaining variables, selecting the ten with non-missing value tokens with the most positive median ΔTimeSHAP values (below the ellipses). Each point represents the mean ΔTimeSHAP value, taken across all 20 repeated cross-validation partitions, for a token preceding an individual patient's change in TIL$^{(Basic)}$. The colour of the point represents the relative ordered value of a token within a variable, and for unordered variables (e.g., patient status during GCS assessment), tokens were sorted alphanumerically (the sort index per possible unordered variable token is provided in the CENTER-TBI data dictionary: https://www.center-tbi.eu/data/dictionary). All abbreviated variable names are decoded in the CENTER-TBI data dictionary.

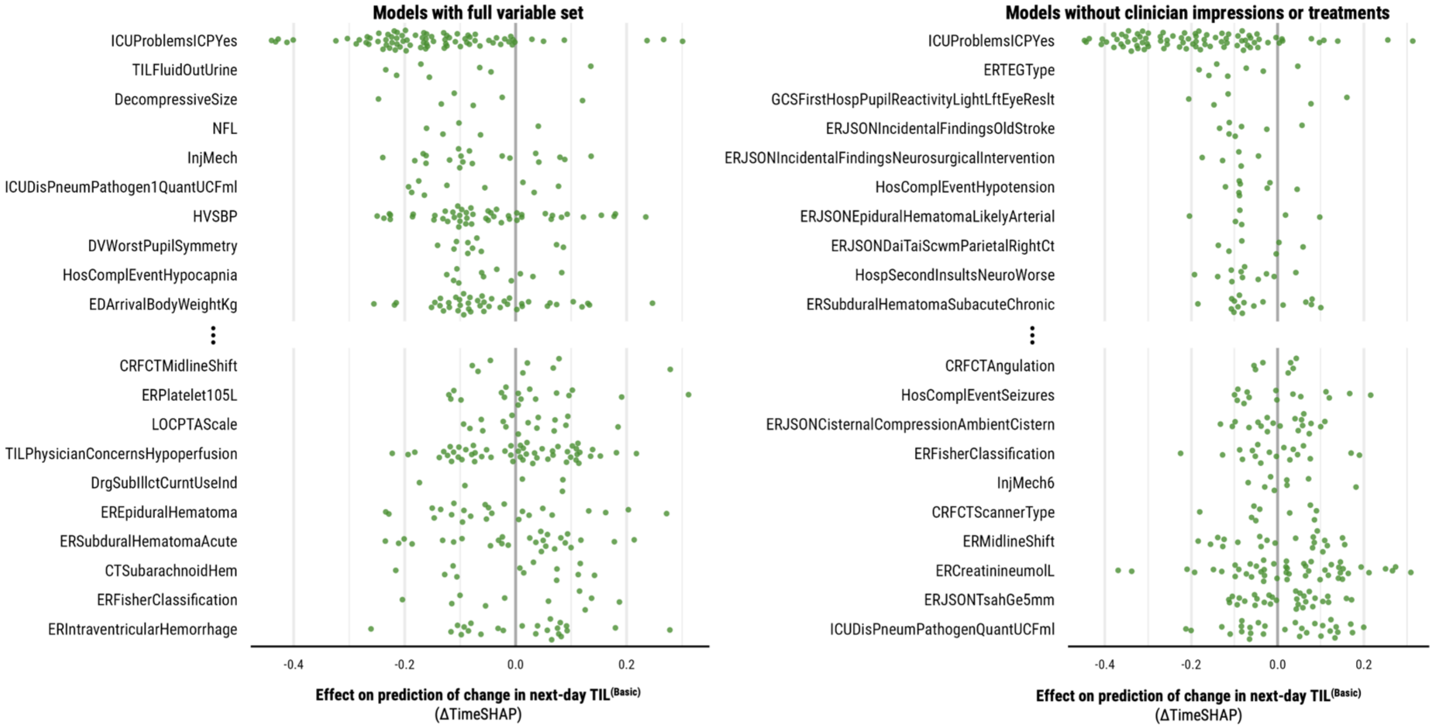


**Supplementary Fig. S6. Population-level ΔTimeSHAP values for missing value tokens.** The ΔTimeSHAP values on the left panel are from the models trained on the full variable set whilst the ΔTimeSHAP values on the right panel are from the models trained without clinician impressions or treatments. ΔTimeSHAP values are interpreted as the relative contributions of a variable's missingness towards the difference in model prediction of next-day TIL$^{(Basic)}$ over the two days directly preceding the change in TIL$^{(Basic)}$ (Supplementary Methods S5). Therefore, the study population represented in this figure is limited to patients who experienced a change in TIL$^{(Basic)}$ after day two of ICU stay (*n*=575). The variables were selected by first identifying the ten variables with missing value tokens with the most negative median ΔTimeSHAP values across the population (above the ellipses) and then, amongst the remaining variables, selecting the ten with missing value tokens with the most positive median ΔTimeSHAP values (below the ellipses). Each point represents the mean ΔTimeSHAP value, taken across all 20 repeated cross-validation partitions, for a token preceding an individual patient's change in TIL$^{(Basic)}$. All abbreviated variable names are decoded in the CENTER-TBI data dictionary: [https://www.center-tbi.eu/data/dictionary](https://www.center-tbi.eu/data/dictionary).

#### SUPPLEMENTARY TABLES

NOTE: Static variables are those with values fixed at ICU admission (e.g., helmet on during accident?). Intervention variable directly represent a treatment or management decision performed during a patient's ICU stay (i.e., administration of hypertonic saline). Since an intervention variable must take place during a patient's ICU stay, a variable cannot be both a static and an intervention variable. However, a variable can be both not static (i.e., dynamic) and not an intervention (e.g., a result from an ICU lab test or imaging report).

##### Supplementary Table S1. Manually excluded variables indicating death or withdrawal of life-sustaining treatment.

| Name | Format | Description | Possible Values |
|---|---|---|---|
| BrainDeathDate | date | Reflects the date of Brain death in case of Withdrawal of life-sustaining measures | Not Applicable |
| BrainDeathTime | text | Reflects the Time of Brain death in case of Withdrawal of life -sustaining measures Check also "Hospital.BrainDeathDate" for the date. | Not Applicable |
| CTPatientLocation | text | This variable describes the in-hospital location of the patient when the CT-scan was performed and was not meant to describe the locat ion of the CT-scanner. Three options: ER, Ward/Admission, ICU | ADMIS=Ward/Admission;ED=ER;ICU=ICU |
| DeadAge | integer | Reflects if the reason for Withdrawal of life-sustaining measures was age. | 0=No;1=Yes;88=Unknown |
| DeadCoMorbidities | integer | Reflects if the reason for Withdrawal of life-sustaining measures was co-morbidities. | 0=No;1=Yes;88=Unknown |
| DeadDeterminationOfBrainDe ath | integer | Reflects if the reason for Withdrawal of life-sustaining measures was Determination of brain death (acco rding to national law). | 0=No;1=Yes;88=Unknown |
| DeadOrganDonation | integer | Reflects if Withdrawal of life-sustaining measures was followed by organ donation. | 0=No;1=Yes;88=Unknown |
| DeadPatWill | integer | Reflects if the reason for Withdrawal of life-sustaining measures was Following living will of patient. | 0=No;1=Yes;88=Unknown |
| DeadRequestRelatives | integer | Reflects if the reason for Withdrawal of life-sustaining measures was On request of relatives. | 0=No;1=Yes;88=Unknown |
| DeadSeverityofTBI | integer | Reflects if the reason for Withdrawal of life-sustaining measures was Severity of TBI. | 0=No;1=Yes;88=Unknown |
| DeathAutopsy | integer | Reflects if an autopsy was performed after the death of the p atient. | 0=No;1=Yes, forensic;2=Yes, clinical;88=Unknown |
| DeathCause | integer | Cause of death in or outside the hospital | 1=Head injury/initial injury;2=Head injury/secondary intracranial damage;3=Systemic trauma;4=Medical complications;88=Unknown;99=Other |
| DeathCauseOther | text | "Other" cause of death in or outside the hospital (than predef ined list). | Not Applicable |
| DeathDate | date | Date of death also recorded on hospital discharge and at foll owup: FollowUp.FUPrincipalDeathCause;Death may also ha ve been recorded in the ER forms: Subject.DeathDate | Not Applicable |
| DeathERDeclaredBrainDeadF ollowingNationalCriteria | integer | Reflects if patient was declared brain dead following national criteria. Only applicable if patient declared "dead" on the ER | 0=No;1=Yes;88=Unknown |
| DeathERDOA | integer | Reflects if patient is declared dead on the ER -- > Dead on arrival (DOA). Only applicable if patient declared "dead" on the ER. | 0=No;1=Yes;88=Unknown |
| DeathERUnsuccResusForExt raCranInj | integer | Reflects unsuccessful resuscitation for extra cranial injuries if patient is declared dead on the ER. Only applicable if patient declared "dead" on the ER | 0=No;1=Yes;88=Unknown |
| DeathERWithdrawalLifeSupp ForSeverityOfTBI | integer | Reflects Withdrawal of life¬≠sustaining measures for severity of TBI if patient is declared dead on the ER. Only applicable if patient declared "dead" on the ER | 0=No;1=Yes;88=Unknown |
| DeathTime | text | Time of death | Not Applicable |
| DischargeStatus | integer | Assessment by investigator Reflects if the patient was dead or alive at discharge. | 0=Dead;1=Alive;88=Unknown |
| EOSReason | integer | Reason for end of study participation | 1=Completion of study;2=Inability to obtain follow-up;3=Withdrawal from study (by patient or represen tative);4=Adverse event(s);5=Decision for DNR*:;6=Withdrawal of support;7=Death;99=Other |
| EOSReasonOtherTxt | text | "Other" reason for end of study participation than the predefi ned list. | Not Applicable |
| ICDCode1 | text | Up to 16 fields available to enter diagnosis as recorded by h ospital administration according to ICD codes; applicable to patients admitted/discharged from hospital. For patients discharged directly from the ER, ICD codes are documented in: InjuryHx.ERDestICDCodes1 | Not Applicable |

| Name | Format | Description | Possible Values |
| --- | --- | --- | --- |
| ICDCode2 | text | Up to 16 fields available to enter diagnosis as recorded by h ospital administration according to ICD codes; applicable to patients admitted/discharged from hospital.<br>For patients discharged directly from the ER, ICD codes are documented in: InjuryHx.ERDestICDCodes1 | Not Applicable |
| ICDCode3 | text | Up to 16 fields available to enter diagnosis as recorded by h ospital administration according to ICD codes; applicable to patients admitted/discharged from hospital.<br>For patients discharged directly from the ER, ICD codes are documented in: InjuryHx.ERDestICDCodes1 | Not Applicable |
| ICDCode4 | text | Up to 16 fields available to enter diagnosis as recorded by h ospital administration according to ICD codes; applicable to patients admitted/discharged from hospital.<br>For patients discharged directly from the ER, ICD codes are documented in: InjuryHx.ERDestICDCodes1 | Not Applicable |
| ICDCode5 | text | Up to 16 fields available to enter diagnosis as recorded by h ospital administration according to ICD codes; applicable to patients admitted/discharged from hospital.<br>For patients discharged directly from the ER, ICD codes are documented in: InjuryHx.ERDestICDCodes1 | Not Applicable |
| ICDCode6 | text | Up to 16 fields available to enter diagnosis as recorded by h ospital administration according to ICD codes; applicable to patients admitted/discharged from hospital.<br>For patients discharged directly from the ER, ICD codes are documented in: InjuryHx.ERDestICDCodes1 | Not Applicable |
| ICDCodeVersion | integer | This variable reflects if the ICD code version 9 or version 10 was used.<br>Up to 16 fields are available to enter diagnosis as recorded b y hospital administration according to ICD codes. | 9=ICD-9;10=ICD-10 |
| ICPStopReason | integer | Reason for stopping ICP. Also check Hospital.ICPMonitorSto p. | 1=Clinically improved;2=ICP stable and < 20 mmHg ;3=Monitor/catheter failure;4=Patient considered un salvagable;5=Patient died;99=Other |
| ICPStopReason | integer | Reason for stopping ICP. Also check Hospital.ICPMonitorSto p. | 1=Clinically improved;2=ICP stable and < 20 mmHg ;3=Monitor/catheter failure;4=Patient considered un salvagable;5=Patient died;99=Other |
| ICUDischargeICDCode1 | text | The intent here is to register ICD code as recorded in hospit al administrative files for patients directly discharged from th e ICU. Up to 16 codes can be entered, ICD codes are further captured at ER discharge and at hospital discharge. | Not Applicable |
| ICUDischargeICDCode2 | text | The intent here is to register ICD code as recorded in hospit al administrative files for patients directly discharged from th e ICU. Up to 16 codes can be entered, ICD codes are further captured at ER discharge and at hospital discharge. | Not Applicable |
| ICUDischargeICDCode3 | text | The intent here is to register ICD code as recorded in hospit al administrative files for patients directly discharged from th e ICU. Up to 16 codes can be entered, ICD codes are further captured at ER discharge and at hospital discharge. | Not Applicable |
| ICUDischargeICDCode4 | text | The intent here is to register ICD code as recorded in hospit al administrative files for patients directly discharged from th e ICU. Up to 16 codes can be entered, ICD codes are further captured at ER discharge and at hospital discharge. | Not Applicable |
| ICUDischargeICDCode5 | text | The intent here is to register ICD code as recorded in hospit al administrative files for patients directly discharged from th e ICU. Up to 16 codes can be entered, ICD codes are further captured at ER discharge and at hospital discharge. | Not Applicable |
| ICUDischargeICDCode6 | text | The intent here is to register ICD code as recorded in hospit al administrative files for patients directly discharged from th e ICU. Up to 16 codes can be entered, ICD codes are further captured at ER discharge and at hospital discharge. | Not Applicable |
| ICUDischargeICDCodeVersio n | integer | This variable reflects if the ICD code version 9 or version 10 was used.<br>Up to 16 fields are available to enter diagnosis as recorded b y hospital administration according to ICD codes. | 9=9;10=10 |
| ICUDischargeStatus | integer | Reflects if patient was alive or dead on discharge from ICU | 1=Alive;2=Dead;88=Unknown |
| ICUDischargeTo | integer | Reflects location to which the patient was discharged from I CU | 1=General ward;2=Other ICU;3=Other hospital;4=R ehab unit;5=Home;6=Nursing home;7=Step down/h igh care unit;88=Unknown;99=Other |
| ICUDischargeToOther | text | Specifies the "other" location to which the patient was discha rged from ICU Check also "Hospital.ICUDischargeTo" | Not Applicable |
| ICUDisPatDeadAtICU | integer | Reflects if the patient was declared dead on the ICU.<br>Intended as an introductory question for the details on withdr awal of treatment, brain death and organ donation | 0=No;1=Yes |

| Name | Format | Description | Possible Values |
|---|---|---|---|
| ICUDisSupportWithdrawnDate | date | This variable documents date and time at which life prolonging therapy was withdrawn (together with "Hospital.ICUDisSupportWithdrawnTime") | Not Applicable |
| ICUDisSupportWithdrawnTime | text | This variable documents date and time at which life prolonging therapy was withdrawn (together with "Hospital.ICUDisSupportWithdrawnDate") | Not Applicable |
| ICUDisWithdrawalTreatmentDecisionDate | date | Investigators were requested to record the details of Withdrawal of Treatment or Life support if applicable. | Not Applicable |
| ICUDisWithdrawalTreatmentDecisionTime | text | Investigators were requested to record the details of Withdrawal of Treatment or Life support if applicable. | Not Applicable |
| ICUDisWithdrawlTreatmentDecision | integer | Investigators were requested to record the details of Withdrawal of Treatment or Life support if applicable. | 1=Multi disciplinary;2=By a single physician;3=With relatives |
| IntubationStopReason | integer | This variable describes the Stop Reason of Extubation in case of Ventilation Management (only for ICU patients). | 1=Respiratory stable;2=Accidental;3=Withdrawal of care |
| LengthOfStay | decimal | This variable reflects the length of stay of the patient at the study hospital.<br>It has been derived using the information of the date and time of arrival at the study hospital and date and time of (study) hospital discharge. | Not Applicable |
| MonJugularSatStopReason | integer | Beside brain specific ICP monitoring in the ICU, details were recorded on other types of monitoring in the ICU.<br>This variable reflects the reason for stopping if there was Jugular oximetry. | 1=Monitor/catheter failure;2=Patient considered unsalvageable;3=Patient died;4=Clinically no longer required |
| MonLicoxStopReason | integer | Beside brain specific ICP monitoring in the ICU, details were recorded on other types of monitoring in the ICU.<br>This variable reflects the reason for stopping if there was Brain tissue PO2 monitoring. | 1=Monitor/catheter failure;2=Patient considered unsalvageable;3=Patient died;4=Clinically no longer required |
| MonLicoxStopReason | integer | Beside brain specific ICP monitoring in the ICU, details were recorded on other types of monitoring in the ICU.<br>This variable reflects the reason for stopping if there was Brain tissue PO2 monitoring. | 1=Monitor/catheter failure;2=Patient considered unsalvageable;3=Patient died;4=Clinically no longer required |
| MonMicrodialysisStopReason | integer | Beside brain specific ICP monitoring in the ICU, details were recorded on other types of monitoring in the ICU.<br>This variable reflects the reason for stopping if there was Microdialysis. | 1=Monitor/catheter failure;2=Patient considered unsalvageable;3=Patient died;4=Clinically no longer required |
| OrganDonationDate | date | Reflects the date of organ donation in case of Withdrawal of life-sustaining measures, if applicable. | Not Applicable |
| OrganDonationTime | text | Reflects the time of organ donation in case of Withdrawal of life-sustaining measures, if applicable. | Not Applicable |
| SupportWithdrawnDate | date | Investigators were requested to record the details of Withdrawal of Treatment or Life support if applicable. | Not Applicable |
| SupportWithdrawnTime | text | Investigators were requested to record the details of Withdrawal of Treatment or Life support if applicable. | Not Applicable |
| TimeSinceICUAdmisDeath | text | Investigators were requested to record the details of Withdrawal of Treatment or Life support if applicable.<br>This reflects the time between admission in the ICU and death | Not Applicable |
| WithdrawalOption | integer | In case of complete withdrawal, all data have been deleted from the database | 1=Complete Withdrawal (no further contact, destruction of all data and samples collected up to that point);2=No further study related activities, but consent to access of clinical notes and use of existing data |
| WithdrawalTreatmentDecision | integer | Intended only to be scored if a medical decision was made to withdraw active treatment because of anticipated poor prognosis. However, some investigators may have scored this when patients had recovered to an extent that active treatment was no longer necessary. | 1=Multi disciplinary;2=By a single physician;3=With relatives |
| WithdrawalTreatmentDecisionDate | date | Investigators were requested to record the details of Withdrawal of Treatment or Life support if applicable.<br>Intended only to be scored if a medical decision was made to withdraw active treatment because of anticipated poor prognosis. However, some investigators may have scored this when patients had recovered to an extent that active treatment was no longer necessary. | Not Applicable |
| WithdrawalTreatmentDecisionTime | text | Investigators were requested to record the details of Withdrawal of Treatment or Life support if applicable.<br>Intended only to be scored if a medical decision was made to withdraw active treatment because of anticipated poor prognosis. However, some investigators may have scored this when patients had recovered to an extent that active treatment was no longer necessary. | Not Applicable |

| Name | Format | Description | Possible Values |
|---|---|---|---|
| WithdrawSuppDateTime | datetime | Withdrawal of Support Date & Time if Withdrawal of life-sustaining support was the reason for end of study participation. | Not Applicable |

#### Supplementary Table S2. Physician-based impression variables.

| Name | Static | Format | Description | Possible Values |
|---|---|---|---|---|
| InjAIS | TRUE | integer | In the original AIS classification of injury severity, the grading is from 1 (minor) to 6 (unsurvivable). We added a score of 0 to designate absence of injuries. This is the AIS score for body regions as specified by AIS.InjBodyRegion. | 0=None;1=Minor: no treatment needed;2=Moderate: requires only outpatient treatment;3=Serious: requires non-ICU hospital admission;4=Severe: requires ICU observation and/or basic treatment;5=Critical: requires intubation, mechanical ventilation or vasopressors for blood pressure support;6=Unsurvivable: not survivable |
| HVTILChangeReason | FALSE | integer | Bihourly reason for change in therapy intensity level | 1=Intensified: Clinical deterioration;2=Intensified: Suspicion of increased of ICP (not measured);3=Intensified: Increased ICP (documented);4=Intensified: Clinical decision to target other mechanism;5=Intensified: Change of doctor (different shift);6=Decreasing: Clinical improvement;7=Decreasing: Adequate control over ICP;8=Decreasing: Upper treatment limit reached/past;9=Decreasing: Further treatment considered futile;10=Decreasing: Change of doctor (different shift) |
| TILPhysicianOverallSatisfaction | FALSE | integer | This variable aims to capture the overall satisfaction of the physician with the clinical course of this patient; "not at all satisfied" would indicate that the patient did much more poorly than expected; "very satisfied" would indicate that the patient did much better than expected. Physician satisfaction should be assessed on a daily basis, | 0=Not at all;1=Slightly;2=Moderately;3=Quite;4=Very |
| TILPhysicianOverallSatisfactionSurvival | FALSE | integer | This variable aims to capture the opinion of the treating physician as to whether the short time survival change have chnged in comparision to the previous assessment | 1=Much worse;2=A little worse;3=Unchanged;4=A little better;5=Much better |
| TILReasonForChange | FALSE | integer | Reflects the reason for change in TIL therapy over the day. | 0=No change;1=Intensified: Clinical deterioration;2=Intensified:Suspicion of increased of ICP (not measured);3=Intensified:Increased ICP (documented);4=Intensified:Clinical decision to target other mechanism;5=Intensified:Change of doctor (different shift);6=Decreasing:Clinical improvement;7=Decreasing:Adequate control over ICP;8=Decreasing:Upper treatment limit reached/past;9=Decreasing:Further treatment considered futile;10=Decreasing:Change of doctor (different shift) |
| TotalTIL | FALSE | integer | Calculated centrally - 24 hour TILS as the worst sum TILs for each day for the ICU timepoints (day 1-7, 10, 14, 21 and 28) | Not Applicable |
| AbdomenPelvicContentsAIS | TRUE | integer | AIS score for the Abdomen/Pelvic Contents In the original AIS classification of injury severity, the grading is from 1 (minor) to 6 (unsurvivable). We added a score of 0 to designate absence of injuries. | 0=None;1=Minor: no treatment needed;2=Moderate: requires only outpatient treatment;3=Serious: requires non-ICU hospital admission;4=Severe: requires ICU observation and/or basic treatment;5=Critical: requires intubation, mechanical ventilation or vasopressors for blood pressure support;6=Unsurvivable: not survivable |
| BaselineGOS6MoExpectedDeathRisk | TRUE | text | At ER discharge, physician estimate of six month outcome was recorded as a baseline risk assessment: "Given all current available information, what is, in your subjective opinion, the most likely 6-month outcome of this patient? To be based upon information on discharge ER or admission to hospital/ICU". This reflects the Risk of death in % | Not Applicable |
| BaselineGOS6MoExpectedOutcome | TRUE | text | At ER discharge, physician estimate of six month outcome was recorded as a baseline risk assessment: "Given all current available information, what is, in your subjective opinion, the most likely 6-month outcome of this patient? To be based upon information on discharge ER or admission to hospital/ICU". This reflects the Expected outcome (GOS) | D=D - Death;GR=GR - Good Recovery;MD=MD - Moderate Disability;SD=SD - Severe Disability;V=V - Vegetative State |
| BaselineGOS6MoUnfavourableOutcomeRisk | TRUE | text | At ER discharge, physician estimate of six month outcome was recorded as a baseline risk assessment: "Given all current available information, what is, in your subjective opinion, the most likely 6-month outcome of this patient? To be based upon information on discharge ER or admission to hospital/ICU". This reflects the Risk of unfavorable outcome (D, VS, SD) in % | Not Applicable |
| BaselinePhysEstOf6MoOutcomePhysicianQual | TRUE | integer | At ER discharge, physician estimate of six month outcome was recorded as a baseline risk assessment: "Given all current available information, what is, in your | 1=Resident;2=Junior staff (< 5 years);3=Senior staff (>= 5 years);4=Head of department |

| | | | | |
|---|---|---|---|---|
| | | | subjective opinion, the most likely 6-month outcome of this patient? To be based upon information on discharge ER or admission to hospital/ICU". This reflects the qualification of the physician who provided prognostic estimate on ER discharge/admission to hospital/ICU | |
| BaselinePhysEstOf6MoOutcomePhysicianType | TRUE | integer | At ER discharge, physician estimate of six month outcome was recorded as a baseline risk assessment: "Given all current available information, what is, in your subjective opinion, the most likely 6-month outcome of this patient? To be based upon information on discharge ER or admission to hospital/ICU". This reflects the type of the physician who provided prognostic estimate on ER discharge/admission to hospital/ICU | 1=ER Physician;2=Intensive Care;3=Neurology;4=Neurosurgery;5=Traumatology;88=Unknown |
| BestOfAbdomenPelvicLumbarISS | TRUE | integer | AbdomenPelvicLumbar region (Highest AIS of the region)^2 compare AbdomenPelvicContentsAIS, LumbarSpineAIS. This score is taken forward for ISS calculation | Not Applicable |
| BestOfChestSpineISS | TRUE | integer | (highest AIS of the region)^2 Compare ThoraxChestAIS, ThoracicSpineAIS and select the highest for ISS calculation | Not Applicable |
| BestOfExternalISS | TRUE | integer | External region (ExternaAIS)^2 select the highest external AIS severity code for ISS calculation. | Not Applicable |
| BestOfExtremitiesISS | TRUE | integer | Extremities region (Highest AIS of the region)^2 compare UpperExtremitiesAIS, LowerExtremitiesAIS, PelvicGirdleAIS select the highest for ISS calculation | Not Applicable |
| BestOfFaceISS | TRUE | integer | Face region (FaceAIS)^2 select the highest facial injury for ISS calculation | Not Applicable |
| BestOfHeadBrainCervicalISS | TRUE | integer | HeadBrainCervical region (Highest AIS of the region)^2 Compare HeadNeckAIS, InjuryHx.BrainInjuryAIS, CervicalSpineAIS select the highest scoring injury in any of these 3 areas for ISS calculation | Not Applicable |
| BrainInjuryAIS | TRUE | integer | AIS score for the Brain Injury In the original AIS classification of injury severity, the grading is from 1 (minor) to 6 (unsurvivable). We added a score of 0 to designate absence of injuries. | 0=None;1=Minor: no treatment needed;2=Moderate: requires only outpatient treatment;3=Serious: requires non-ICU hospital admission;4=Severe: requires ICU observation and/or basic treatment;5=Critical: requires intubation, mechanical ventilation or vasopressors for blood pressure support;6=Unsurvivable: not survivable |
| CervicalSpineAIS | TRUE | integer | AIS score for the Cervical Spine region. In the original AIS classification of injury severity, the grading is from 1 (minor) to 6 (unsurvivable). We added a score of 0 to designate absence of injuries. | 0=None;1=Minor: no treatment needed;2=Moderate: requires only outpatient treatment;3=Serious: requires non-ICU hospital admission;4=Severe: requires ICU observation and/or basic treatment;5=Critical: requires intubation, mechanical ventilation or vasopressors for blood pressure support;6=Unsurvivable: not survivable |
| DispER | TRUE | integer | Destination of the patient at ER discharge. | 1=Discharge home;2=Discharge other facility;3=Hospital admission--Ward;4=Hospital admission--Intermediate/high care unit;5=Hospital admission--ICU;6=Hospital admission--OR for immediate surgical procedure;7=Death;8=Hospital admission--Other (e.g. observation unit);88=Unknown |
| EmerSurgIntraCranSurviveNoSurg | TRUE | integer | "InjuryHx.EmerSurgIntraCranSurviveNoSurg" and "InjuryHx.EmerSurgIntraCranSurviveYesSurg" These 2 variables aim to capture information on the surgeon's expectations, eg if the surgeon considers a realistic expectation of benefit, or performs the surgery as a "last resort" in a likely hopeless case. 'The short term survival chances of the patients if I DO NOT operate will be (in %)' | Not Applicable |
| EmerSurgIntraCranSurviveYesSurg | TRUE | integer | "InjuryHx.EmerSurgIntraCranSurviveNoSurg" and "InjuryHx.EmerSurgIntraCranSurviveYesSurg" These 2 variables aim to capture information on the surgeon's expectations, eg if the surgeon considers a realistic expectation of benefit, or performs the surgery as a "last resort" in a likely hopeless case. 'The short term survival chances of the patients if I DO operate will be (in %)' | Not Applicable |
| ExternaAIS | TRUE | integer | AIS score for the External skin In the original AIS classification of injury severity, the grading is from 1 (minor) to 6 (unsurvivable). We added a score of 0 to designate absence of injuries. | 0=None;1=Minor: no treatment needed;2=Moderate: requires only outpatient treatment;3=Serious: requires non-ICU hospital admission;4=Severe: requires ICU observation and/or basic treatment;5=Critical: requires intubation, mechanical ventilation or vasopressors for blood pressure support;6=Unsurvivable: not survivable |
| FaceAIS | TRUE | integer | AIS score for Face (incl.maxillofacial) In the original AIS classification of injury severity, the grading is from 1 (minor) to 6 (unsurvivable). We added a score of 0 to designate absence of injuries. | 0=None;1=Minor: no treatment needed;2=Moderate: requires only outpatient treatment;3=Serious: requires non-ICU hospital admission;4=Severe: requires ICU observation and/or basic treatment;5=Critical: requires |

| | | | | |
|---|---|---|---|---|
| | | | | intubation, mechanical ventilation or vasopressors for blood pressure support;6=Unsurvivable: not survivable |
| HeadNeckAIS | TRUE | integer | AIS score for the Head Neck region In the original AIS classification of injury severity, the grading is from 1 (minor) to 6 (unsurvivable). We added a score of 0 to designate absence of injuries. | 0=None;1=Minor: no treatment needed;2=Moderate: requires only outpatient treatment;3=Serious: requires non-ICU hospital admission;4=Severe: requires ICU observation and/or basic treatment;5=Critical: requires intubation, mechanical ventilation or vasopressors for blood pressure support;6=Unsurvivable: not survivable |
| LowerExtremitiesAIS | TRUE | integer | AIS score for the Lower extremities. In the original AIS classification of injury severity, the grading is from 1 (minor) to 6 (unsurvivable). We added a score of 0 to designate absence of injuries. | 0=None;1=Minor: no treatment needed;2=Moderate: requires only outpatient treatment;3=Serious: requires non-ICU hospital admission;4=Severe: requires ICU observation and/or basic treatment;5=Critical: requires intubation, mechanical ventilation or vasopressors for blood pressure support;6=Unsurvivable: not survivable |
| LumbarSpineAIS | TRUE | integer | AIS score for the Lumbar Spine region. In the original AIS classification of injury severity, the grading is from 1 (minor) to 6 (unsurvivable). We added a score of 0 to designate absence of injuries. | 0=None;1=Minor: no treatment needed;2=Moderate: requires only outpatient treatment;3=Serious: requires non-ICU hospital admission;4=Severe: requires ICU observation and/or basic treatment;5=Critical: requires intubation, mechanical ventilation or vasopressors for blood pressure support;6=Unsurvivable: not survivable |
| PelvicGirdleAIS | TRUE | integer | AIS score for the Pelvic Girdle region. In the original AIS classification of injury severity, the grading is from 1 (minor) to 6 (unsurvivable). We added a score of 0 to designate absence of injuries. | 0=None;1=Minor: no treatment needed;2=Moderate: requires only outpatient treatment;3=Serious: requires non-ICU hospital admission;4=Severe: requires ICU observation and/or basic treatment;5=Critical: requires intubation, mechanical ventilation or vasopressors for blood pressure support;6=Unsurvivable: not survivable |
| SurgIntervenAppro | TRUE | integer | WHY question: How strongly does the surgeon feels that this surgical intervention is appropriate in terms of the expected benefit to final clinical outcome? | 0=0;1=1;2=2;3=3;4=4;5=5;6=6;7=7;8=8;9=9;10=10 |
| ThoracicSpineAIS | TRUE | integer | AIS score for the Thoracic spine Region In the original AIS classification of injury severity, the grading is from 1 (minor) to 6 (unsurvivable). We added a score of 0 to designate absence of injuries. | 0=None;1=Minor: no treatment needed;2=Moderate: requires only outpatient treatment;3=Serious: requires non-ICU hospital admission;4=Severe: requires ICU observation and/or basic treatment;5=Critical: requires intubation, mechanical ventilation or vasopressors for blood pressure support;6=Unsurvivable: not survivable |
| ThoraxChestAIS | TRUE | integer | AIS score for the Thorax Chest region. In the original AIS classification of injury severity, the grading is from 1 (minor) to 6 (unsurvivable). We added a score of 0 to designate absence of injuries. | 0=None;1=Minor: no treatment needed;2=Moderate: requires only outpatient treatment;3=Serious: requires non-ICU hospital admission;4=Severe: requires ICU observation and/or basic treatment;5=Critical: requires intubation, mechanical ventilation or vasopressors for blood pressure support;6=Unsurvivable: not survivable |
| TotalISS | TRUE | integer | The Injury Severity Score is calculated as the sum of the squares of the the 3 body regions with the highest AIS score. The max score for the ISS = 75. If any body region AIS is assigned a score of "6", the ISS is automatically set to 75 (highest score). In the calculation of the ISS, only the 6 main body regions are taken into consideration. | Not Applicable |
| UpperExtremitiesAIS | TRUE | integer | AIS score for the Upper extremities. In the original AIS classification of injury severity, the grading is from 1 (minor) to 6 (unsurvivable). We added a score of 0 to designate absence of injuries. | 0=None;1=Minor: no treatment needed;2=Moderate: requires only outpatient treatment;3=Serious: requires non-ICU hospital admission;4=Severe: requires ICU observation and/or basic treatment;5=Critical: requires intubation, mechanical ventilation or vasopressors for blood pressure support;6=Unsurvivable: not survivable |
| CRFCTReason | FALSE | text | This variable contains the main reason why a CT-scan, during hospital stay, was performed. One of following options: standard follow-up, post-operative control, clinical deterioration, (suspicion of) increasing ICP, lack of improvement, unknown, other (specified in CTMRI.CTReasonOther) The reason for making an early CT-scan/ER scan can be found in: CTMRI.CTERReason | CD=Clinical deterioration;ICUADM88=Unknown;ICUADM99=Other;IICP=(Suspicion of) Increasing ICP;LOP=Lack of improvement;POC=Post-operative control;SFU=Standard follow-up |
| CTNoOpMotiv | FALSE | integer | WHY question: documents reason for not having an indication for (intra)cranial surgery. | 0=No surgical lesion;1=Lesion present, but Acceptable/good neurologic condition;2=Lesion present, but Guideline adherence;3=Lesion present, but Little/no mass effect;4=Lesion present, but Not hospital policy;5=Lesion present, but Extremely poor prognosis;6=Lesion present, but Brain Death;7=Lesion present, but Old age;8=Lesion present, but Wish family;88=Unknown;99=Lesion present, but Other |
| CTNoOpMotivOther | FALSE | text | Specification, only applicable if "CTMRI.CTNoOpMotiv" was "other" | Not Applicable |
| CTYesOpMotiv | FALSE | integer | WHY question: documents reason for having an indication for (intra)cranial surgery. | 1=Emergency/life saving;2=Clinical deterioration;3=Mass effect on CT;4=Radiological progression;5=(Suspicion |

| | | | | |
|---|---|---|---|---|
| | | | | of) raised ICP;6=Guideline adherence;7=To prevent deterioration;8=Depressed skull fracture;99=Other |
| CTYesOpMotivOther | FALSE | text | Free text if "CTMRI.CTYesOpMotiv" was marked as 'Other'. Relates to the WHY question: documents reason for having an indication for (intra)cranial surgery. | Not Applicable |
| ERCTNoOpMotiv | TRUE | integer | In emergency room: WHY question: documents reason for not having an indication for (intra)cranial surgery. | 0=No surgical lesion;1=Lesion present, but Acceptable/good neurologic condition;2=Lesion present, but Guideline adherence;3=Lesion present, but Little/no mass effect;4=Lesion present, but Not hospital policy;5=Lesion present, but Extremely poor prognosis;6=Lesion present, but Brain Death;7=Lesion present, but Old age;8=Lesion present, but Wish family;88=Unknown;99=Lesion present, but Other |
| ERCTNoOpMotivOther | TRUE | text | In emergency room: Specification, only applicable if "CTMRI.CTNoOpMotiv" was "other" | Not Applicable |
| ERCTYesOpMotiv | TRUE | integer | In emergency room: WHY question: documents reason for having an indication for (intra)cranial surgery. | 1=Emergency/life saving;2=Clinical deterioration;3=Mass effect on CT;4=Radiological progression;5=(Suspicion of) raised ICP;6=Guideline adherence;7=To prevent deterioration;8=Depressed skull fracture;99=Other |
| ERCTYesOpMotivOther | TRUE | text | In emergency room: Free text if "CTMRI.CTYesOpMotiv" was marked as 'Other'. Relates to the WHY question: documents reason for having an indication for (intra)cranial surgery. | Not Applicable |
| ShortTermSurvivalNoSurg | FALSE | integer | Aims to capture information on the surgeon's expectations, eg if the surgeon considers a realistic expectation of benefit, or performs the surgery as a "last resort" in a likely hopeless case --> The short term survival chances of the patient if I DO NOT operate (1-100) | Not Applicable |
| ShortTermSurvivalYesSurg | FALSE | integer | Aims to capture information on the surgeon's expectations, eg if the surgeon considers a realistic expectation of benefit, or performs the surgery as a "last resort" in a likely hopeless case --> The short term survival chances of the patient if I DO operate (1-100) | Not Applicable |
| SurgeryCranialReason | FALSE | integer | WHY Question: aims to document the reason for intracranial surgery | 1=Emergency/Life saving;2=Clinical deterioration;3=Mass effect on CT;4=Radiological progression;5=suspicion of) raised ICP;6=Guideline adherence;7=To prevent deterioration |
| SurgeryExtraCranialReason | FALSE | integer | The extra-cranial surgeries information was to be entered in the e-CRF in tables for which you could add as many rows as you wish. The tables consisted of - Surgery start date - Surgery start time - Surgery end date - Surgery end time - Extracranial surgery code - Reason - Delay - Short time survival if you do not operate - Short time survival if you do operate | 1=Emergency/Lifesaving;2=Elective;3=Treatment of complication;4=Airway management;99=Other |
| TransReason | FALSE | integer | WHY question: documents reason for transition of care | 1=Mechanical ventilation;2=Frequent neurological observations;3=Haemodynamic invasive monitoring;4=Extracranial injuries;5=Neurological operation;6=Clinical deterioration;7=CT abnormalities;8=Clinical observation for TBI;9=No ICU bed available;10=Could be discharged home, but no adequate supervision;11=Improvement;12=Neurological deterioration;13=Systemic compilation;14=CT progression;15=Planned surgery;16=Condition stable;17=(acute) Treatment goals accomplished;18=Need to free a bed;19=Further improvement;20=Clinical rehab completed;21=Lack of improvement;22=Late neurological deterioration;23=Problems unrelated to trauma;24=Post operative care;25=Neurological complication;99=Other |

### SUPPLEMENTARY METHODS

#### Supplementary Methods S1. Description of model endpoints and outputs for TILTomorrow.

Let $\mathbf{y}^{(i)}$ represent the vector of next-day TIL$^{(\text{Basic})}$ scores for a patient, represented by index $i \in \{1, 2, \dots, N\}$, in an assessment population of $N$ patients:

$$\mathbf{y}^{(i)} = \left[y_1^{(i)}, y_2^{(i)}, \dots, y_{\mathcal{T}^{(i)}}^{(i)}\right]^{\mathsf{T}}$$

where $y_t^{(i)} \in \{0, 1, 2, 3, 4\}$ is the next-day TIL$^{(\text{Basic})}$ score (Table 1) at day $t \in \{1, 2, \dots, \mathcal{T}^{(i)}\}$. In other words, $y_t^{(i)}$ is the TIL$^{(\text{Basic})}$ score at day $t+1$, and $\mathcal{T}^{(i)}+1$ is the number of calendar days patient $i$ was in the ICU. In the CENTER-TBI study, $y_t^{(i)}$ was regularly recorded for $t \in \{1, 2, 3, 4, 5, 6, 9, 13, 20, 27\} \cap \{1,2, \dots, \mathcal{T}^{(i)}\}$. The softmax output layer of the TILTomorrow models returns a trajectory of estimated probabilities $\left(p_{k,t}^{(i)}\right)$ for each possible score ($k \in \{0,1,2,3,4\}$) of next-day TIL$^{(\text{Basic})}$:

$$p_{k,t}^{(i)} = \widehat{\Pr}\left(y_t^{(i)} = k\right).$$

From score-specific probability scores, we calculated two interpretable probability scores. The first was an estimated probability at each possible threshold $\left(p_{>k,t}^{(i)}\right)$ of next-day TIL$^{(\text{Basic})}$:

$$p_{>k,t}^{(i)} = \sum_{k'=k+1}^{4} p_{k',t}^{(i)}$$

$\forall k \in \{0, 1, 2, 3\}$. The second was the probability of TIL$^{(\text{Basic})}$ decreasing $\left(\pi_{-1,t}^{(i)}\right)$, staying the same $\left(\pi_{0,t}^{(i)}\right)$, or increasing $\left(\pi_{1,t}^{(i)}\right)$ tomorrow in relation to the last available TIL$^{(\text{Basic})}$ score. Let $y_0^{(i)}$ represent the TIL$^{(\text{Basic})}$ score of the first calendar day of a patient's ICU stay. Moreover, if $y_{t-1}^{(i)}$ (i.e., today's TIL$^{(\text{Basic})}$ score) is missing, let it be replaced with the last available TIL$^{(\text{Basic})}$ score for the following formulae. Then, $\pi_{-1,t}^{(i)}$ is defined as:

$$\pi_{-1,t}^{(i)} = \begin{cases} 0 & \text{if } y_{t-1}^{(i)} = 0, \\ \displaystyle\sum_{k'=0}^{y_{t-1}^{(i)}-1} p_{k',t}^{(i)} & \text{otherwise.} \end{cases}$$

$\pi_{0,t}^{(i)}$ is defined as:

$$\pi_{0,t}^{(i)} = p_{y_{t-1}^{(i)},t}^{(i)}.$$

$\pi_{1,t}^{(i)}$ is defined as:

$$\pi_{1,t}^{(i)} = \begin{cases} 0 & \text{if } y_{t-1}^{(i)} = 4, \\ \displaystyle\sum_{k'=y_{t-1}^{(i)}+1}^{4} p_{k',t}^{(i)} & \text{otherwise.} \end{cases}$$

Moreover, let $\gamma_t^{(i)} \in \{-1, 0, 1\}$ be the corresponding endpoint label that represents whether the next-day TIL$^{(\text{Basic})}$ score is a decrease, stasis, or increase from the last available TIL$^{(\text{Basic})}$ score:

$$\gamma_t^{(i)} = \begin{cases} -1 & \text{if } y_{t-1}^{(i)} > y_t^{(i)}, \\ 0 & \text{if } y_{t-1}^{(i)} = y_t^{(i)}, \\ 1 & \text{if } y_{t-1}^{(i)} < y_t^{(i)}, \end{cases}$$

and:

$$\boldsymbol{\gamma}^{(i)} = \left[\gamma_1^{(i)}, \gamma_2^{(i)}, \dots, \gamma_{\mathcal{T}^{(i)}}^{(i)}\right]^{\mathsf{T}}.$$

*Post-processing calibration*

Once model weights were trained, we used vector scaling to improve the calibration (i.e., reliability) of estimated probability scores based on the validation sets. Post-processing calibration methods, including vector scaling, are described in greater detail by Guo *et al.*[R1]

The motivation behind vector scaling is to find a single linear transformation of uncalibrated logits which helps account for the effect of over-fitting on the training set. Let $\mathbf{q}_t^{(i)}$ represent the $5 \times 1$ vector of uncalibrated logits for patient $i$ at day $t$, and let $\sigma_{\text{SM}}$ represent the softmax function:

$$p_{k,t}^{(i)} = \sigma_{\text{SM}}\left(\mathbf{W}_t \mathbf{q}_t^{(i)} + \mathbf{b}_t\right)$$

where $\mathbf{W}_t \in \mathbb{R}^{5\times5}$ is fixed as a diagonal matrix and $\mathbf{b}_t \in \mathbb{R}^{5\times1}$. $\mathbf{W}_t$ and $\mathbf{b}_t$ are learned by training a multinomial logistic regression model between uncalibrated logits and next-day $\text{TIL}^{(\text{Basic})}$ scores on the validation set at each assessment day $t$.

#### Supplementary Methods S2. Repeated Bootstrap Bias Corrected with Dropping Cross-Validation (BBCD-CV).

To make the tuning and assessment of our hyperparametric modelling strategy computationally tractable, we implemented a slightly modified version of the Repeated Bootstrap Bias Corrected with Dropping Cross-Validation (BBCD-CV) method proposed by Tsamardinos *et al*.[R2] This method has been reported to achieve similar bias performance to nested CV with considerably greater efficiency.[R2,3]

*Dropout of low-performing hyperparametric configurations*
One of the challenges in training our modelling strategy is the high number of hyperparameter combinations (i.e., configurations). The intuition behind BBCD-CV is to dropout significantly low-performing configurations, determined by bias-corrected bootstrapping of validation set performance, at certain checkpoints of the repeated CV process to make training more efficient.

Let $\Theta = \{\boldsymbol{\theta}_1, \boldsymbol{\theta}_2, \dots, \boldsymbol{\theta}_C\}$ denote the set of configurations. After training models of each of the $C$ configurations on the training sets of the first full repeat (i.e., first five partitions), we collected all the validation set model outputs (i.e., predictions). On these outputs, we calculated two performance metrics for each of the configurations: the ordinal $c$-index[R4] (ORC) for discrimination as well as the macro-averaged calibration slope[R5] ($\overline{\beta_1}$) for calibration. For each metric, we selected the configuration with the optimal performance:

$$i^*_{\mathrm{ORC}} = \arg\max_i \mathrm{ORC}\,(\boldsymbol{\theta}_i)$$
$$i^*_{\overline{\beta_1}} = \arg\min_i \left|1 - \overline{\beta_1}(\boldsymbol{\theta}_i)\right|$$

Then, we drew 1,000 resamples of unique patients from the validation set outputs for bootstrapping and calculate the ORC and $\overline{\beta_1}$ values of each $\boldsymbol{\theta}$ in each resample. For each $\boldsymbol{\theta}_i$, we calculated the proportion of resamples in which $\boldsymbol{\theta}_i$ had a lower ORC than that of $\boldsymbol{\theta}_{i^*_{\mathrm{ORC}}}$ as well as the proportion of resamples in which $\boldsymbol{\theta}_i$ had a higher calibration slope error $\left(\left|1 - \overline{\beta_1}(\boldsymbol{\theta}_i)\right|\right)$ than that of $\boldsymbol{\theta}_{i^*_{\overline{\beta_1}}}$. Moreover, we estimated a 95% confidence interval (CI) of $\overline{\beta_1}$ for each $\boldsymbol{\theta}_i$ based on the configuration's 2.5th and 97.5th percentile of $\overline{\beta_1}$ values across the resamples. If a configuration's proportion of lower-performing resamples was greater than 0.99 for either metric and its 95% CI of $\overline{\beta_1}$ did not include 1, then that configuration was dropped from further training or assessment.

We repeated this process after each full repeat (i.e., every five partitions), until 20 or fewer configurations remained. After training was complete on all 100 repeated CV partitions, we repeated the dropout procedure one last time to remove configurations from testing set assessment.

*Confidence intervals for testing set performance*
After model training and configuration dropout was complete, we assessed the performance of our modelling strategies with bias-corrected bootstrapping. We compiled the set of testing set outputs for the remaining configurations and drew 1,000 resamples of unique patients in the population for bootstrapping. We iterated through each of the resamples and determined the optimal configuration for each performance metric in the current resample. Then, we calculated the corresponding performance metric for the optimal configuration in the set of patients not in the current resample. The collection of 1,000 out-of-sample performance metric values formed the estimated distribution of the metric for statistical inference, from which the 2.5th and 97.5th percentiles formed the bounds for the metric's 95% confidence interval.

It is important to note that repeated CV assesses the performance of a modelling strategy and not the performance of a specific trained model or a specific hyperparametric configuration. The modelling strategy encompasses the full range of tested configurations, and the optimal configuration for a given metric may differ between resamples. Moreover, by choosing the optimal configuration within one set of patients and then assessing its performance in another for each resample, the BBCD-CV algorithm reduces the bias in configuration selection without needing to train additional models.

#### Supplementary Methods S3. Hyperparameter optimisation report.

*Summary*

Combinations of the listed hyperparameters were tested on the validation sets of our repeated *k*-fold cross-validation (20 repeats, 5 folds) in successive model versions. A single combination of model hyperparameters is known as a configuration. Configurations which significantly ($\alpha = 0.01$) underperformed in calibration and discrimination on the validation set were dropped out after each repeat using the Bootstrap Bias Corrected with Dropping Cross-Validation (BBCD-CV) method, as detailed in Supplementary Methods S2. For greater detail regarding the role of each hyperparameter in model function, please see the model code in https://github.com/sbhattacharyay/TILTomorrow/blob/main/scripts/models/dynamic_TTM.py. Moreover the selection of hyperparameters in this study was informed by the optimal configurations of our prior, dynamic GOSE modelling study.[R6]

*Overview of tested hyperparameters*

- Embedding vector dimension: length of vectors learned for each token in the embedding layer.
  - Tested values: 128, 256, 512, 1024
  - Optimal value: 512
- Recurrent neural network (RNN) architecture: type of RNN structure.
  - Tested values: long short-term memory (LSTM), gated recurrent unit (GRU)
  - Optimal value: GRU
- RNN hidden state dimension: dimension of the RNN hidden state.
  - Tested values: 128, 256, 512
  - Optimal value: 256
- Window limit during training: limit to the number of time windows per training set patient considered during training.
  - Tested values: None, 6, 13
  - Optimal value: 13
- Minimum variable representation: minimum proportion of patients with non-missing value for a variable for it to be included in the model embedding layer dictionary.
  - Tested values: None, 0.05
  - Optimal value: None
- Maximum number of tokens: maximum number of tokens a single variable can have for it to be included in the embedding layer dictionary.
  - Tested values: None, 100
  - Optimal value: None

*Tested hyperparameters per model version*

We had two iterations of model development. Attached are the high-dimensional parallel plots (HiPlots) to visualise the effect of hyperparameters on the validation set ordinal $c$-index (ORC) and calibration slope error $\left(\left|1 - \overline{\beta_1}\right|\right)$.

Version 1-0:

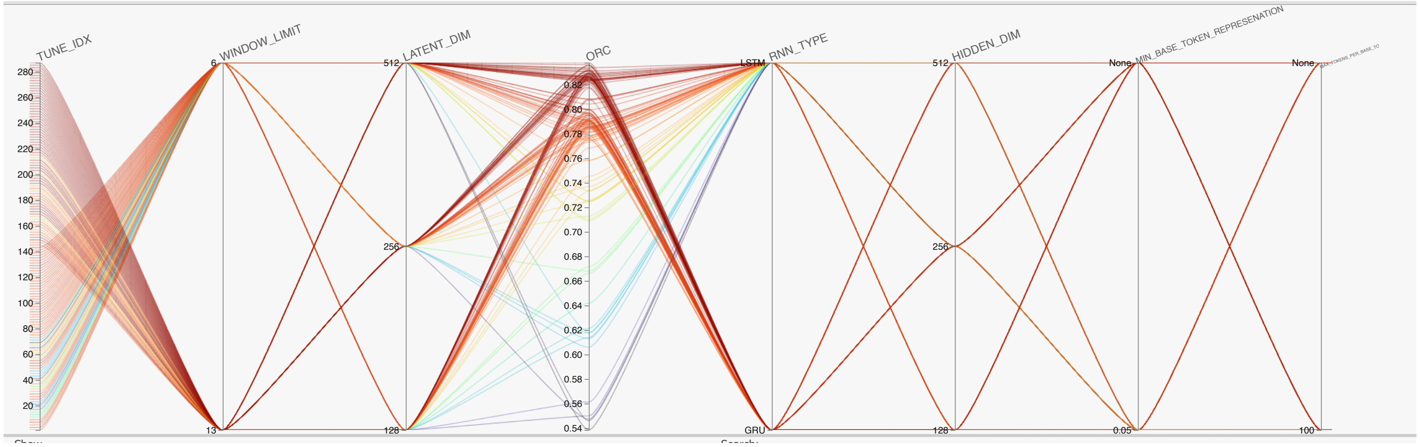


*Version 1-0 HiPlots of ORC*. An interactive version of the HiPlot is available on GitHub: https://sbhattacharyay.github.io/TILTomorrow/TILTomorrow_model_performance/v1-0/ORC_hiplot.html.

| | TUNE_IDX | WINDOW_LIMIT | RNN_TYPE | LATENT_DIM | HIDDEN_DIM | MIN_BASE_TOKEN_REPRESENATION | MAX_TOKENS_PER_BASE_TOKEN | ORC | uid | from_uid |
|---|---|---|---|---|---|---|---|---|---|---|
| ■ | 277 | 13 | GRU | 512 | 256 | 0.05 | None | 0.8379508363195278 | 0 | null |
| ■ | 265 | 13 | GRU | 512 | 128 | None | None | 0.8362549909194259 | 1 | null |
| ■ | 245 | 13 | GRU | 256 | 128 | 0.05 | None | 0.8357516317744974 | 2 | null |
| ■ | 259 | 13 | GRU | 256 | 512 | None | 100 | 0.8355192502123274 | 3 | null |
| ■ | 207 | 13 | LSTM | 512 | 256 | 0.05 | 100 | 0.833763214970582 | 4 | null |
| ■ | 279 | 13 | GRU | 512 | 256 | 0.05 | 100 | 0.8332464485253526 | 5 | null |
| ■ | 283 | 13 | GRU | 512 | 512 | None | 100 | 0.8322209465823152 | 6 | null |
| ■ | 253 | 13 | GRU | 256 | 256 | 0.05 | None | 0.8317453677058259 | 7 | null |
| ■ | 243 | 13 | GRU | 256 | 128 | None | 100 | 0.8316034788036919 | 8 | null |
| ■ | 249 | 13 | GRU | 256 | 256 | None | None | 0.8313614775903667 | 9 | null |
| ■ | 211 | 13 | LSTM | 512 | 512 | None | 100 | 0.8309341709083858 | 10 | null |
| ■ | 285 | 13 | GRU | 512 | 512 | 0.05 | None | 0.8304756587307744 | 11 | null |
| ■ | 227 | 13 | GRU | 128 | 256 | None | 100 | 0.8296171243455466 | 12 | null |
| ■ | 261 | 13 | GRU | 256 | 512 | 0.05 | None | 0.8292864544753716 | 13 | null |
| ■ | 171 | 13 | LSTM | 256 | 128 | None | 100 | 0.8292560103508316 | 14 | null |
| ■ | 273 | 13 | GRU | 512 | 256 | None | None | 0.8290438050713731 | 15 | null |
| ■ | 237 | 13 | GRU | 128 | 512 | 0.05 | None | 0.8287387921356838 | 16 | null |
| ■ | 233 | 13 | GRU | 128 | 512 | None | None | 0.8285117441289452 | 17 | null |
| ■ | 149 | 13 | LSTM | 128 | 128 | 0.05 | None | 0.8284087633698562 | 18 | null |
| ■ | 203 | 13 | LSTM | 512 | 256 | None | 100 | 0.8276074143996436 | 19 | null |
| ■ | 247 | 13 | GRU | 256 | 128 | 0.05 | 100 | 0.8274887803012904 | 20 | null |
| ■ | 251 | 13 | GRU | 256 | 256 | None | 100 | 0.8270750403167227 | 21 | null |
| ■ | 287 | 13 | GRU | 512 | 512 | 0.05 | 100 | 0.8268932274408687 | 22 | null |
| ■ | 195 | 13 | LSTM | 512 | 128 | None | 100 | 0.826596759454209 | 23 | null |
| ■ | 271 | 13 | GRU | 512 | 128 | 0.05 | 100 | 0.8264529090181327 | 24 | null |

*Version 1-0 top 25 hyperparametric configurations based on ORC*. An interactive version of this chart is available on GitHub: https://sbhattacharyay.github.io/TILTomorrow/TILTomorrow_model_performance/v1-0/ORC_hiplot.html.

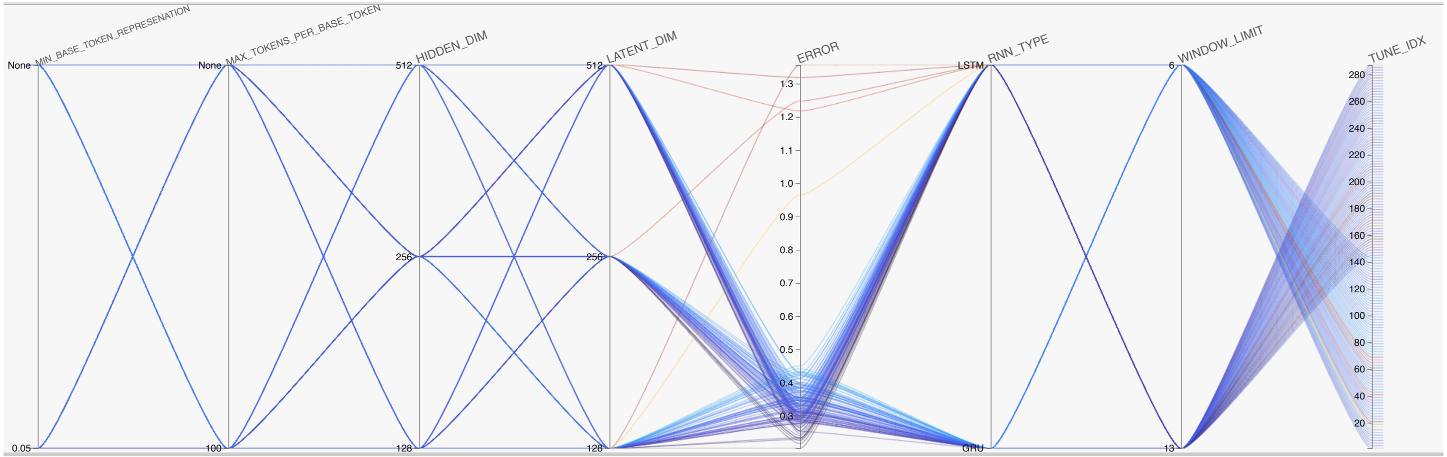


*Version 1-0 HiPlots of macro-averaged calibration slope error.* An interactive version of the HiPlot is available on GitHub: https://sbhattacharyay.github.io/TILTomorrow/TILTomorrow_model_performance/v1-0/thresh_calibration_hiplot.html.

| TUNE_IDX | WINDOW_LIMIT | RNN_TYPE | LATENT_DIM | HIDDEN_DIM | MIN_BASE_TOKEN_REPRESENATION | MAX_TOKENS_PER_BASE_TOKEN | ERROR |
|---|---|---|---|---|---|---|---|
| 157 | 13 | LSTM | 128 | 256 | 0.05 | None | 0.2029465656318122 |
| 191 | 13 | LSTM | 256 | 512 | 0.05 | 100 | 0.21724159025475107 |
| 171 | 13 | LSTM | 256 | 128 | None | 100 | 0.2264563378255279 |
| 155 | 13 | LSTM | 128 | 256 | None | 100 | 0.23001757761345892 |
| 187 | 13 | LSTM | 256 | 512 | None | 100 | 0.23400563938119193 |
| 189 | 13 | LSTM | 256 | 512 | 0.05 | None | 0.2341144599076415 |
| 149 | 13 | LSTM | 128 | 128 | 0.05 | None | 0.2349617742179136 |
| 175 | 13 | LSTM | 256 | 128 | 0.05 | 100 | 0.24222775043586983 |
| 61 | 6 | LSTM | 512 | 256 | 0.05 | None | 0.24819935079851194 |
| 277 | 13 | GRU | 512 | 256 | 0.05 | None | 0.25415229742201695 |
| 207 | 13 | LSTM | 512 | 256 | 0.05 | 100 | 0.25565114547886836 |
| 209 | 13 | LSTM | 512 | 512 | None | None | 0.2565276979970379 |
| 211 | 13 | LSTM | 512 | 512 | None | 100 | 0.2652089646139199 |
| 183 | 13 | LSTM | 256 | 256 | 0.05 | 100 | 0.26772146475502395 |
| 169 | 13 | LSTM | 256 | 128 | None | None | 0.2714102630505819 |
| 199 | 13 | LSTM | 512 | 128 | 0.05 | 100 | 0.2756491254201819 |
| 7 | 6 | LSTM | 128 | 128 | 0.05 | 100 | 0.2774463304798968 |
| 167 | 13 | LSTM | 128 | 512 | 0.05 | 100 | 0.27757104033660013 |
| 165 | 13 | LSTM | 128 | 512 | 0.05 | None | 0.27835888742558573 |
| 245 | 13 | GRU | 256 | 128 | 0.05 | None | 0.28036300312348 |
| 227 | 13 | GRU | 128 | 256 | None | 100 | 0.2830082783141855 |
| 285 | 13 | GRU | 512 | 512 | 0.05 | None | 0.2833660281970304 |
| 263 | 13 | GRU | 256 | 512 | 0.05 | 100 | 0.28423038020215674 |
| 213 | 13 | LSTM | 512 | 512 | 0.05 | None | 0.2857221826161927 |
| 261 | 13 | GRU | 256 | 512 | 0.05 | None | 0.2882236051565642 |

*Version 1-0 top 25 hyperparametric configurations based on macro-averaged calibration slope error.* An interactive version of this chart is available on GitHub: [https://sbhattacharyay.github.io/TILTomorrow/TILTomorrow_model_performance/v1-0/thresh_calibration_hiplot.html](https://sbhattacharyay.github.io/TILTomorrow/TILTomorrow_model_performance/v1-0/thresh_calibration_hiplot.html).

<u>Version 2-0</u>:

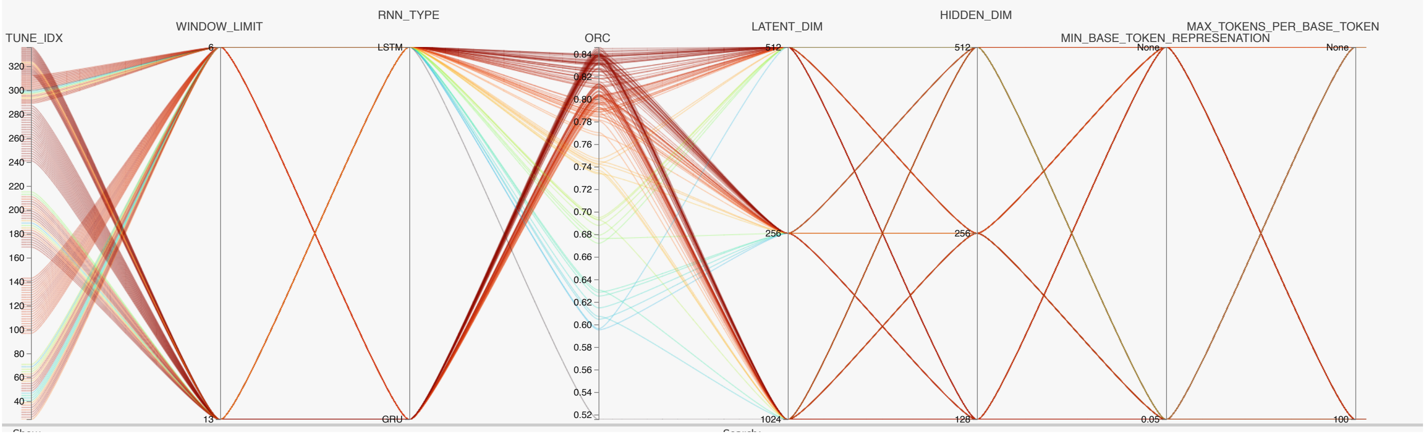


*Version 2-0 HiPlots of ORC*. An interactive version of the HiPlot is available on GitHub: [https://sbhattacharyay.github.io/TILTomorrow/TILTomorrow_model_performance/v2-0/ORC_hiplot.html](https://sbhattacharyay.github.io/TILTomorrow/TILTomorrow_model_performance/v2-0/ORC_hiplot.html).

| | TUNE_IDX | WINDOW_LIMIT | RNN_TYPE | LATENT_DIM | HIDDEN_DIM | MIN_BASE_TOKEN_REPRESENATION | MAX_TOKENS_PER_BASE_TOKEN | ORC |
|---|---|---|---|---|---|---|---|---|
| ■ | 333 | 13 | GRU | 1024 | 512 | None | None | 0.8460361262840276 |
| ■ | 269 | 13 | GRU | 512 | 128 | 0.05 | None | 0.844614332489145 |
| ■ | 336 | 13 | GRU | 1024 | 512 | 0.05 | 100 | 0.8441096763652861 |
| ■ | 241 | 13 | GRU | 256 | 128 | None | None | 0.8428358220706831 |
| ■ | 265 | 13 | GRU | 512 | 128 | None | None | 0.8426742510973863 |
| ■ | 281 | 13 | GRU | 512 | 512 | None | None | 0.84168484650292 |
| ■ | 330 | 13 | GRU | 1024 | 256 | None | 100 | 0.8414519746724569 |
| ■ | 325 | 13 | GRU | 1024 | 128 | None | None | 0.8408002699568409 |
| ■ | 332 | 13 | GRU | 1024 | 256 | 0.05 | 100 | 0.8402506566109267 |
| ■ | 247 | 13 | GRU | 256 | 128 | 0.05 | 100 | 0.8399068920069712 |
| ■ | 283 | 13 | GRU | 512 | 512 | None | 100 | 0.8398277297507862 |
| ■ | 287 | 13 | GRU | 512 | 512 | 0.05 | 100 | 0.8397874373011506 |
| ■ | 326 | 13 | GRU | 1024 | 128 | None | 100 | 0.839512964715698 |
| ■ | 316 | 13 | LSTM | 1024 | 128 | 0.05 | 100 | 0.8394968128621728 |
| ■ | 251 | 13 | GRU | 256 | 256 | None | 100 | 0.8394796453697052 |
| ■ | 175 | 13 | LSTM | 256 | 128 | 0.05 | 100 | 0.8393192154613309 |
| ■ | 329 | 13 | GRU | 1024 | 256 | None | None | 0.8390900349902727 |
| ■ | 328 | 13 | GRU | 1024 | 128 | 0.05 | 100 | 0.839054413867727 |
| ■ | 263 | 13 | GRU | 256 | 512 | 0.05 | 100 | 0.8389797170280731 |
| ■ | 199 | 13 | LSTM | 512 | 128 | 0.05 | 100 | 0.838605991353371 |
| ■ | 320 | 13 | LSTM | 1024 | 256 | 0.05 | 100 | 0.8382258141295637 |
| ■ | 334 | 13 | GRU | 1024 | 512 | None | 100 | 0.838140482303067 |
| ■ | 279 | 13 | GRU | 512 | 256 | 0.05 | 100 | 0.8381249583390414 |
| ■ | 331 | 13 | GRU | 1024 | 256 | 0.05 | None | 0.8379533310023022 |
| ■ | 267 | 13 | GRU | 512 | 128 | None | 100 | 0.837781993880267 |

*Version 2-0 top 25 hyperparametric configurations based on ORC*. An interactive version of this chart is available on GitHub: https://sbhattacharyay.github.io/TILTomorrow/TILTomorrow_model_performance/v2-0/ORC_hiplot.html.

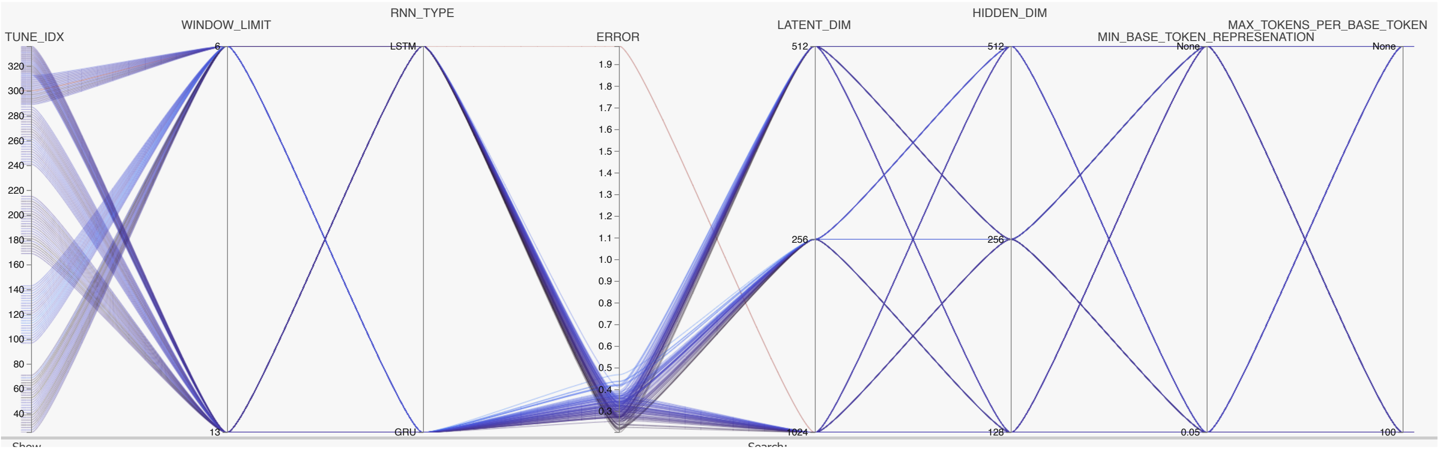


*Version 2-0 HiPlots of macro-averaged calibration slope error.* An interactive version of the HiPlot is available on GitHub: https://sbhattacharyay.github.io/TILTomorrow/TILTomorrow_model_performance/v2-0/thresh_calibration_hiplot.html.

| | TUNE_IDX | WINDOW_LIMIT | RNN_TYPE | LATENT_DIM | HIDDEN_DIM | MIN_BASE_TOKEN_REPRESENATION | MAX_TOKENS_PER_BASE_TOKEN | ERROR |
|---|---|---|---|---|---|---|---|---|
| ■ | 53 | 6 | LSTM | 512 | 128 | 0.05 | None | 0.2027243284878944 |
| ■ | 65 | 6 | LSTM | 512 | 512 | None | None | 0.212854050669090 3 |
| ■ | 39 | 6 | LSTM | 256 | 256 | 0.05 | 100 | 0.21734548483198873 |
| ■ | 181 | 13 | LSTM | 256 | 256 | 0.05 | None | 0.2190650489945436 |
| ■ | 63 | 6 | LSTM | 512 | 256 | 0.05 | 100 | 0.22024893048456684 |
| ■ | 177 | 13 | LSTM | 256 | 256 | None | None | 0.22203808385460866 |
| ■ | 31 | 6 | LSTM | 256 | 128 | 0.05 | 100 | 0.2264820278197573 |
| ■ | 320 | 13 | LSTM | 1024 | 256 | 0.05 | 100 | 0.22667236901330745 |
| ■ | 211 | 13 | LSTM | 512 | 512 | None | 100 | 0.23626360318497205 |
| ■ | 297 | 6 | LSTM | 1024 | 512 | None | None | 0.23730447320492137 |
| ■ | 175 | 13 | LSTM | 256 | 128 | 0.05 | 100 | 0.237418278366817 |
| ■ | 294 | 6 | LSTM | 1024 | 256 | None | 100 | 0.24011129666603126 |
| ■ | 67 | 6 | LSTM | 512 | 512 | None | 100 | 0.2434552211485323 |
| ■ | 35 | 6 | LSTM | 256 | 256 | None | 100 | 0.2499676164652886 |
| ■ | 191 | 13 | LSTM | 256 | 512 | 0.05 | 100 | 0.2516177026768438 |
| ■ | 267 | 13 | GRU | 512 | 128 | None | 100 | 0.2541600858933404 |
| ■ | 201 | 13 | LSTM | 512 | 256 | None | None | 0.2557812950950744 |
| ■ | 316 | 13 | LSTM | 1024 | 128 | 0.05 | 100 | 0.25905303624606385 |
| ■ | 183 | 13 | LSTM | 256 | 256 | 0.05 | 100 | 0.2599552967513904 |
| ■ | 33 | 6 | LSTM | 256 | 256 | None | None | 0.2641243560257474 |
| ■ | 57 | 6 | LSTM | 512 | 256 | None | None | 0.2657096651743324 |
| ■ | 291 | 6 | LSTM | 1024 | 128 | 0.05 | None | 0.2657932320234061 |
| ■ | 318 | 13 | LSTM | 1024 | 256 | None | 100 | 0.2683017451805711 |
| ■ | 207 | 13 | LSTM | 512 | 256 | 0.05 | 100 | 0.26929298783368893 |
| ■ | 71 | 6 | LSTM | 512 | 512 | 0.05 | 100 | 0.271057399569399 |

*Version 2-0 top 25 hyperparametric configurations based on macro-averaged calibration slope error.* An interactive version of this chart is available on GitHub: [https://sbhattacharyay.github.io/TILTomorrow/TILTomorrow_model_performance/v2-0/thresh_calibration_hiplot.html](https://sbhattacharyay.github.io/TILTomorrow/TILTomorrow_model_performance/v2-0/thresh_calibration_hiplot.html).

#### Supplementary Methods S4. Calculation of Somers' $D_{xy}$.

Somers' $D_{xy}$, as proposed by Somers[R7] and Kim,[R8] is used as the primary metric for quantifying uncertainty in terms of explanation of the ordinal variation in next-day changes in TIL(Basic) from the variables in the CENTER-TBI dataset.

Carrying over the notation defined in Supplementary Methods S1, let us define $\epsilon_t^{(i)}$ as:

$$\epsilon_t^{(i)} = \sum_{l \in \{-1,0,1\}} l \cdot \pi_{l,t}^{(i)}$$

which corresponds to the expected direction of change in next-day TIL(Basic) from the last available score. At each of the days of performance assessment $(\text{i.e.}, \forall t \in \{1,2,3,4,5,6,9,13\})$, the $\epsilon_t$ scores and the $\gamma_t$ labels from across the assessment population are compiled into vectors:

$$\boldsymbol{\epsilon}_t = \left[\epsilon_t^{(1)}, \epsilon_t^{(2)}, \dots, \epsilon_t^{(N)}\right]^{\mathsf{T}}$$
$$\boldsymbol{\gamma}_t = \left[\gamma_t^{(1)}, \gamma_t^{(2)}, \dots, \gamma_t^{(N)}\right]^{\mathsf{T}}.$$

Somers' $D_{xy}$ is defined by:

$$D_{xy,t} = \frac{\tau(\boldsymbol{\gamma}_t, \boldsymbol{\epsilon}_t)}{\tau(\boldsymbol{\gamma}_t, \boldsymbol{\gamma}_t)}$$

where $\tau$ is the Kendall's $\tau$ coefficient, defined for any two vectors $\mathbf{a}$ and $\mathbf{b}$:

$$\tau(\mathbf{b}, \mathbf{a}) = \frac{n_C(\mathbf{b}, \mathbf{a}) - n_D(\mathbf{b}, \mathbf{a})}{\binom{n}{2}}$$

where $n$ is the length of $\mathbf{a}$ or $\mathbf{b}$, and $n_C(\mathbf{b}, \mathbf{a})$ is the number of concordant pairs between $\mathbf{a}$ and $\mathbf{b}$ and $n_D(\mathbf{b}, \mathbf{a})$ is the number of discordant pairs between $\mathbf{a}$ and $\mathbf{b}$.

Pairs between two vectors are concordant if both elements of the pair agree in rank. Between vectors $\boldsymbol{\gamma}_t$ and $\boldsymbol{\epsilon}_t$, a pair of patients $\{i,j\}$ is concordant if either $\epsilon_t^{(i)} > \epsilon_t^{(j)}$ and $\gamma_t^{(i)} > \gamma_t^{(j)}$ or $\epsilon_t^{(i)} < \epsilon_t^{(j)}$ and $\gamma_t^{(i)} < \gamma_t^{(j)}$. Between the vector $\boldsymbol{\gamma}_t$ and itself, a pair of patients $\{i,j\}$ is concordant if they have different endpoint classes. Pairs between two vectors are discordant if either element of the pair disagrees in rank. Between vectors $\boldsymbol{\gamma}_t$ and $\boldsymbol{\epsilon}_t$, a pair of patients $\{i,j\}$ is discordant if either $\epsilon_t^{(i)} > \epsilon_t^{(j)}$ and $\gamma_t^{(i)} < \gamma_t^{(j)}$ or $\epsilon_t^{(i)} < \epsilon_t^{(j)}$ and $\gamma_t^{(i)} > \gamma_t^{(j)}$. Between the vector $\boldsymbol{\gamma}_t$ and itself, there are no pairs that are discordant. Therefore, $\tau(\boldsymbol{\gamma}_t, \boldsymbol{\gamma}_t)$ is equivalent to the proportion of possible pairs of patients in the assessment population that have different endpoint classes at day $t$. This is considered a measure of the ordinal variation in the endpoint.[R4]

Let $n^{(\text{conc})}$ denote the number of concordant pairs between $\boldsymbol{\gamma}_t$ and $\boldsymbol{\epsilon}_t$, and let $n^{(\text{disc})}$ denote the number of discordant pairs between $\boldsymbol{\gamma}_t$ and $\boldsymbol{\epsilon}_t$. Let $n^{(\text{comp})}$ denote the number of pairs of patients within the assessment population with different endpoint classes (i.e., comparable pairs). The formula for Somers' $D_{xy}$ can then be simplified to:

$$D_{xy,t} = \frac{n_C(\boldsymbol{\gamma}_t, \boldsymbol{\epsilon}_t) - n_D(\boldsymbol{\gamma}_t, \boldsymbol{\epsilon}_t)}{n_C(\boldsymbol{\gamma}_t, \boldsymbol{\gamma}_t)}$$
$$= \frac{n^{(\text{conc})} - n^{(\text{disc})}}{n^{(\text{comp})}}.$$

Somers' $D_{xy}$ equals the ratio of the difference between the number of concordant pairs and number of discordant pairs to the total number of comparable pairs. Assuming there are no ties in $\epsilon_t^{(i)}$ between patients of different $\gamma_t^{(i)}$,

$$= \frac{n^{(\text{conc})} - \left(n^{(\text{comp})} - n^{(\text{conc})}\right)}{n^{(\text{comp})}}$$
$$= \frac{2n^{(\text{conc})} - n^{(\text{comp})}}{n^{(\text{comp})}}$$
$$= 2\frac{n^{(\text{conc})}}{n^{(\text{comp})}} - 1$$
$$= 2\left[\frac{n^{(\text{conc})}}{\sum_{\text{l}=-1}^{0}\sum_{\text{m}=\text{l}+1}^{1}\left|\Pi_{\text{l},t}\right|\left|\Pi_{\text{m},t}\right|}\right] - 1$$
$$= 2\left[\frac{\sum_{\text{l}'=-1}^{0}\sum_{\text{m}'=\text{l}'+1}^{1}\left|\Pi_{\text{l}',t}\right|\left|\Pi_{\text{m}',t}\right|\text{c}_{\text{l}'\text{m}',t}}{\sum_{l=-1}^{0}\sum_{m=l+1}^{1}\left|\Pi_{l,t}\right|\left|\Pi_{m,t}\right|}\right] - 1$$

where $\Pi_{l,t} \subseteq \{1,2,\dots,N\}$ denotes the subset of indices of patients with $\gamma_t^{(i)} = l$ for each $l \in \{-1,0,1\}$ and $c_{l'm',t}$ denotes the pairwise $c$-index (i.e., area under the receiver operating characteristic curve [AUC]) between patients with $\gamma_t^{(i)} = l'$ and those with $\gamma_t^{(i)} = m'$. In other words, Somers' $D_{xy}$ is equivalent to twice the prevalence-weighted average of pairwise $c$-indices minus one. Therefore, the feasible range of Somers' $D_{xy}$ is 0 (or 0%) to 1 (or 100%). Somers' $D_{xy}$ can also be interpreted as the **proportion of ordinal variation in the endpoint that can be explained by the variation in model output**.

#### Supplementary Methods S5. Explanation of model outputs with Shapley value estimations.

On an individual patient level, we estimated the contribution of specific variables towards trained model outputs with algorithmic approximations of Shapley values. Shapley values, developed originally for cooperative game theory,[R9] distribute a reward (or loss) amongst members of a team based on their positive or negative contributions. Now, suppose we represent a patient's feature values – in our case, tokens – as teammates, and we let the difference between a patient's model output and the average model output be the reward. Then, Shapley values can theoretically provide a window into how the model's output is affected by the values of specific features, regardless of the model's structure.

*Shapley values*

Suppose we have a trained, static version of a TILTomorrow model which only predicts next-day TIL$^{(\text{Basic})}$ on day one of ICU stay. Let $M$ represent the total number of tokens stored in the embedding layer dictionary and let $\mathbf{x}^{(i)} \in \{0,1\}^M$ be a binary vector representing a patient's set of tokens for the first calendar day of ICU stay such that a 1 represents the existence of the corresponding dictionary token in the time window. The Shapley value of a token with index $j \in \{1,2,\ldots,M\}$ where $x_j^{(i)} = 1$ is defined as:

$$\phi_j^{(i)} = \sum_{S \subseteq \{1,2,\ldots,M\} \setminus \{j\}} \frac{|S|!\,(M - |S| - 1)!}{M!} \left( v_{\mathbf{x}^{(i)}}(S \cup \{j\}) - v_{\mathbf{x}^{(i)}}(S) \right)$$

where $S$ is a subset of tokens (i.e., coalition) for which the patient's true values are taken and $v_{\mathbf{x}^{(i)}}(S)$ is a function which calculates the marginal contribution of a coalition towards model output:

$$v_{\mathbf{x}^{(i)}}(S) = \int \ldots \int \hat{f}(\mathbf{x}^{(i)}) d\mathbb{T}_{\setminus S} - \mathbb{E}_{\mathbf{x}}[\hat{f}(\mathbf{x})]$$

where $\mathbb{T}_{\setminus S}$ is the token space excluding tokens in the coalition $S$, $\hat{f}$ is a function that returns the trained model output for a given token set $\mathbf{x}$, and $\mathbb{E}_{\mathbf{x}}[\hat{f}(\mathbf{x})]$ is the average model output. In other words, the Shapley value of a specific token equals its average marginal contribution across all possible coalitions. Coalitions are weighted by size to provide greater influence on a specific token's effect when it is closer to isolation (i.e., $|S| \rightarrow 0$) or the patient's true token set (i.e., $|S| \rightarrow M$). $v_{\mathbf{x}^{(i)}}(S)$ integrates out all the effects of tokens not in the given coalition and subtracts the average model output to return the marginal contribution of the coalition of variables towards model output. In this analysis, the chosen model output for Shapley value estimation is the expected next-day TIL$^{(\text{Basic})}$ score:

$$\omega_t^{(i)} = \sum_{k=0}^{4} k \cdot p_{k,t}^{(i)}$$

with notation defined in Supplementary Methods S1. Shapley values can be interpreted as **a token's contribution to the difference between an individual patient's model output and the population-average model output, given the patient's full set of tokens**.

However, Shapley values pose several practical challenges for our application. Direct Shapley value calculation is infeasible, as it would require iterating through up to $2^M$ (where $M \approx 30{,}000$) coalitions per patient. Moreover, in the sparse latent space of our embedding layer, integration over coalitions of tokens is not trivial. TILTomorrow is a dynamic modelling strategy, and Shapley values would have to be extended into the temporal dimension, further complicating the feasibility of their estimation.

*KernelSHAP*

The SHapley Additive exPlanations (SHAP) method, proposed by Lundberg *et al.*,[R10] has become a popular tool for estimating Shapley values with a linear model. Suppose we have a patient $i$ with binary token vector $\mathbf{x}^{(i)}$. We are interested in understanding how the tokens contribute towards $\omega^{(i)}$, and we designate $\hat{f}: \{0,1\}^{M \times 1} \rightarrow \mathbb{R}_{[0,4]}$ as the trained model function:

$$\hat{f}(\mathbf{x}^{(i)}) = \omega^{(i)}.$$

For this specific case, SHAP intends to learn a linear explanation function $g^{(i)}$ which maps a binary coalition vector $\boldsymbol{\zeta}^{(i)} \in \{0,1\}^{M \times 1}$ – which specifies the elements of $\mathbf{x}^{(i)}$ that are maintained in the coalition – to a value that approximates $\hat{f}(\mathbf{x}^{(i)})$ when $\boldsymbol{\zeta}^{(i)} \approx 1$. Then, $g^{(i)}$ can be represented as:

$$g^{(i)}(\boldsymbol{\zeta}^{(i)}) = \phi_0^{(i)} + \sum_{j=1}^{M} \phi_j^{(i)} \zeta_j^{(i)},$$

i.e., the sum of Shapley values $\phi_j^{(i)}, \forall j \in \{1,2,\dots,M\}$.

Lundberg *et al*. proposed the KernelSHAP algorithm which estimates the Shapley values by sampling coalition vector data and fitting a weighted linear regression model.[R10] First, we need to define a mapping function $h_{\mathbf{x}^{(i)}}(\boldsymbol{\zeta}^{(i)})$ which transforms the coalition assignments from $\boldsymbol{\zeta}^{(i)}$ to the space of $\mathbf{x}^{(i)}$. For our application, this is quite simple, since $\mathbf{x}^{(i)}$ is itself a binary vector:

$$h_{\mathbf{x}^{(i)}}(\boldsymbol{\zeta}^{(i)}) = \mathbf{x}^{(i)} \odot \boldsymbol{\zeta}^{(i)} + (1 - \boldsymbol{\zeta}^{(i)}) \odot \mathbf{b}$$

where $\mathbf{b} \in \{0,1\}^{M \times 1}$ is a baseline vector which replaces each out-of-coalition value in $\mathbf{x}^{(i)}$ with a value from elsewhere. In this work, we used replacement with the mode of that index across the training set. Then, the algorithm samples $Z$ different combinations of $\boldsymbol{\zeta}^{(i)}$ (i.e., coalitions) and calculates $\hat{f}\left(h_{\mathbf{x}^{(i)}}(\boldsymbol{\zeta}^{(i)})\right)$ for each one. In our applications, we constrained coalition sampling so that: (1) only indices corresponding to a token represented in $\mathbf{x}^{(i)}$ could be perturbed, i.e., only sampling from $\left\{j \in \{1,2,\dots,M\}: x_j^{(i)} = 1\right\}$, and (2) sampling would exhaust coalitions of large and small sizes first before working towards middle-size coalitions until $Z$ samples were obtained. This is motivated by the Shapley value equation, which weighs coalitions of small and large sizes more heavily. After all coalitions were sampled and combined into set $\mathcal{Z} = \left\{\boldsymbol{\zeta}_1^{(i)}, \boldsymbol{\zeta}_2^{(i)}, \dots, \boldsymbol{\zeta}_Z^{(i)}\right\}$, Shapley values were estimated by optimising the following loss function:

$$\begin{aligned}\ell^{(i)}(\hat{f}, g^{(i)}, \pi_{\mathbf{x}^{(i)}}) &= \sum_{\boldsymbol{\zeta}_j^{(i)} \in \mathcal{Z}} \left[\hat{f}\left(h_{\mathbf{x}^{(i)}}(\boldsymbol{\zeta}^{(i)})\right) - g^{(i)}(\boldsymbol{\zeta}_j^{(i)})\right]^2 \pi_{\mathbf{x}^{(i)}}(\boldsymbol{\zeta}_j^{(i)}) \\ &= \sum_{\boldsymbol{\zeta}_j^{(i)} \in \mathcal{Z}} \left[\hat{f}\left(h_{\mathbf{x}^{(i)}}(\boldsymbol{\zeta}^{(i)})\right) - \phi^{(i)\top}\boldsymbol{\zeta}_j^{(i)}\right]^2 \pi_{\mathbf{x}^{(i)}}(\boldsymbol{\zeta}_j^{(i)})\end{aligned}$$

where $\pi_{\mathbf{x}^{(i)}}$ is the kernel set to achieve similar weighting as the Shapley equation:

$$\pi_{\mathbf{x}^{(i)}}(\boldsymbol{\zeta}_j^{(i)}) = \frac{(M-1)}{\binom{M}{|\boldsymbol{\zeta}_j^{(i)}|}\left|\boldsymbol{\zeta}_j^{(i)}\right|\left(M - \left|\boldsymbol{\zeta}_j^{(i)}\right|\right)}.$$

*TimeSHAP and ΔTimeSHAP*

TimeSHAP is a temporal extension of the KernelSHAP algorithm proposed by Bento *et al.*[R11] for efficient and multi-level model output explanation. Like several other temporal extensions of KernelSHAP, TimeSHAP estimates the contribution of tokens and time windows before a certain model output. However, TimeSHAP also groups combinations of tokens and time windows in meaningful ways to enhance the feasibility and focus of KernelSHAP. This starts with a temporal coalition pruning algorithm.

<u>Temporal coalition pruning</u>:

TimeSHAP starts by finding a point back in time before which tokens have a negligible effect on the current model output. Let the binary matrix $\mathbf{X}^{(i)} \in \{0,1\}^{M \times \mathcal{T}^{(i)}}$ be the tokenised representation of a patient's ICU record, where each row represents a token in the training set dictionary and each column represents a calendar day in the patient's ICU stay. Suppose we are interested in explaining the output of a trained dynamic model $(\hat{f})$ at the last time window, $\mathcal{T}^{(i)}$. The temporal coalition pruning algorithm first groups all the tokens at $\mathcal{T}^{(i)}$ $\left(\mathbf{X}_{:,\mathcal{T}^{(i)}}^{(i)}\right)$ as one "feature" and groups all the tokens from time $\{1,2,\dots,\mathcal{T}^{(i)} - 1\}$ $\left(\mathbf{X}_{:,1:\mathcal{T}^{(i)}-1}^{(i)}\right)$ as another feature, and runs KernelSHAP on just these two features $(2^2 = 4 \text{ total coalitions})$. Then, the algorithm pushes back one step in time, groups tokens from $\{\mathcal{T}^{(i)} - 1, \mathcal{T}^{(i)}\}$ and $\{1,2,\dots,\mathcal{T}^{(i)} - 2\}$ into two separate features $\left(\mathbf{X}_{:,\mathcal{T}^{(i)}-1:\mathcal{T}^{(i)}}^{(i)} \text{ and } \mathbf{X}_{:,1:\mathcal{T}^{(i)}-2}^{(i)}\right)$, and runs KernelSHAP again. This process is iteratively repeated, pushing back one step at a time, until the estimated Shapley value corresponding to the block of earlier time windows falls below a certain tolerance criterion, $\eta \in \mathbb{R}_{>0}$. Let $\mathcal{T}^{(i)} - l$ represent the time window threshold at which this happens. Then, tokens of at time windows $\{1,2,\dots,\mathcal{T}^{(i)} - l\}$ are pruned together as one feature, thereby reducing the number of possible coalitions in future KernelSHAP runs. Our selected criterion value was $\eta = 0.025$ based on the recommendations of the original TimeSHAP report.[R11]

<u>Token- and time-level explanations</u>:

Once the pruned time windows $\{1,2,\dots,\mathcal{T}^{(i)} - l\}$ are lumped into a single feature, TimeSHAP then groups each of the tokens across the remaining time windows (i.e., the recent past) as features. In other words, each row of $\mathbf{X}_{:,\mathcal{T}^{(i)}-l+1:\mathcal{T}^{(i)}}^{(i)}$ is grouped as a feature, and these $M$ features (along with the pruned time windows as a single feature) are fed into KernelSHAP to estimate the token-level Shapley values. Thereafter, each of the $l$ remaining time windows – i.e., each column of $\mathbf{X}^{(i)}_{:,\mathcal{T}^{(i)}-l+1:\mathcal{T}^{(i)}}$ – is grouped as a feature, and KernelSHAP is used to estimate the time-level Shapley values for each of them. TimeSHAP also permits estimation of cell-level Shapley values (i.e., at specific combinations of tokens and time windows), but we did not calculate these values for our analyses.

ΔTimeSHAP:
The ordinal endpoint of our dynamic model is itself a dynamic variable. Therefore, we were interested in using TimeSHAP to uncover features associated with changes in next-day TIL$^{\text{(Basic)}}$.

Let $t^* \in \{1,2,\dots,\mathcal{T}^{(i)}\}$ denote a day at which the next-day TIL$^{\text{(Basic)}}$ score is different from the last available TIL$^{\text{(Basic)}}$ score. With the TimeSHAP algorithm, we estimated Shapley values ($\phi$) for each token $j \in \{1,2,\dots,M\}$ in two days directly preceding a day-to-day change in TIL$^{\text{(Basic)}}$ (i.e., $\{t^*, t^*-1\}$) to calculate:

$$\Delta\phi^{(i)}_{j,t^*} = \phi^{(i)}_{j,t^*} - \phi^{(i)}_{j,t^*-1},$$

which we refer to as the token's ΔTimeSHAP value. If a token did not exist in the window of either of the two days, then its $\phi$ value for that day was zero. Assuming the population-average model output $\left(\mathbb{E}_{\mathbf{X}}\left[\widehat{f_\omega}(\mathbf{X})\right]\right)$ does not change substantially between the two days, ΔTimeSHAP values can be interpreted as **a token's contribution to the difference in an individual patient's model output over the two days directly preceding the change in TIL$^{\text{(Basic)}}$, given the patient's full set of tokens**. If a variable had a positive (or negative) ΔTimeSHAP value, it was associated with an increased likelihood of escalation (or de-escalation) in next-day treatment intensity. Moreover, since the calculation of ΔTimeSHAP values required two days of information before the change in TIL$^{\text{(Basic)}}$, we only calculated the variable contributions to day-to-day changes in TIL$^{\text{(Basic)}}$ that occurred after day two of ICU stay.
